# Improving Pre-eclampsia Risk Prediction by Modeling Individualized Pregnancy Trajectories Derived from Routinely Collected Electronic Medical Record Data

**DOI:** 10.1101/2021.03.23.21254178

**Authors:** Shilong Li, Zichen Wang, Luciana A. Vieira, Amanda B. Zheutlin, Boshu Ru, Emilio Schadt, Pei Wang, Alan B. Copperman, Joanne Stone, Susan J. Gross, Eric E. Schadt, Li Li

**Affiliations:** Sema4, Stamford, CT, USA; Department of Obstetrics, Gynecology, and Reproductive Science, Icahn School of Medicine at Mount Sinai, New York, NY, USA; Reproductive Endocrinology and Infertility, Reproductive Medicine associates of New York, New York, USA; Department of Genetics and Genomic Sciences, The Icahn Institute for Genomics and Multiscale Biology, Icahn School of Medicine at Mount Sinai, New York, NY, USA

**Author notes:** These authors contributed equally to this work (Shilong Li, Zichen Wang).

## Abstract

Preeclampsia (PE) is a heterogeneous and complex disease associated with rising morbidity and mortality in pregnant women and newborns in the US. Early recognition of patients at risk is a pressing clinical need to significantly reduce the risk of adverse pregnancy outcomes. We assessed whether information routinely collected and stored on women in their electronic medical records (EMR) could enhance the prediction of PE risk beyond what is achieved in standard of care assessments today. We developed a digital phenotyping algorithm to assemble and curate 108,557 pregnancies from EMRs across the Mount Sinai Health System (MSHS), accurately reconstructing pregnancy journeys and normalizing these journeys across different hospital EMR systems. We then applied machine learning approaches to a training dataset from Mount Sinai Hospital (MSH) (N = 60,879) to construct predictive models of PE across three major pregnancy time periods (ante-, intra-, and postpartum). The resulting models predicted PE with high accuracy across the different pregnancy periods, with areas under the receiver operating characteristic curves (AUC) of 0.92, 0.83 and 0.89 at 37 gestational weeks, intrapartum and postpartum, respectively. We observed comparable performance in two independent patient cohorts with diverse patient populations (MSH validation dataset N = 38,421 and Mount Sinai West dataset N = 9,257). While our machine learning approach identified known risk factors of PE (such as blood pressure, weight and maternal age), it also identified novel PE risk factors, such as complete blood count related characteristics for the antepartum time period and ibuprofen usage for the postpartum time period. Our model not only has utility for earlier identification of patients at risk for PE, but given the prediction accuracy substantially exceeds what is achieved today in clinical practice, our model provides a path for promoting personalized precision therapeutic strategies for patients at risk.

## Introduction

Preeclampsia (PE) remains one of the great challenges in obstetrics. It contributes substantially to maternal morbidity and maternal mortality worldwide, and within the US, accounted for 6.9% of pregnancy-related deaths from 2011 to 2016 (CDC Reproductive Health: Maternal Mortality) and is substantially higher in other regions. There are significant implications for newborns as well, with PE being responsible for a large percentage of medically indicated preterm deliveries

PE is characterized by elevated blood pressure during pregnancy, starting after 20 gestational weeks. While moderately elevated blood pressure itself is not necessarily harmful, in the case of PE, elevated blood pressure reflects the multi-system endothelial dysfunction leading to vascular, renal and liver impairment associated with this disease. Eclampsia, defined as convulsions during pregnancy and/or postpartum irrespective of hypertension, is an especially devastating outcome and may be associated with maternal hypoxia and death. The underlying mechanisms are not fully understood but recent evidence suggests involvement of multiple factors and pathways, including maternal factors and abnormal trophoblast differentiation ^2^. This underlying complexity helps to explain the unpredictable nature of PE. PE can vary not only in severity, but also in timing of onset and impact on fetal growth. Although there are serious clinical sequalae due to PE, antenatal monitoring to determine when delivery outweighs the risk of ongoing expectant management delivery is the standard clinical care plan for PE patients, given deliver is currently the only recognized treatment for PE.

Currently, women are routinely screened for PE at the first prenatal visit using clinical factors. Some centers may also include serum protein markers and ultrasound Doppler studies to screen for early PE. During subsequent visits, blood pressure and proteinuria screening are conducted. Ideally, improved screening could direct clinical care through increased prenatal surveillance and adoption of prophylactic measures, such as low dose aspirin that has been shown to reduce risk of preterm PE and potentially other perinatal complications ^3^ (ACOG Committee Opinion No. 743; USPSTF, 2017). In addition, accurate identification of risk could allow for escalation to a higher level of care facility for delivery. However, there is still a lot of room for improvement with respect to PE screening. The genome, transcriptome, proteome, and metabolome have all been interrogated and have generated some promising data ^4–9^. However, currently there are no omics-based biomarkers available for clinical use. Furthermore, all the current screening methodologies focus on a relatively small number of maternal characteristics, and usually just one time point at early pregnancy that remains the same over the course of gestation ^10^. Considering the number of prenatal visits that occur over a well-defined time range, there remains an unmet need for longitudinal PE assessment at each encounter that accounts for changes in clinical measurements within an individual’s characteristics throughout pregnancy. With PE rates rising along with maternal mortality in the U.S. ^11^, a more robust approach that can predict antenatal, intrapartum and postpartum PE is still very much needed.

To the best of our knowledge, large-scale EMR data have not been systematically mined to identify novel features associated with PE risk and to model these data using machine learning approaches to establish whether this wealth of longitudinal, high-dimensional patient-level data contained in EMRs can improve PE risk prediction. The increasing accessibility of large-scale EMR data integrates laboratory-based molecular and biochemical tests, disease diagnoses, procedures, and prescriptions, along with outcomes during the pregnancy journey. Further, abstracting patient journey information from these records, normalizing the data across systems, reconstructing pregnancy journeys, and modeling these journeys using state of the art data analytic approaches that can account for dynamic state changes provides for the potential to better model PE risk through the course of pregnancy, compared to what is achieved in today’s standard of care.

Here we build predictive models from digitally reconstructed pregnancy journeys derived from the EMR data from the Mount Sinai Health System in New York City, among the largest and most diverse health systems in the U.S., to assess the risk of PE across 17-time points throughout the antepartum, intrapartum and postpartum periods of pregnancy. Appropriately curated pregnancy journeys derived from EMR data provide a more expansive, feature-rich context in which to study the pathophysiology of PE towards constructing more predictive models to identify patients at risk for PE. After identifying 83,954 patients with pregnancies represented in the MSHS EMR, we reconstructed the full longitudinal health course through these pregnancies (referred to as pregnancy journeys) using a pregnancy journey construction algorithm resulting in the identification of 80,021 patients in which 108,557 complete pregnancy journeys were captured by the EMR. We then developed a digital PE phenotyping rules-based algorithm based on clinical criteria established by the American College of Obstetricians and Gynecologists (ACOG)^12^ to identify patients diagnosed with PE at different periods of their pregnancy. With complete pregnancy journey information and the PE diagnosis labels, we constructed predictive models at 19 different time points across the three major pregnancy time periods (ante-, intra-, and postpartum) by applying state-of-the-art statistical and machine learning methods to data collected for patients throughout their pregnancy journey. We validated the predictive models we trained using data from one hospital within the MSHS, and another independent dataset derived from other hospitals within the MSHS. Our PE risk assessment model could be applied in clinical practice by extracting the relevant input features for the model from the patients’ electronic medical records and running the model on those data. Furthermore, using different approaches to interpret predictions, we reveal the connections between clinical features and PE risk to help understand the potential research areas for exploring pathophysiology of PE.

## Results

### Reconstructing Pregnancy Journeys from Electronic Medical Record Data

One of the limitations of current-day EMR systems in widespread use is that they do not naturally capture and represent patient journeys through specific episodes through a patient’s health course. EMR systems are transactional, automating the capturing of a patient visit and recording of the clinical measures, labs, procedures, and prescriptions generated on a patient over the course of their visit. Most EMRs in widespread use are not set up to provide a longitudinal view of a patient along a particular health course journey such as pregnancy with all of the corresponding data generated on the patient over that journey. Thus, we developed a pregnancy journey construction algorithm to identify 83,954 patients with 114,312 pregnancies represented in the MSHS EMR systems and to reconstruct 108,557 full pregnancy journeys of 80,021 patients between 2002 and 2019 (see supplemental material).

### Patient characteristics across a training and two independent test datasets

We retrieved all relevant clinical characteristics on the patients in this dataset, including demographics, diagnoses, drug prescriptions, procedures, vital signs and lab values (Fig 1A). In total we captured 3230, 4136 and 5391 clinical features for ante-, intra- and postpartum, respectively, represented in the EMR on these patients and 46,725,028 datapoints overall, providing among the most comprehensive datasets available in the context of the pregnancy journey, to take a more data-driven approach to evaluating PE risk. Women delivered at one of two main inpatient facilities, Mount Sinai Hospital (MSH) and Mount Sinai West (MSW). We split patient journeys collected from MSH into a training set (N=60,879) and a test set (N=38,421) irrespective of the timing, and we used MSW (N=9,257) from a different geographic region in NYC as a second test set.

**Fig 1.**
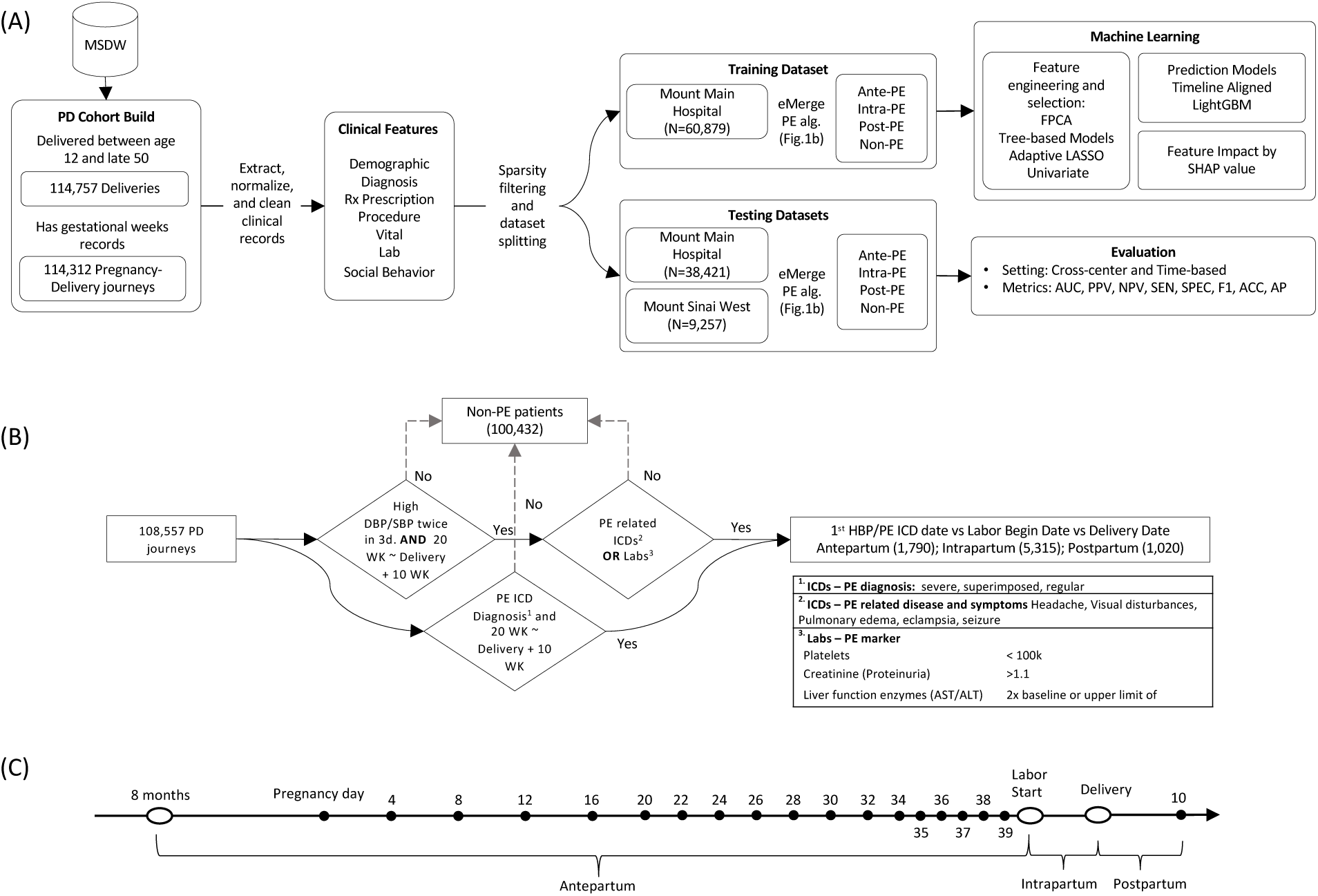
Overview of study design and model development. **(A)** The workflow of the study outlines the cohort construction, patient characteristics extraction, dataset splitting into training and testing datasets (including subdivision into antepartum, intrapartum and postpartum), machine learning model development and final evaluation. **(B)** The proposed eMerge algorithm to identify preeclampsia patients to construct the binary prediction problem. **(C)** The schematic of 19 timeline models including: monthly models, weeks 4-20; biweekly models, weeks 22-34; weekly models, weeks 35-39; intrapartum and postpartum model.

To identify patients diagnosed with PE during the course of their pregnancy from these datasets, we developed and applied a rule-based digital phenotyping algorithm (Fig 1B) to identify 5,663 (9.3%) PE patients from the 60,879 patients in the training dataset. We further identified 2,064 (5.4%) PE patients from the MSH test dataset and 398 (4.3%) PE patients from the MSW test dataset, respectively. The PE prevalence observed across the various datasets is consistent with prior published literature: 2%-8% in the general population ^13^.

Patient demographics and characteristics collected 8 months prior to pregnancy as baseline were significantly different between the MSH training set, the MSH test set, and the MSW test set, indicating differences in regional geographic and socioeconomic status, and shifting demographics over time. More detailed summaries of the characteristics of these different datasets are provided in Table 1, where we note statistically significant differences with respect to Medicaid rates, population composition, and average pregnancy ages, among several other features, between the different datasets.

**Table 1.** Characteristics of patients in MSH training dataset, MSH and MSW test dataset.

| Cohorts | Mount Sinai Hospital (MSH) training set | Mount Sinai Hospital (MSH) test set | Mount Sinai West/UW/BI/SL test set | P-Value |
| --- | --- | --- | --- | --- |
| Features at Baseline | 60879 | 38421 | 9257 |  |
| Age at pregnancy, median [Q1,Q3] | 31.0 [26.0,35.0] | 31.0 [27.0,35.0] | 33.0 [29.0,36.0] | <0.001 |
| Weight (kg), median [Q1,Q3] | 63.5 [56.0,74.4] | 63.5 [56.2,74.4] | 63.0 [56.2,72.6] | 0.084 |
| Height (cm), median [Q1,Q3] | 162.6 [157.5,167.6] | 162.6 [157.5,167.6] | 163.8 [160.0,168.9] | <0.001 |
| BMI (kg/m^2), median [Q1,Q3] | 23.8 [21.1,28.0] | 24.0 [21.2,28.1] | 23.2 [20.9,26.6] | <0.001 |
| SBP (mmHg), median [Q1,Q3] | 110.0 [104.0,120.0] | 112.0 [106.0,120.0] | 112.5 [108.0,120.0] | <0.001 |
| DBP (mmHg), median [Q1,Q3] | 67.5 [60.0,72.0] | 69.0 [62.0,74.0] | 70.0 [66.0,76.0] | <0.001 |
| **Race, n (%)** |  |  |  |  |
| African-American/Black | 8336 (13.7) | 3246 (8.4) | 1399 (15.1) | <0.001 |
| Asian | 4623 (7.6) | 2754 (7.2) | 1123 (12.1) |  |
| Caucasian/White | 32662 (53.7) | 25686 (66.9) | 4622 (49.9) |  |
| Hispanic/Latino | 9928 (16.3) | 3253 (8.5) | 280 (3.0) |  |
| Multi-racial | 589 (1.0) | 208 (0.5) | 110 (1.2) |  |
| Native American | 227 (0.4) | 111 (0.3) | 73 (0.8) |  |
| Other | 4112 (6.8) | 2386 (6.2) | 1090 (11.8) |  |
| Unknown | 402 (0.7) | 777 (2.0) | 560 (6.0) |  |
| Medicaid, n (%) | 22773 (37.4) | 9956 (25.9) | 1362 (14.7) | <0.001 |
| Miscarriage history, n (%) | 1167 (1.9) | 331 (0.9) | 299 (3.2) | <0.001 |
| PE history, n (%) | 350 (0.6) | 140 (0.4) | 28 (0.3) | <0.001 |
| Smoking history, n (%) | 5876 (9.7) | 2135 (5.6) | 544 (5.9) | <0.001 |
| Alcohol use history, n (%) | 8941 (14.7) | 3902 (10.2) | 546 (5.9) | <0.001 |
| **hospital, n (%)** |  |  |  |  |
| Mount Sinai Hospital - Main Hospital | 60879 (100.0) | 38421 (100.0) |  | <0.001 |
| Mount Sinai West/UW/BI/SL |  |  | 9257 (100.0) |  |
| **PE type, n (%)** |  |  |  |  |
| Antepartum | 1213 (2.0) | 437 (1.1) | 140 (1.5) | <0.001 |
| Intrapartum | 3787 (6.2) | 1322 (3.4) | 206 (2.2) |  |
| None | 55216 (90.7) | 36357 (94.6) | 8859 (95.7) |  |
| Postpartum | 663 (1.1) | 305 (0.8) | 52 (0.6) |  |
| Pregnancy journey length (days), median [Q1,Q3] | 273 [266,280] | 275 [267,281] | 277 [270,282] | <0.001 |

### Performance of predictive model across pregnancy in training set

In order to train predictive models for PE along the pregnancy journey, we divided the journey up into 19 time points that included dividing the antepartum period into 17 time points following a standard of care protocol for prenatal office visits at the participating site: 5 monthly visits spanning weeks 4-20, 7 biweekly visits spanning weeks 22-34, and 5 weekly visits spanning weeks 35-39 ^14, 15^; followed by intrapartum and postpartum periods as two independent time points with respect to the pathophysiology of PE (Fig 1C). Given the large number of clinical features available from the EMR database for our datasets, for each of the 19 time points we employed several feature selection methods to choose features robustly that were consistently significantly different between patients diagnosed with PE and those without PE. Several features demonstrated consistently changing effects throughout the pregnancy (Fig 2A), reinforcing the importance of partitioning the antepartum period into more granular time points to better isolate signals that may associate with the clinical manifestation of PE. For the monthly models (spanning weeks 4-20), our feature selection process identified between 19 and 36 unique features depending on the month; between 34 and 40 unique features for the biweekly models (weeks 22-34); and 35 to 40 unique features for the weekly models (weeks 35-39). We also selected 68 and 48 unique features, respectively, for the intrapartum and postpartum periods. All of the selected unique features across the 19 time points are summarized in Table 1-19 in the Supplement. For each of the 19 time points, we trained gradient-boosting models and tuned the parameters of these models using cross-subject validation. The cross-subject validation performance for each time point is summarized according to the area under the receiver operating characteristic curve (Figure 2B; AUC), the positive predictive value (Figure 2C; PPV), the negative predictive value (Figure 2D; NPV) and specificity (Figure 2E; SPE). These performance measures assess the diagnostic ability of the models (AUC) as well as the sensitivity and specificity of the models taking into account the population prevalence of the disease (PPV and NPV). For predictive performance comparison, we also built the ACOG criteria-based model based on risk factors constructed from patient characteristic and medical history recommended by ACOG^12^ (see Methods).

**Fig 2.**
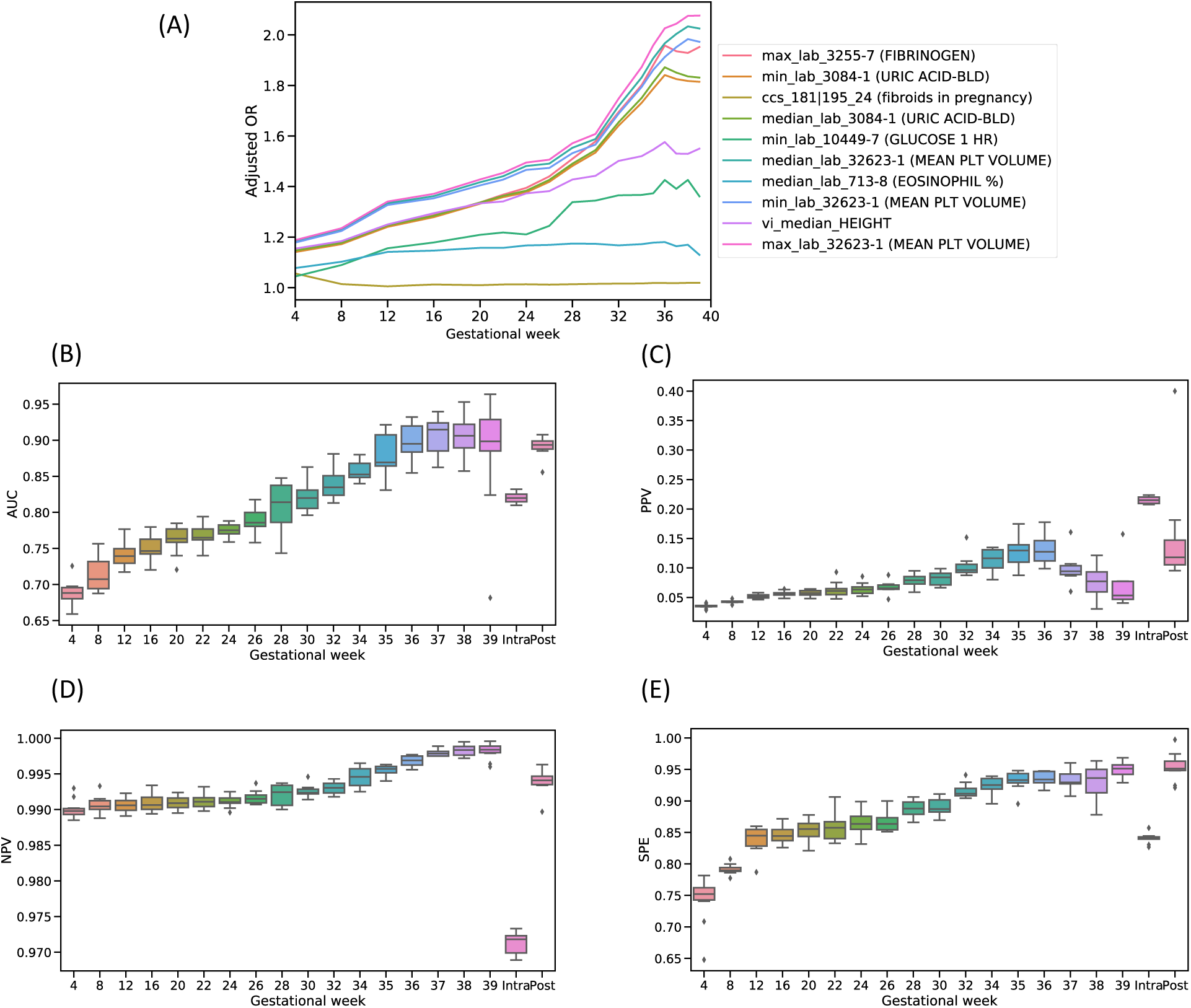
Model performance at different time points. **(A)** Features indicate different dynamical signals across the gestational weeks based on different adjusted odds ratio (OR). **(B)** Area under receiver operating characteristic curve (AUC) score for each time point. **(C)** Positive predictive value (PPV), along with preeclampsia risk in the population, at each time point. **(D)** Negative predictive value (NPV) at each time point. **(E)** Specificity (SPE) at each time point. The variation estimates were derived from 10-folds cross-subject validation from training set.

As the density of data increased across the antepartum period, the median AUC score increased from 0.69 (interquartile (first quartile-third quartile) [IQ]: 0.68-0.70) at week 4 where most of clinical attributes obtained from the patient’s historical information, to 0.92 (IQ: 0.89-0.92) at week 37, which captures nearly all feature values through the pregnancy course. We calculated an median AUC score of 0.82 (IQ: 0.82-0.83) for intrapartum and 0.89 (IQ: 0.89-0.90) for postpartum in the cross-subject validation analysis. In comparison, the ACOG criteria-based model for antepartum achieved a median AUC score of 0.62 (IQ: 0.62-0.63) with high risk factors and 0.67 (IQ: 0.67-0.68) using all risk factors. We also compared our model PPVs to existing PE risk assessments used as part of standard of care (i.e., population prevalence during the same gestational week) using our models. For example, the PPV for our model at week 4 was 0.04 (IQ: 0.03-0.04) compared to a prevalence of 0.02 (a greater than 2-fold increase). Similarly, the PPV for our model at week 37 was 0.094 (IQ: 0.089-0.104) compared to a prevalence of 0.015 (a greater than 8-fold increase). Complete performance summaries across all models are provided at Table 20 in the Supplement.

### Refining key features during the pregnancy journey

We identified 78, 68 and 48 uniquely influential clinical features across the entire antepartum, intrapartum and postpartum periods, respectively (Fig 3A). Twenty-one features were significant predictors in all three periods, and 42, 30 and 15 features, respectively, were specific to antepartum, intrapartum and postpartum. Among the 21 common features, which were enriched for patient demographics and baseline characteristics, we identified 48% features supported in the literature as associating with PE risk, including systolic blood pressure (SBP) ^12, 16^, diastolic blood pressure (DBP) ^17^, weight ^16^, maternal age ^18^, hemoglobin ^19^, white blood cell count ^20^, gestational hypertension ^12^, PE history ^21^, chronic hypertension ^20^ and headache (including migraine) ^22^ (Table 1-19 in the Supplement). Features specific to antepartum were enriched with CBC findings that suggest inflammation and/or infection, such as elevated neutrophil, monocyte, eosinophil, and lymphocyte levels. Intrapartum-specific factors included pregnancy complications such as malposition, malpresentation, premature rupture of membranes (PROM), and sodium chloride (salt) use. Predictors in the postpartum period included many indications relating to follow-up care, such as immunizations, screening for infectious diseases, OB-related trauma, and ibuprofen usage (Fig 3B).

**Fig 3.**
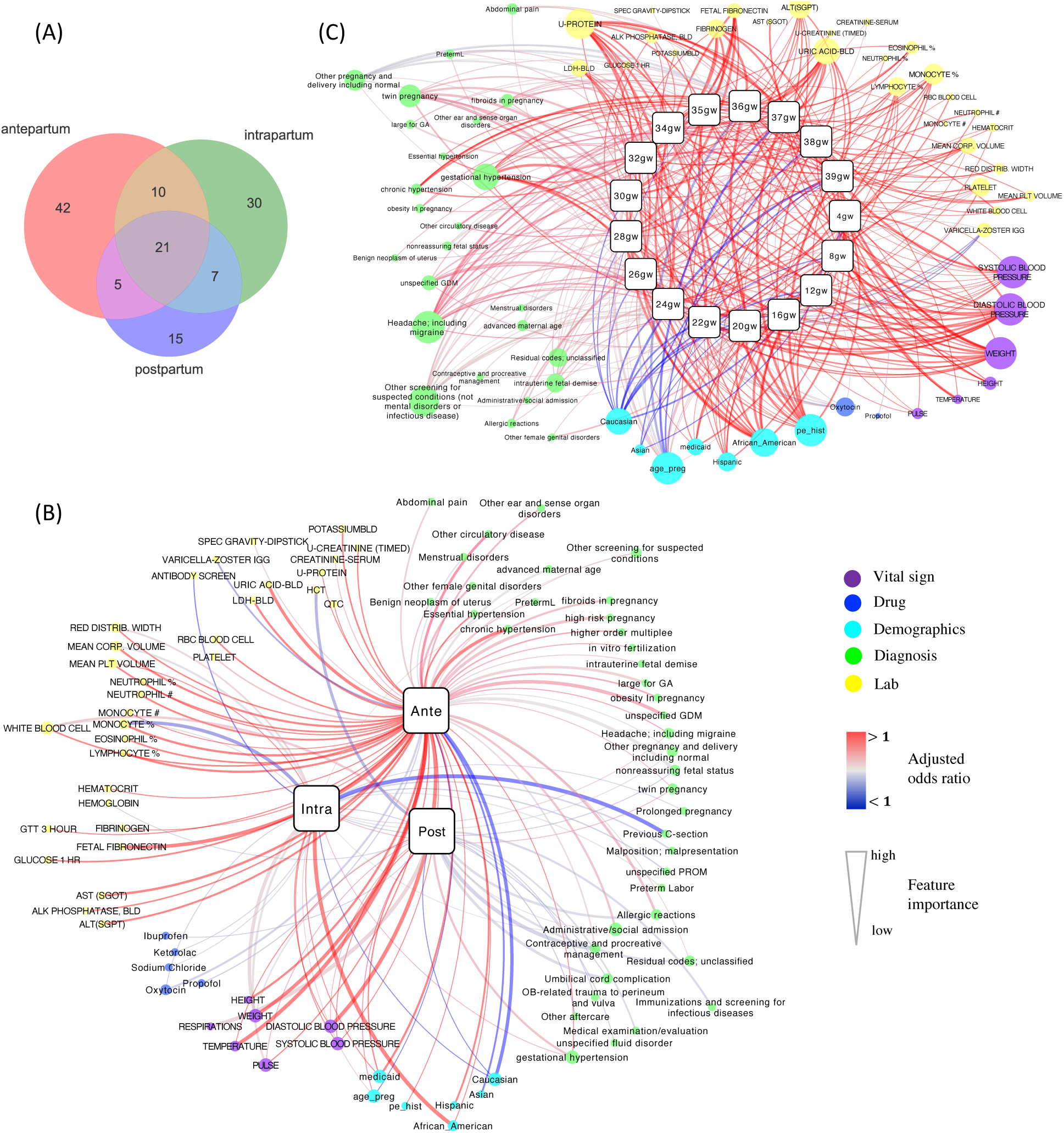
Networks of feature associations through pregnancy. **(A)** Venn diagram to show common features shared with three pregnancy periods, and specific features to each period. **(B)** The network to display the associations of selected clinical features with each pregnancy period. **(C)** The network for the 17 time points in the antepartum. The two networks were constructed by connecting predictive features with respective PE time point. The squares signify different time points of PE, and the round nodes represent the identified predictive features with their sizes proportional to the feature importance. The red edge color indicates the risk association (adjusted OR > 1) while the blue edge color indicates the protective association (adjusted OR < 1). The edge width reflects the significance of predictive features. Different feature categories are represented with different colors and also laid out together. The networks were visualized using Cytoscape 3.7.2.

To better visualize the contributions of the most predictive features across the pregnancy time periods, we further reduced the number of features during intrapartum and postpartum periods to 30 and 24 unique features, respectively, while maintaining the same level of performance (Methods in the Supplement). Associations between each clinical feature and PE were visualized by each time period (Fig 3B), confirming known relationships such as Caucasian and Asian patients being less likely to develop PE, while African American and Hispanic patients were more prone to PE, especially during the intrapartum period (OR:1.25 [95% CI, 1.09-1.43]). Additionally, we identified that patients covered by Medicaid insurance were more likely to develop PE ^23, 24^. Additionally, we have found features that have not been reported before, such as our identification of pulse rate as a risk factor that was consistently associated with PE in each time period.

To further characterize features we identified as predictive for PE risk, we constructed an interaction network of predictive clinical features and PE across the 17 time points within the antepartum period (Fig 3C). From the resulting network, we identified clusters of unique lab test features (N=33), diagnoses (N=28), vital signs (N=8), demographics (N=7), and drug prescriptions (N=2). We confirmed well known risk factors for antepartum PE ^12^ (Table 1-17 in the Supplement). Moreover, we identified PE biomarkers previously reported in the literature, including fibrinogen ^25^, mean platelet volume (MPV) ^26^, mean corpuscular volume (MCV) ^27^, red cell distribution width ^27^, fetal fibronectin ^28^, and lactate dehydrogenase (LDH) ^29^. Finally, we identified potential novel features that have not been previously reported as associated with PE. For example, median value of varicella zoster virus antibody (IgG) titer is lower in PE patients compared to non-PE patients from 12 to 28 gestational weeks of pregnancy.

### Assessing the dynamic progression of PE associated risk features

To better characterize the dynamic progression of PE features, we generated moving average plots for the significant risk factors, revealing interesting patterns of association even among well known risk factors. For example, while abnormally high SBP is a well-known risk factor used as a diagnostic marker for PE ^16^, by examining longitudinal SBP measures across more than 100,000 pregnancy journeys, the data show that patients who developed PE in the antepartum period generally had elevated SBP measurements compared to patients without PE, even though the elevated measures fall within a normal range and would not be classified as abnormal during a clinical office visit (Fig 4A). The average SBP for PE patients in the antepartum time period was only ∼120 mmHg ^30^, but then consistently through the antepartum period 10 mmHg (one standard deviation of mean from control) higher compared to the control cohort, an important predictive signal for PE picked in nearly all of the models. DBP showed a similar pattern, albeit at a reduced signal strength compared to SBP ^16^. Similarly, while protein in urine (U-Protein) is also a well-established diagnostic marker for PE, our data show that the presence of protein in urine even in trace amounts, is a significant predictor for antepartum PE (Fig 4B). As with SBP, the trace urinary protein levels were supported by our models as a significant predictive feature of PE, even though on their own, recorded at a single visit rather than a longitudinal pattern, trace levels would not be deemed as relevant in current clinical practice.

**Fig 4.**
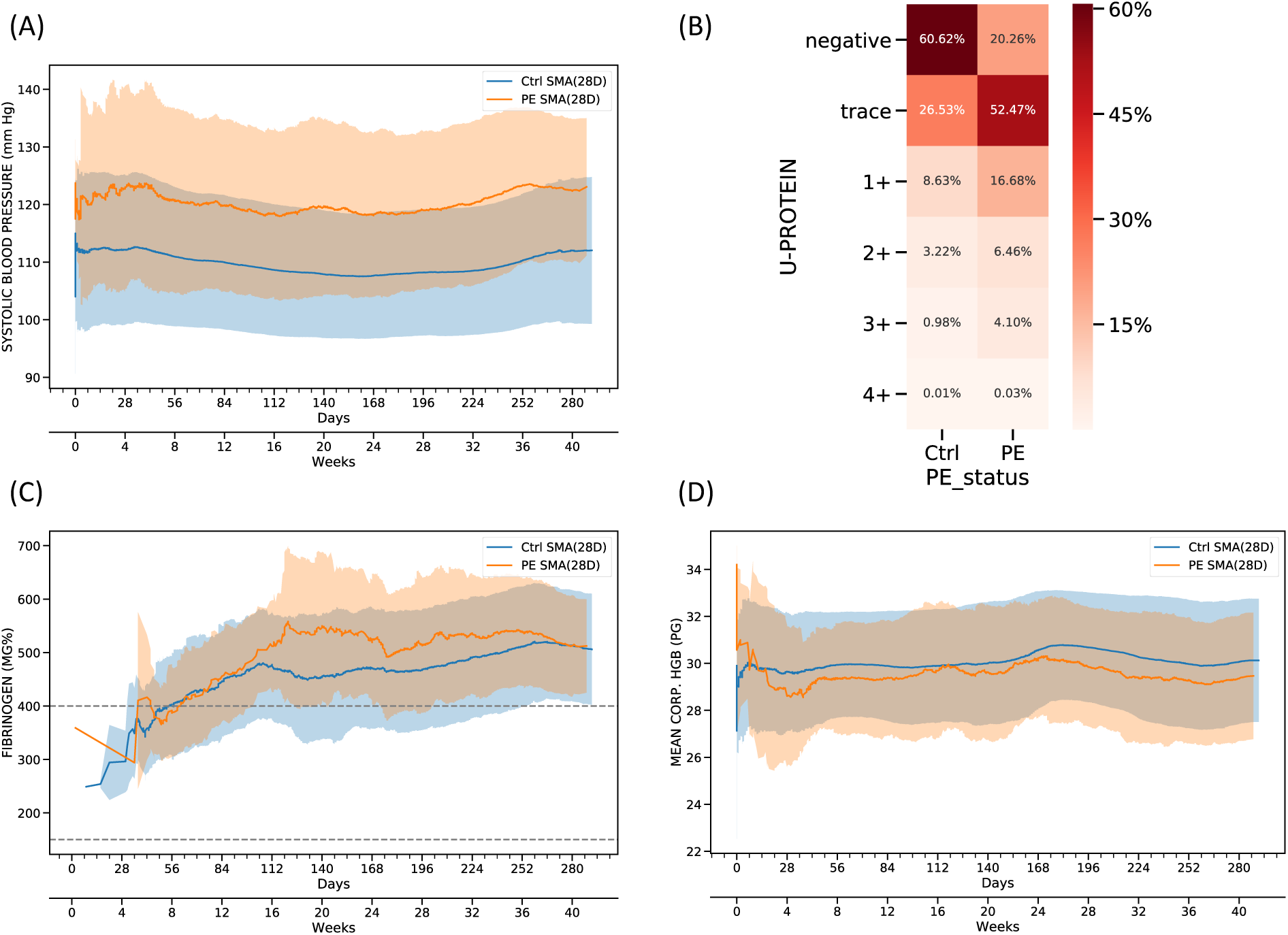
Feature inspection for antepartum based on moving average. **(A)** 28 days moving average of systolic blood pressure for PE and control patients. **(B)** Distribution of urine protein for PE and control patients. **(C)** 28 days moving average of fibrinogen for PE and control patients. The dashed line represents the reference ranges for fibrinogen. **(D)** 28 days moving average of mean corpuscular hemoglobin (HGB) for PE and control patients. In the moving average plots, the shaded areas indicate the standard deviation and solid lines represent the average value across the pregnancy.

In addition to the physiologic and urinary findings, our antepartum models also identified and quantified several biomarkers scored in routine laboratory tests, including fibrinogen, blood uric acid, and mean platelet volume (Fig 3C). Each of these biomarkers exhibited increasing effect sizes in PE cases compared to controls, as measured by adjusted odds ratio over the course of pregnancy journey. These results suggest the corresponding clinical factors predict a greater risk of antepartum PE onset during the later periods of pregnancy. As an example, fibrinogen has been previously associated with PE (especially early onset) ^31^. By examining the moving average of fibrinogen along the course of the antepartum time period, we found that the levels of fibrinogen exhibited a moderate increase at 16 weeks in patients who later developed PE (Fig 4C), suggesting that fibrinogen could be closely monitored over time to enhance the prediction of PE. Along with enhanced utility of known PE risk factors by examining signals longitudinally, mean corpuscular hemoglobin (HGB) was found to be a novel predictor of PE, with slightly lower values observed throughout the antepartum time period in patients who later developed PE (Fig 4D). Taken together, our PE prediction models were able to recover known and novel clinical factors that enhanced power to predict PE.

### Intrapartum features prioritized by importance based on SHAP values

We utilized the framework, SHAP values ^32, 33^, to prioritize the feature contributions to PE predictions by averaging feature importance estimates (Fig 5A). Median SBP measured in antepartum was the most predictive feature for PE in the intrapartum period followed by Caucasian race and oxytocin administration. We also calculated the average contribution of each clinical category for PE predictions (Fig 5B). We found medications provided 40.76% predictive power, demographics 22.82%, vital sign contributing 17.40% followed by diagnoses (13.24%), labs (5.64%) and procedures (0.14%). To uncover the relationship between PE risk and changes in specific feature characteristics, we explored dependence plots which show relative risk (RR) against feature values. To illustrate this point, we provided a representative selection (Fig 5C), demonstrating PE relative risk in terms of antepartum maximum SBP values and the interaction with African American race. We observed that maximum SBP antepartum tended to become a risk factor after 130 mmHg, and the relative risk values changed rapidly around 130 mmHg.

**Fig 5.**
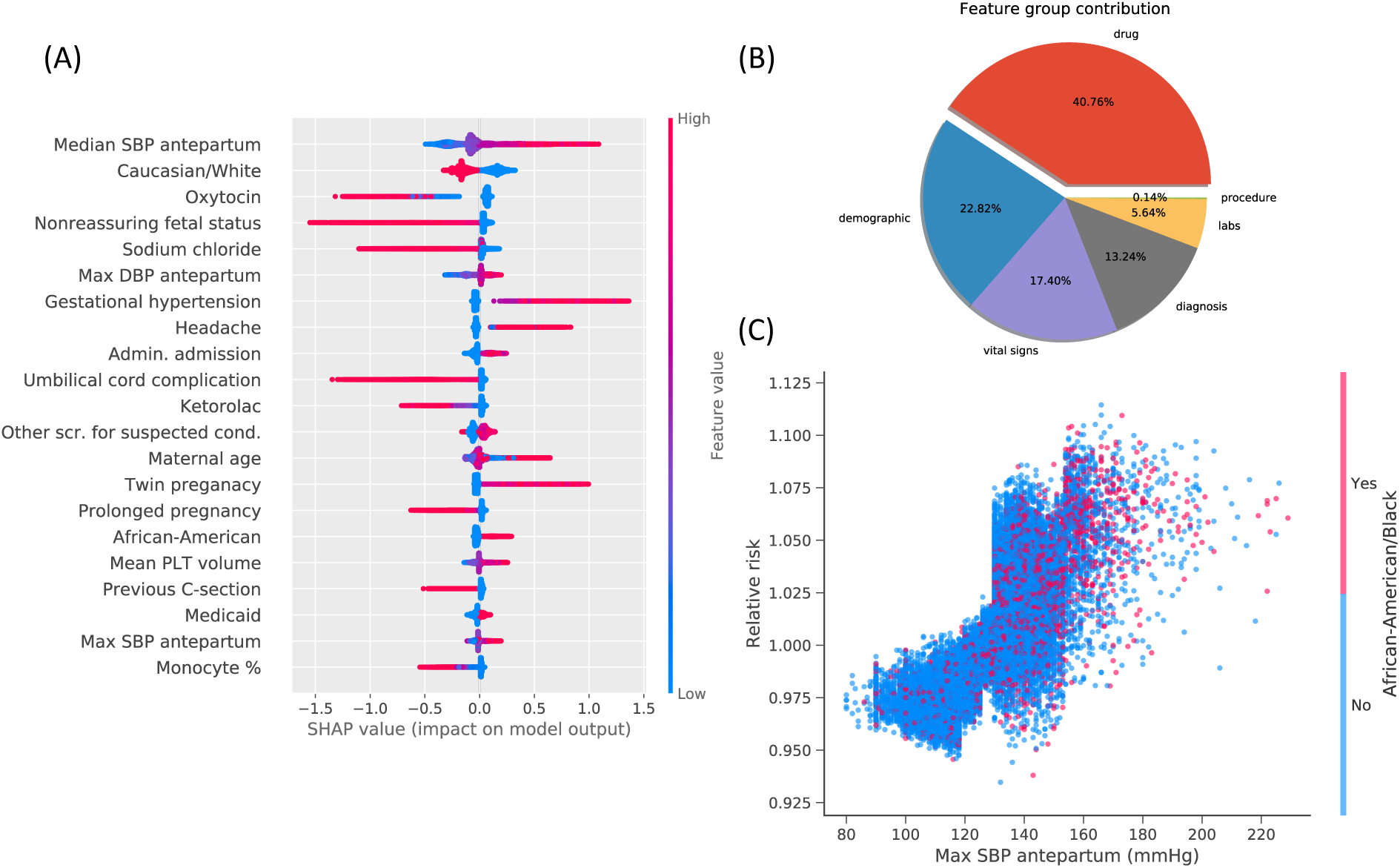
Feature inspection for intrapartum based on SHAP value. **(A)** Top 20 features with highest mean absolute SHAP values. The horizontal axis represents the SHAP value (greater than zero means the risk association; less than zero indicates the protective association) and the vertical axis shows SHAP value distribution for each feature. The color bar indicates the feature values. **(B)** The average feature group contribution calculated from averaging mean absolute SHAP values for each feature set. **(C)** The dependence plot with maximum SBP measured in antepartum versus PE relative risk, along with the interaction of African American race.

### Postpartum features reveal novel medication effects related to racial disparities

In postpartum period, ibuprofen was the best predictor for PE risk, followed by maximum and median SBP measured during postpartum (Fig 6A). Both Caucasian race and OB-related trauma showed protective benefit for PE risk reduction. OB-related trauma is common among vaginal deliveries, so this feature likely reflects the protective effect of a vaginal delivery relative to a Cesarean delivery. As a category, medications provided the highest average predictive contribution (46.83%), followed by diagnoses (15.39%), demographics (14.33%), lab tests (10.30%), and procedures (0.08%) (Fig 6B).

**Fig 6.**
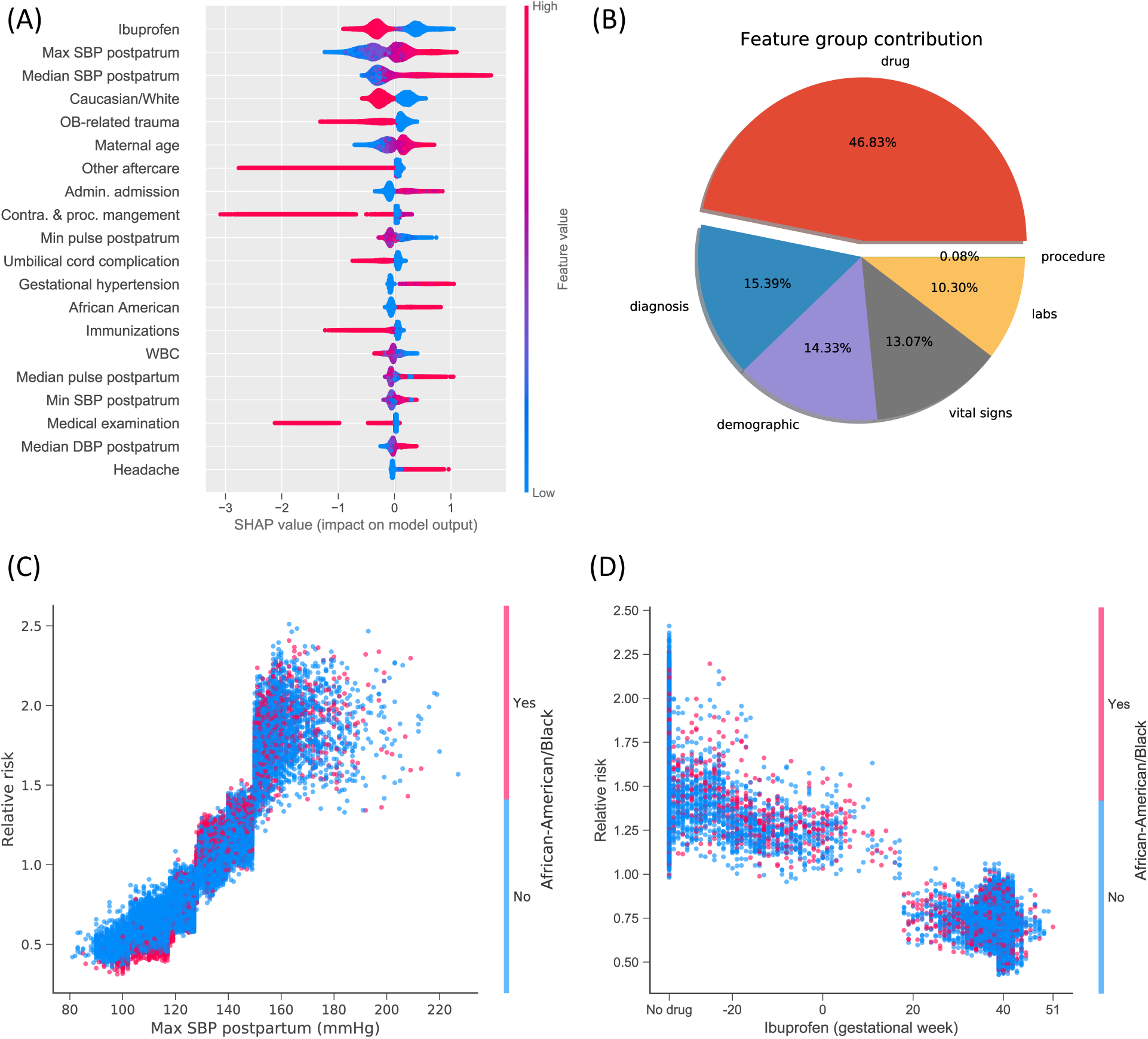
Feature inspection for postpartum based on SHAP value. **(A)** Top 20 features with highest mean absolute SHAP values. **(B)** The average feature category contribution. **(C)** The dependence plots of PE relative risk in terms of maximum SBP measured in postpartum. **(D)** The dependence plot of PE relative risk versus ibuprofen.

Among predictive features during the postpartum period, we observed that maximum SBP measured in postpartum had a clear effect on the risk of PE (Fig 6C). The risk of PE increased almost linearly as the elevation of SBP until around 150 mmHg where relative risk steeply increased. Evidently, maximum SBP postpartum would become a risk factor when it exceeded 130 mmHg. Among the patients with maximum SBP ranging from 130 mmHg to 150 mmHg, African American patients were at higher odds to develop PE compared to other races. Among the 18,214 pregnancy journeys in this range, 2,978 were African American patients. Within African American race group, the ratio of patients with PE risk (RR >=1) to those without PE risk (RR <1) was 12.23 while the ratio within other race groups was 3.63. Interestingly, the protective effect of ibuprofen appeared limited to this time period and may increase risk used prior to pregnancy (Fig 6D).

### PE predictive model validated in two independent data sets at Mount Sinai Health System

We tested the external validity of our predictive models using two independent datasets, a withheld test set from Mount Sinai Hospital (MSH) and all data collected from Mount Sinai West (MSW). Demographic and clinical characteristics were reported in Table 1 and PE prevalence for each period is listed in Table 21 in the Supplement. We evaluated performance in these two datasets using four predictive performance metrics, AUC, PPV (positive predictive value), NPV (negative predictive value) and specificity (SPE) (Fig 7). Other detailed metrics are reported in Table 22 and 23 in the Supplement.

**Fig 7.**
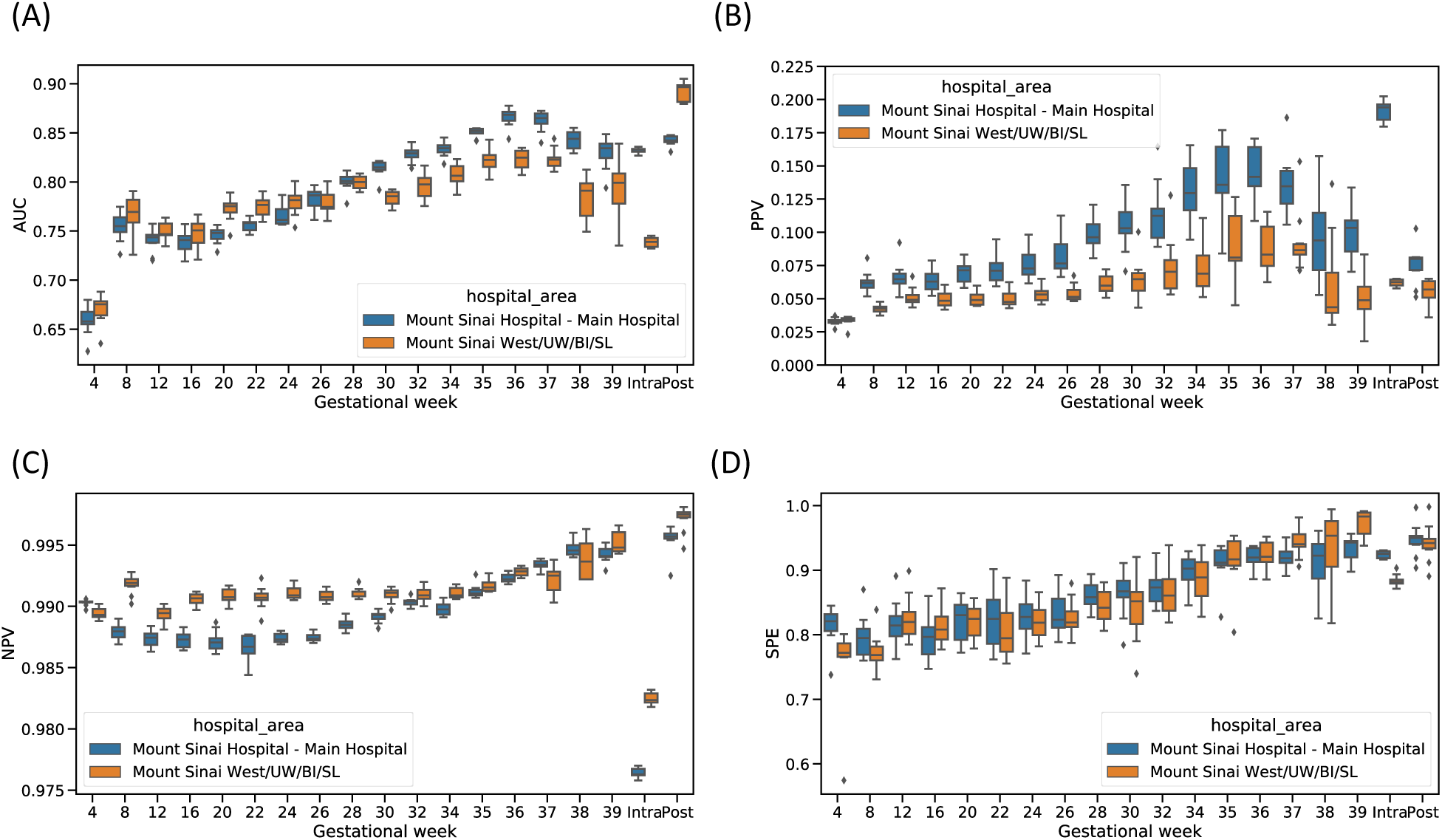
Model validation on two independent datasets in MSHS. **(A)** Area under receiver operating characteristic curve (AUC) score for each time point. **(B)** Positive predictive value (PPV), along with preeclampsia risk in the population, at each time point. **(C)** Negative predictive value (NPV) at each time point. **(D)** Specificity (SPE) at each time point. Blue curve indicates the validation in MSH testing set and yellow curve represents the validation in MSW dataset.

For the MSH test set, we achieved an AUC of 0.66 (IQ: 0.65-0.67) at week 4, which rose continuously as more clinical information became available and reached 0.87 (IQ: 0.86-0.87) at week 37. Consistent with this trend, our intrapartum and postpartum models had AUC scores of 0.83 (IQ: 0.83-0.84) and 0.84 (IQ: 0.84-0.85), respectively. In comparison, we also assessed prediction performance from ACOG criteria-based model for antepartum, AUC score was 0.58 (IQ: 0.58-0.59) using high risk factors, and AUC score was 0.66 (IQ: 0.65-0.67) using all risk factors. All of our models had much higher precision than case prevalence. The lowest PPV we reported was at our first timepoint, week 4, where a PPV of 0.033 (IQ: 0.032-0.034) was observed and case prevalence was 0.013. By week 37, we observed a PPV of 0.16 (IQ: 0.12-0.15) and case prevalence was 0.007. For intrapartum (prevalence = 0.035) and postpartum (prevalence = 0.008), the PPV was 0.19 (IQ: 0.18-0.20) and 0.08 (IQ: 0.07-0.08), respectively. The median NPV scores for all the periods were at or above 0.98 (Table 22 in the Supplement). The SPE within antepartum was 0.82 (IQ: 0.81-0.83) at week 4 and boosted to 0.92 (IQ: 0.91-0.93) at week 37. We estimated SPE was 0.92 (IQ: 0.92-0.93) for intrapartum and 0.95 (IQ: 0.94-0.95) for postpartum.

Performance was similar in the MSW test set. AUC score was 0.68 (IQ: 0.66-0.68) at week 4 and increased to 0.82 (IQ: 0.82-0.83) at week 37, compared to 0.58 (IQ: 0.56-0.59) by high risk factors, and 0.64 (IQ: 0.62-0.66) with all risk factors by the ACOG criteria-based model. Intrapartum and postpartum had AUC scores of 0.74 (IQ: 0.73-0.74) and 0.90 (IQ: 0.88-0.90). PPVs ranged from 0.034 (IQ: 0.033-0.035) at week 4 to 0.086 (IQ: 0.083-0.091) at week 37 compared to existing PE risk (0.016 at week 4 and 0.011 at week 37). All NPV scores surpassed 0.98 on median for every model throughout pregnancy. More details can be found at Table 23 in the Supplement.

## Discussion

This study represents the first data-driven effort to predict PE events across the entire pregnancy journey (antepartum, intrapartum and postpartum) by comprehensively integrating all clinical characteristics extracted from large-scale EMR data. Our predictive models can identify PE at different time points in accordance with the OB visit protocol, significantly outperforming the ACOG criteria-based model with commonly assessed risk factors. We have tested our developed framework in an independent dataset from a different geographic region and observed comparable performance, demonstrating portability of our PE predictive system to other new facilities.

We captured important features contributing to the PE prediction across pregnancy time points. To provide maximum interpretability to physicians, we calculated moving averages across timepoints for key features, as well as SHAP values to indicate relative importance of individual features to overall risk prediction. We identified features common across all three pregnancy periods and features unique to each period. Specifically, other than common features, CBC related characteristics dominated in antepartum; pregnancy complications associated with intrapartum; follow-up cares impacted postpartum.

Some of the findings give further credence to the underlying mechanisms associated with preeclampsia, especially that of dysregulated inflammatory processes ^34^. We found several laboratory makers that are complementary predictive factors of preeclampsia in our model and are routine available in the antenatal period. Elevated neutrophil, monocyte, eosinophil, and lymphocyte levels were noted in the antepartum time frame. Fibrinogen, aside from being an important molecule in coagulation, also has an important role in inflammation and serves as an acute-phase protein ^35^. Additionally, the temporal relationship of these markers allows for a more specific and nuanced prediction of preeclampsia at multiple time points in pregnancy. Previous prediction models have focused on risk factors at a single time point in pregnancy, such as the ASPRE trial^36^. In contrast, our model accumulates risk factors over time and allows for refined prediction as the pregnancy journey progresses and postpartum. Of interest, ibuprofen was noted to show protective association in the postpartum preeclampsia model, which further the findings from a double-masked randomized trial that ibuprofen did not lengthen the duration of severe-range hypertension in women with PE with severe features ^37, 38^.

Other interesting findings included several insights regarding blood pressure measurements. One advantage to the algorithm is that it does not require any special BP measurements, beyond those commonly performed in the office and recorded in the medical record. Rather than the specific BP measurement, it is the trajectory that consequent signal that drives the algorithm. SBP was a more powerful driver vs DBP, which has been noted previously ^39^. However, in our study, we were able to confirm the importance of SPB >130 mmHg as an important threshold for concern. This finding was readily apparent and lends further support to the those who consider the 140/90 threshold to be too high ^39^, especially in light of the new AHA recommendations ^40^. The association with elevated SBPs and African American race, particularly in the postpartum period, also affirms recent literature that suggests a different BP pattern in African American women following delivery, which warrants further research and assessment. Medicaid has been picked as a significant feature to PE, indicating that these patients may be at increased risk because of limited access to healthcare or other barriers due to low socioeconomic status.

While features associated with inflammatory processes and BP were anticipated, there were other features that could be potentially novel and merits further investigation. The median value of varicella zoster virus antibody (IgG) titer was significantly lower in PE patients compared to non-PE patients from 12 to 28 gestational weeks of pregnancy. This association suggests that higher IgG against varicella zoster, developed from vaccination prior to their pregnancy, or an underlying mechanism, may indicate protective association with PE ^41^.

Our study had several limitations. As our clinical data was extracted from MSH, which is close to other medical centers in the area, patients may have received prenatal care at other nearby hospitals or clinics but then chose to deliver at MSH, resulting in the loss of valuable information from our EMR system. Moreover, patients might not come for follow-up care after discharge. To tackle this issue, we designed sparsity filters to ignore some journeys only with minimal available features, e.g. journeys only with demographics. Nonetheless, even patients receiving care at a single facility will often have missing values. Here, since the gradient boosted tree algorithm can accommodate the missing values, we chose not to explicitly impute them, as this better reflects clinical practice where some patient information might be not available. Additionally, our methodology used ICD9/10 codes to identify maternal comorbidities and excluded detailed physician notes which may have excluded “over the counter” medications or some comorbidities and diagnoses. However, we were well aware of the ICD9/10 code limitations at the outset of this project and therefore chose to use a digital phenotyping approach to identify and confirm PE cases.

Extensive research has identified three important biomarkers for preeclampsia ^42–44^, mean arterial pressure (MAP), uterine artery pulsatility index (UtA-PI) and serum placental growth factor (PlGF). Due to data access restrictions on identifiable data such as ultrasounds, our PE prediction system was developed solely based on structured EMR data. However, we achieved similar or even better prediction performance compared to the models incorporating these biomarkers ^45^. Our methodology allows for the incorporation of biomarkers into our current PE prediction system that would be expected to generate more robust performance. Similarly, certain ‘Omic’ data has shown promise for identifying PE ^4–9^, but this type of data is not collected routinely in a clinical setting. Although further studies are needed to incorporate known biomarkers as well as ‘omics’ data, some of which are still investigational while others such as PlGF are already in clinical use, our algorithm – based on EHR data alone – has significant potential implications for clinical care and management.

Our results showed both common features shared among all periods, and unique features specific to each pregnancy period exist, suggesting significant pathophysiologic differences in each pregnancy period in terms of risk for PE. We have confirmed previously known risks to PE, and also uncovered potentially novel connections between clinical features and PE, some of which are supported by other clinical and experimental data. Furthermore, we have validated our models with similar predictive performance on two independent test data sets with population diversity. The results open the door for optimizing monitoring tools to mitigate risks and for individualizing assessment based on patient risk profiles. In addition, this paper provides the most complete assessment of vital sign patterns and trajectories in patients with and without preeclampsia. We have demonstrated that by harnessing the power of data science, we can enhance predictive PE algorithms throughout the pregnancy journey. Hopefully, with continued research, better screening performance based on precision monitoring strategies, will ultimately lead to preemptive clinical strategies and improved perinatal outcomes.

## Methods

The aim of the study was to develop and validate a prediction tool to screen for and monitor patients at risk for PE using clinical information from 108,557 pregnancies at MSHS in New York City, a large health system with a highly diverse population. We built and implemented a digital phenotype for PE based on ACOG recommendations ^12^ to incorporate multiple diagnostic tests and criteria. We performed data processing, model training and validation, and results interpretation for predicting PE risk and interpreting associations between clinical features and PE.

### Data source and pregnancy journey construction

We utilized de-identified EMR from MSHS. By March 2019, the system contains records for more than 9 million unique patients since 2002. The Mount Sinai EMR covers heterogenous clinical information including patient characteristics, diagnosis, procedure, medications, vital signs, and lab tests for visits. We identified 114,312 deliveries (83,954 unique patients) (Fig 1A), with the average material age of 31.06 (standard deviation [std]: 6.09) at pregnancy (Methods in the Supplement). We extracted, normalized clinical data, and performed quality control (Methods in the Supplement). This data usage is approved by institutional review board (IRB) of Icahn School of Medicine at Mount Sinai: IRB-17-01245.

### Digital phenotyping for PE

The World Health Organization recommended patients meeting the following criteria being diagnosed for preeclampsia: (1) Persistent hypertension and (2) Development of substantial proteinuria ^46^. In Mount Sinai Hospitals, OB/GYNs used diastolic blood pressure (DBP) of 90 mm hg or systolic blood pressure (SBP) of 140 mm hg as the threshold for hypertension. From ACOG guideline, we added the following clinical features: platelets counts, creatinine, liver function enzymes (AST/ALT), proteinuria, and related diseases such as headache, visual disturbances, pulmonary edema, eclampsia, and seizure (Fig 1B).

We implemented a diagnosis and rule-based digital phenotyping algorithm to identify PE patient (Fig 1B). We first selected 2,291 patients who were diagnosed with PE ICD9/10 codes between the gestational weeks of 20 and 10 weeks after the delivery and used the first date of the diagnosis as the PE date. Additionally, we have implemented additional criteria from ACOG guidelines to capture the underdiagnosed PE which were not coded by ICDs (Fig 1B). For 6,279 patients who met both criteria, they were recognized as preeclamptic during the pregnancy-delivery journey. We used the first day of the high blood pressure as the PE date for these patients. If the patient were not diagnosed with PE and did not meet the hypertension – proteinuria defined criteria within pregnancy and until 10 weeks after the delivery, they were defined as the control group. Among 2,291 patients who were diagnosed with PE ICD9/10 codes, we found 91.36% of them have either repeating high blood pressure above the threshold value or diagnosis and lab results of proteinuria. This demonstrates that with comprehensive EMR data and proper digital phenotyping strategy, we can recognize more patients than just using diagnosis codes.

Based on when the preeclampsia occurred, we further split patients into three sub-types: 1,790 patient PE in the ante-partum (before admission for labor and delivery), 5,315 patients in the intra-partum (between admission of labor and delivery and delivery), and 1,020 patients in the post-partum (after delivery).

### Experimental design

We analyzed data for each pregnancy period separately (antepartum, intrapartum, and postpartum) in respect to the pathophysiology of PE. We collected the clinical data between 8 months prior to pregnancy and the cutoff time which was defined differently for control and PE group. The PE onset day was the cutoff time for PE patients in each period, while the delivery day and 10 weeks after delivery represented the cutoff time for the control group in the intrapartum and postpartum, respectively.

We split the data collected from MSH into a training set (60%) and a test set (40%), and trained our models using the training set. For each pregnancy visit in antepartum and each of pregnancy periods, we tuned a set of hyperparameters (learning rate, number of trees, depth of trees, number of leaves, sample rates, L1 and L2 regularization, and number of cases in leaf nodes) with Bayesian optimization approach using ten-fold cross-subject validation and repeated 100 times. Specifically, we developed a cross-subject validation strategy called “StratifiedGroupKFold”. We divided the training set into ten folds with respect to patients, meaning that the pregnancy journeys of a patient could only belong to one fold to avoid the information leakage. Considering our imbalanced labels, we also employed stratified sampling to ensure that relative class frequencies were approximately preserved in each train and validation fold. We used the ten final models to report the median performance and interquartile (first and third quartile). Then, we validated our established models on two independent datasets: the 40% held-out test set from MSH and independent MSW set available until 2019 (including Mount Sinai Beth Israel, Mount Sinai West, Mount Sinai St. Luke and Mount Sinai Upper West). All population characteristics for each data set are shown in Table 1. We have reported AUC, SPE, SEN, PPV and NPV for our model validation performance and comparison with current standard of care, a ACOG criteria-based model (Supplemental tables 20, 22, 23).

### Feature engineering

For diagnoses, drug prescriptions and procedures, we selected the first occurring week as the feature value potentially providing the timing information to the machine learning models compared with the form of binary feature. In order to distinguish the mode of delivery, we identified the journeys associated with the Cesarean section using both diagnosis and procedure codes, and the vaginal delivery by the corresponding procedure codes, thereby leading to two additional features.

We split the vital sign data into three ranges on par with the definition of the three pregnancy periods that might help capture the explicit contribution of pregnancy period information to the model predictions. In each range, we calculated the maximum pain score for the journeys if applicable, and also included the minimum, median and maximum values for other numerical vital sign values observed in the interval. We observed that different journeys involved various lengths of available vital sign data, which increased the difficulty of directly injecting these time-related data into the prediction models. To unify the data length and also account for the time-related information, we applied the functional principal component analysis (FPCA) method ^47^ to features including diastolic blood pressure, systolic blood pressure, O2 saturation, pulse, respirations, temperature and weight. The FPCA method is able to find the functional principal components and their functional principal component scores representing the variations of time series curves explained by the components which naturally keep distinct information in the time series data. We computed the top 10 functional principal component scores with R package fdapace ^48^ as the additional features for the journeys, if available, to interpret the time-related vital sign features.

For lab features, we used the similar process as vital signs that we obtained the maximum ordinal values for journeys and statistical values (minimum, median and maximum) for other numerical lab features in every range. As the functional principal components are approximated with the summation of basis functions, e.g. B-spline, we chose the lab features at least with more than 3 data points to perform the FPCA, otherwise, the program would be aborted. Based on this principle, we finally selected 15 lab features and calculated the top 10 functional principal component scores as the additional features for each selected lab feature.

Collectively, we concatenated all built diagnosis, medications, procedure, vital sign, and lab features along with the demographic features in total of 3230, 4136 and 5391 clinical features for antepartum, intrapartum and postpartum, respectively.

### Feature selection

Since we attained a large volume of features, it is prone to make our final predictive models be overfitted to the training data. Consequently, we adopted three feature selection methods, panelized logistic regression with the adaptive LASSO ^49^, univariate analysis and tree-based models (including XGBoost and random forest) ^50^, to robustly identify the most relevant features in terms of our target. Given that we marked diagnosis, drug and procedure features as zeros if the codes were not available in the journeys, we thus performed the adaptive LASSO on the diagnosis, drug and procedure features to recognize the important features with respect to sparse coefficients and the corresponding p-values. Considering the proportion of missing values in vital sign and lab features, we utilized the univariate analysis to obtain the coefficients and p-values. Specifically, we combined all the demographic features with a single vital sign or lab feature each time to train a logistic regression model, in which the journeys with missing values were not considered and the vital sign or lab feature required to have at least 10 number of valid values. We lastly exploited XGBoost to train a gradient boosted decision tree model using all the features without the imputation of missing values and also took into account the variability through the bootstrap sampling. We chose the 75-percentile of feature important scores obtained from the models with bootstrap sampling which was functioned as the important score for each feature.

### Learning algorithm

In light of the complex nonlinear interactions among the extracted features, we employed the gradient boosted tree models^33, 51^. These models are able to address the missing values inherently that are ubiquitous in the EMR and also the subsequent retrieved clinical features, such that we could avoid the basis/variance induced by imputing these missing values via the conventional approaches, e.g. mean, median, maximum and minimum etc. We also utilized LightGBM ^52^, a high-performance implementation of gradient boosted tree models, to fit our clinical models and then predict the corresponding targets, specifically, the binary classification in our PE prediction. We used the hyperparameter optimization package Hyperopt ^53^, on the basis of Bayesian optimization approaches, to automatically choose the optimal hyperparameters in the search space with the best performances on the designated metrics.

### Model interpretation and network

Interpretability becomes critical in the clinical settings to explain the specific impacts of each feature values on the predictive results not only in the granular level (the feature influences on each sample prediction) but also in the global model (the overall feature impacts on model outputs) ^33, 51, 54^. To accurately explain the contribution of each feature to the model output, we employed Shapley values realized by SHAP python package to obtain both the local and global interpretation. The Shapley values are attributed to the game theory, the only way that assigns the feature importance while maintaining two important foundations, the local accuracy and consistency. The Shapley values were successful to explain the machine learning model outputs in the clinical fields and capture the underlying clinical feature attributions and influences on the clinical predictions, e.g. chronic kidney disease ^51^. For the local explanation, the Shapley values allocate the model prediction for each sample into the single contributions of its associated feature values, thereby clearly interpreting the feature impacts in the granular level. By averaging across all the samples, Shapley values evaluate the overall contribution of each feature to the model output, thus achieved in the global level.

Generally, the outputs of the Shapley values using the TreeExplainer from SHAP package are log odds of the predicted values relative to the base value (the expected predictive value over the training dataset) which are additive. To draw the dependence plots, we transformed from the logit space into the probability space with the sigmoid function and calculated the relative risk (*RR*) score that is broadly used in the clinical fields ^55^. In the logit space, the Shapely values can be expressed as

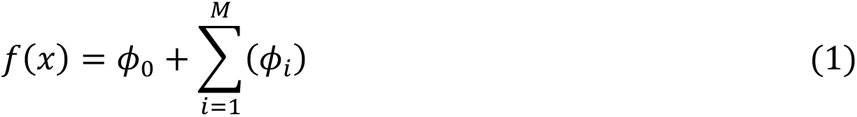

where *ϕ*_0_ is the base value representing the population prevalence; *ϕ_i_* is the Shapley value for each feature capturing the difference between the expected model output and output for the current prediction; *M* is the number of features. To display the dependence relationship for a single feature, we only computed the relative risk score through the Shapley value of that feature as follows

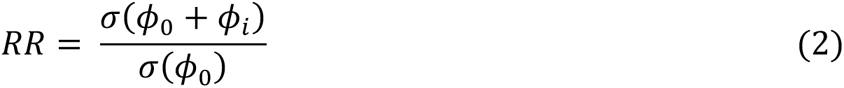

where *σ* is the sigmoid function. We could also aggregate certain related features into a higher level to investigate the corresponding overall feature effects on the model outputs. To this end, we only need to replace *ϕ_i_* with ∑*_i_*_∈*s*_ *ϕ_i_*, where *S* is the subset of features desired to be grouped.

Furthermore, we constructed networks connecting predictive features with respective PE (Methods in the Supplement).

### Current standard of care: ACOG criteria-based model

To evaluate PE risk in the clinical practice, we assessed the predictive model performance based on high risk factors and all risk factors (including high and moderate risk factors) recommended by ACOG^12^ (Methods in Supplement). We treated each risk factor as a binary feature and summed the binary features up to a risk score for every journey. We implemented the bootstrapping sampling to evaluate the standard deviation.

## Data Availability

The data used for this study are available from Mount Sinai Genomics Inc dba Sema4, but restrictions apply to the availability of these data, which were used under license for the current study, and so are not publicly available. Data are, however, available from the authors upon reasonable request and with the permission of Mount Sinai Genomics Inc dba Sema4.

## Acknowledgements

We thank the IT group in Sema4 and Mount Sinai Health System for database support.

## Author contributions

L.L., E.E.S., S.L. and Z.W. conceived and designed the study; S.L., Z.W., B.R., A.B.Z, E.S. performed data extraction from MSHS; S.L. and Z.W. performed statistical analysis and established machine learning models; S.L., Z.W., S.J.G, L.A.V. and L.L. contributed clinical interpretation; S.L., Z.W., L.A.V, A.B.Z., B.R., J.S., A.B.C, S.J.G, E.E.S. and L.L. wrote and edited the paper.

## Supplementary Material

### Methods

#### A. Patient population and pregnancy journey construction algorithm

We selected patients from Mount Sinai Hospital (MSH) and Mount Sinai West (MSW, we additionally added Mount Sinai Upper West, Mount Sinai St. Luke and Mount Sinai Beth Israel together) who are biologically female between age 12 and late 50 with either: (A) diagnosed with labor and delivery related International Classification of Disease 9^th^ or 10^th^ billing codes; (B) has vaginal or caesarean section delivery Current Procedural Terminology 4^th^ billing codes; or (C) admission records to labor and delivery facility. We identified 114,757 standalone delivery events for 88,907 unique patients, with 1.29 deliveries per patient.

We extracted gestational week mentioned in the admission records to labor and delivery facility, admit reason for inpatient and outpatient visits, and ICD9/10 diagnosis codes associated with specific gestational weeks. Then, we calculated the pregnancy date as *gestational week report date* - 7 * *gestational week*. We were able to find gestational week records and calculated the accurate pregnancy dates for 114,312 deliveries (83,954 unique patients), with the average age of 31.06 (std: 6.09) at pregnancy, the earliest delivery in 2002 and the latest delivery in 2019.

#### B. Data extraction, standardization and quality control

We extracted patient demographics, diagnoses, prescription drugs, anesthesia involved procedures, vital signs, and lab tests from MSDW EMR for patients in the study cohort (eTable 24). For each journey, we collected data from as early as 8 months before the pregnancy to as late as 10 weeks past the delivery. This (A) minimizes the influence of clinical signals associated with previous delivery yet preserves as much as possible prior-pregnancy information of the patient; and (B) corresponds to the timeline of preeclampsia development, which can happen as late as 10 weeks postpartum.

The demographic information includes age at the pregnancy, race, tobacco usage, alcohol usage, recent preeclampsia history, and Medicaid insurance. For patient who had reported multiple race groups, we assigned them to all race groups they had reported. We considered the patient under tobacco or alcohol usage, if they had reported such use during or before the 10 weeks after delivery.

The original diagnosis records in the MSHS EMR contains 14,688 ICD9/10 codes for the PD journey cohort. We grouped these ICD9/10 codes into 279 (of 285) Clinical Classification Software (CCS) single level categories ^1^ and 121 reproductive disease categories defined by our OB/GYN. This helps to reduce dimensionality of heterogenous diagnosis features to the granularity level suitable for building machine models and interpret clinical meaning.

We did not differentiate prescriptions of the same drug with difference dosage or under different brand names, and common ingredients of the different drugs may impact development of preeclampsia in the same way. Therefore, we mapped 8,682 unique prescribed drug names to 1,618 drug ingredients concepts registered in the RxNorm, using the RxNav API from NLM [https://rxnav.nlm.nih.gov/APIsOverview.html].

The PD journey cohort contains 718 unique CPT codes for anesthesia involved procedures, which were directly retrieved from the EMR.

We collected vital signs including pulse, systolic blood pressure, diastolic blood pressure, temperature, respirations, weight, height, O2 saturation, and pain scores for each journey and unified unit of measurements to Beats/Min, mm Hg, Fahrenheit degree, kilogram, centimeter, percentage, and 10-point scale respectively. For all but pain scores, we removed vital values beyond the range of Guinness World Records.

Patients in the PD journey cohort took a total of 603 lab tests. We normalized the lab names by mapping free text 603 lab names to 348 LOINC codes, using RELMA software [https://loinc.org/relma/] and manually validated the mapping results. We unified the unit of measurements for the same tests to the default unit of the corresponding LOINC. Out of all labs, 514 (283 LOINC) has numeric values, for which we unified the unit of measurements. For the 89 lab tests (65 LOINCs) that has descriptive text values, we unified nominal values and encoded nominal values to ordinal numbers based on test strip description ^2^ and color charts (e.g., “negative” -> 1, “trace” -> 2, “small” -> 3, “moderate”-> 4, “large” -> 5).

#### C. Current standard of care: ACOG criteria-based model

We constructed the ACOG criteria-based model for antepartum using risk factors for PE recommended by ACOG^3^. We treated each risk factor as a binary feature and calculated a risk score for every pregnancy journey in the corresponding cohort by summing the risk factors. The risk factors are subdivided into high risk factors and moderate risk factors which are describe as follows^3^. We established ACOG criteria-based models based on high risk factor and all the risk factors (high and moderate risk factors), respectively.

High risk factors:

- History of preeclampsia, especially when accomplished by an adverse outcome
- Multifetal gestation
- Chronic hypertension
- Type 1 and 2 diabetes
- Renal disease
- Autoimmune disease (i.e. systemic lupus erythematosus, the antiphospholipid syndrome) Moderate risk factors:
- Nulliparity
- Obesity (body mass index greater than 30)
- Family history of preeclampsia (mother or sister)
- Sociodemographic characteristics (African American race, low socioeconomic status)
- Age 35 years or older
- Personal history factors (e.g., low birth weight or small for gestational age, previous adverse pregnancy outcome, more than 10-year pregnancy interval)

#### D. Network

We constructed networks connecting predictive features with respective PE stages including 17 gestational weeks during antepartum as well as intrapartum and postpartum. It is worth noting that, to better visually illustrate the important features and their associations with each pregnancy period in the network, we reduced the unique feature number from 68 to 30 for intrapartum and from 48 to 24 for postpartum, respectively, by adding features one by one based on the rank of SHAP importance until the prediction performance became flat (Supplementary Figure 1 and 2 show the feature sweeping where the features were derived from the unique features). The nodes in the network represent the stages of PE and identified predictive features. The edges in the network reflect two layers of information: feature importance and adjusted odds ratio. We applied grouped attribute layout in Cytoscape 3.7.2 ^4^ to draw the network, with node sizes proportional to their degrees, edge width proportional to the feature importance and edge color correspond to adjusted OR. Two networks are visualized: one with different time points across antepartum and one with aggregated antepartum models, together with intrapartum and postpartum models. For simplicity, features that are predictive to only one time point in antepartum are removed from the visualization.

---

**Supplementary Figure 1:**
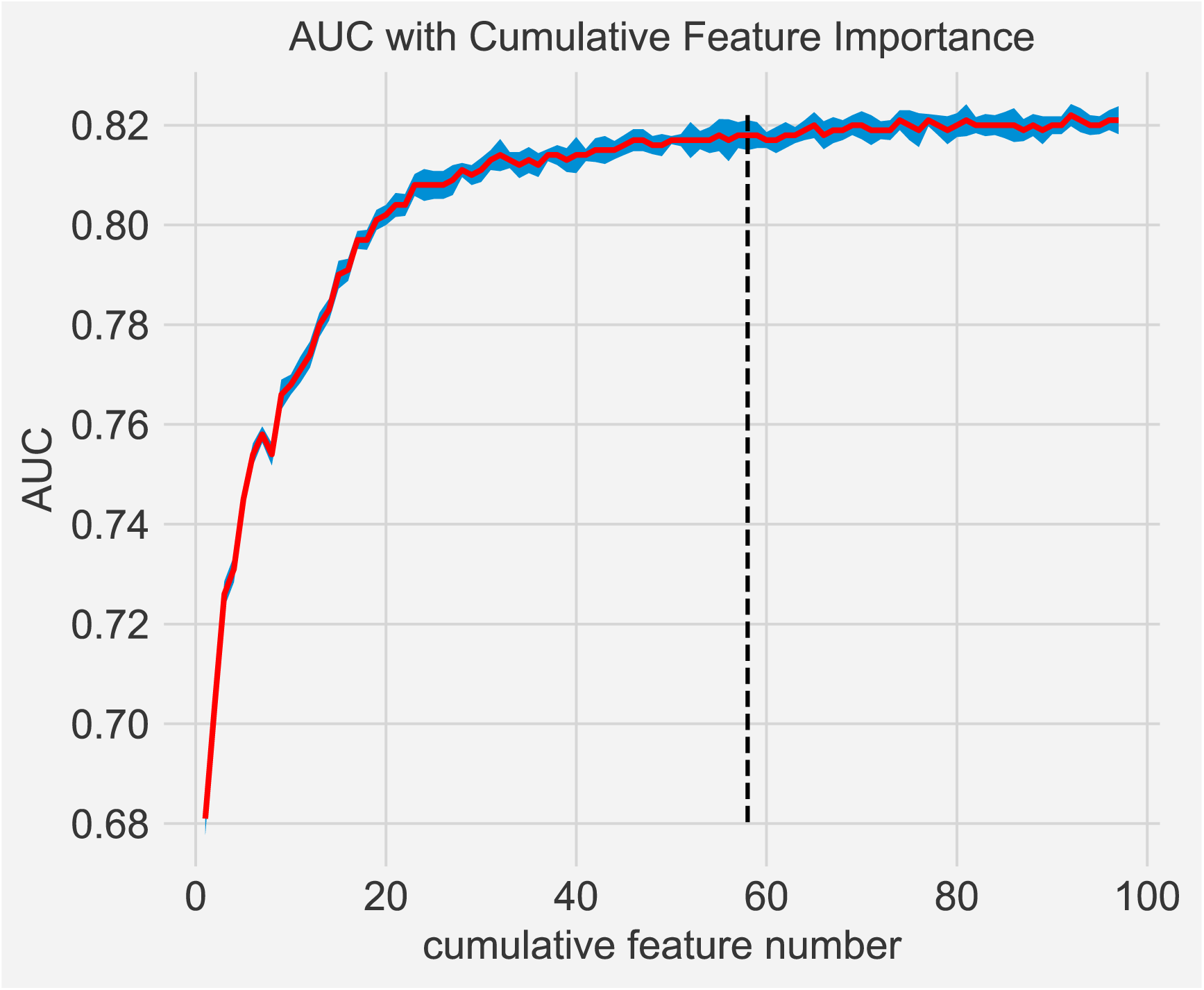
AUC score of features cumulation for the intrapartum

**Supplementary Figure 2:**
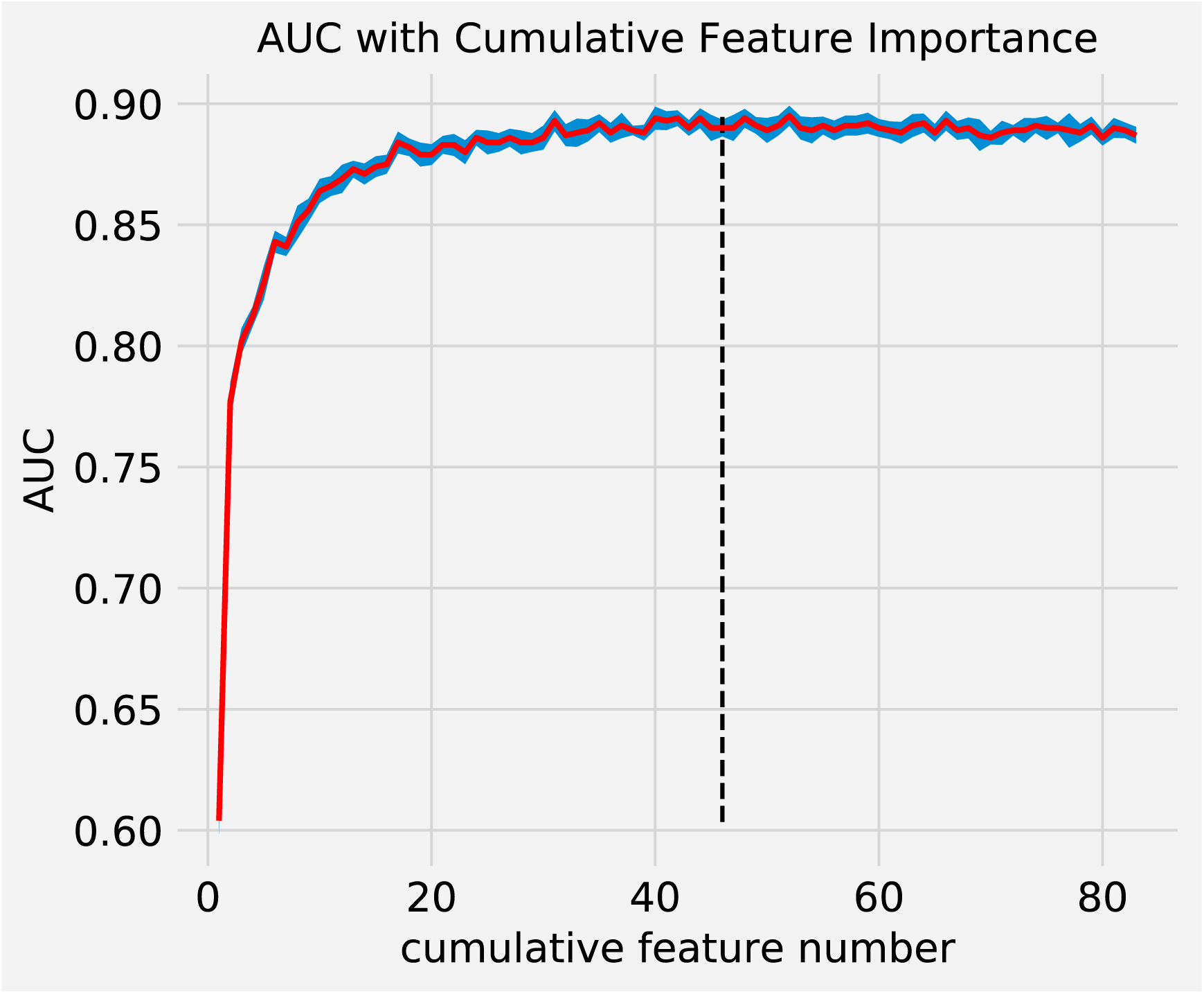
AUC score of features cumulation for the postpartum

**Supplementary Table 1:** Selected unique features for model week 4.

| Source feature | Feature importance | Estimate | Adjusted odds ratio | p-value | type | Known associations |
| --- | --- | --- | --- | --- | --- | --- |
| age_preg | 3.89 | 0.017295784 | 1.017446223 | 0.550880887 | demo | Y |
| SYSTOLIC BLOOD PRESSURE | 2.43 | 0.33475149 | 1.397593027 | 4.03249E-74 | vital | Y |
| DIASTOLIC BLOOD PRESSURE | 1.85 | 0.31477006 | 1.369944269 | 1.29053E-56 | vital | Y |
| African_American | 1.28 | 0.955756514 | 2.600637259 | 1.1267E-52 | demo | N |
| pe_hist | 1.17 | 2.45635387 | 11.66221198 | 3.7361E-154 | demo | Y |
| Caucasian | 1.15 | -0.686828704 | 0.50316924 | 2.59001E-31 | demo | N |
| medicaid | 1.03 | 0.640257955 | 1.896970148 | 3.24087E-28 | demo | N |
| Other female genital disorders | 1.02 | 0.021272725 | 1.021500603 | 6.40031E-06 | dx | N |
| WEIGHT | 0.94 | 0.185615074 | 1.203958737 | 8.10297E-23 | vital | Y |
| Hispanic | 0.82 | 0.412908536 | 1.511206799 | 1.08281E-09 | demo | N |
| Asian | 0.68 | -0.507282478 | 0.602129658 | 0.000111467 | demo | N |
| MEAN CORP. VOLUME | 0.61 | -0.084442121 | 0.919024846 | 1.41535E-05 | lab | N |
| Contraceptive and procreative management | 0.6 | 0.00690972 | 1.006933647 | 0.229379453 | dx | N |
| TEMPERATURE | 0.52 | -0.092326185 | 0.911807683 | 8.15368E-05 | vital | N |
| Propofol | 0.5 | -0.011373243 | 0.988691188 | 0.35733058 | rx | N |
| PULSE | 0.49 | 0.197880047 | 1.218816184 | 1.99776E-16 | vital | N |
| Headache; including migraine | 0.42 | 0.047285439 | 1.048421227 | 1.61654E-14 | dx | Y |
| Menstrual disorders | 0.38 | 0.004744737 | 1.004756011 | 0.436202365 | dx | N |
| RBC BLOOD CELL | 0.37 | 0.081440293 | 1.084848443 | 0.002882815 | lab | N |

**Supplementary Table 2:**
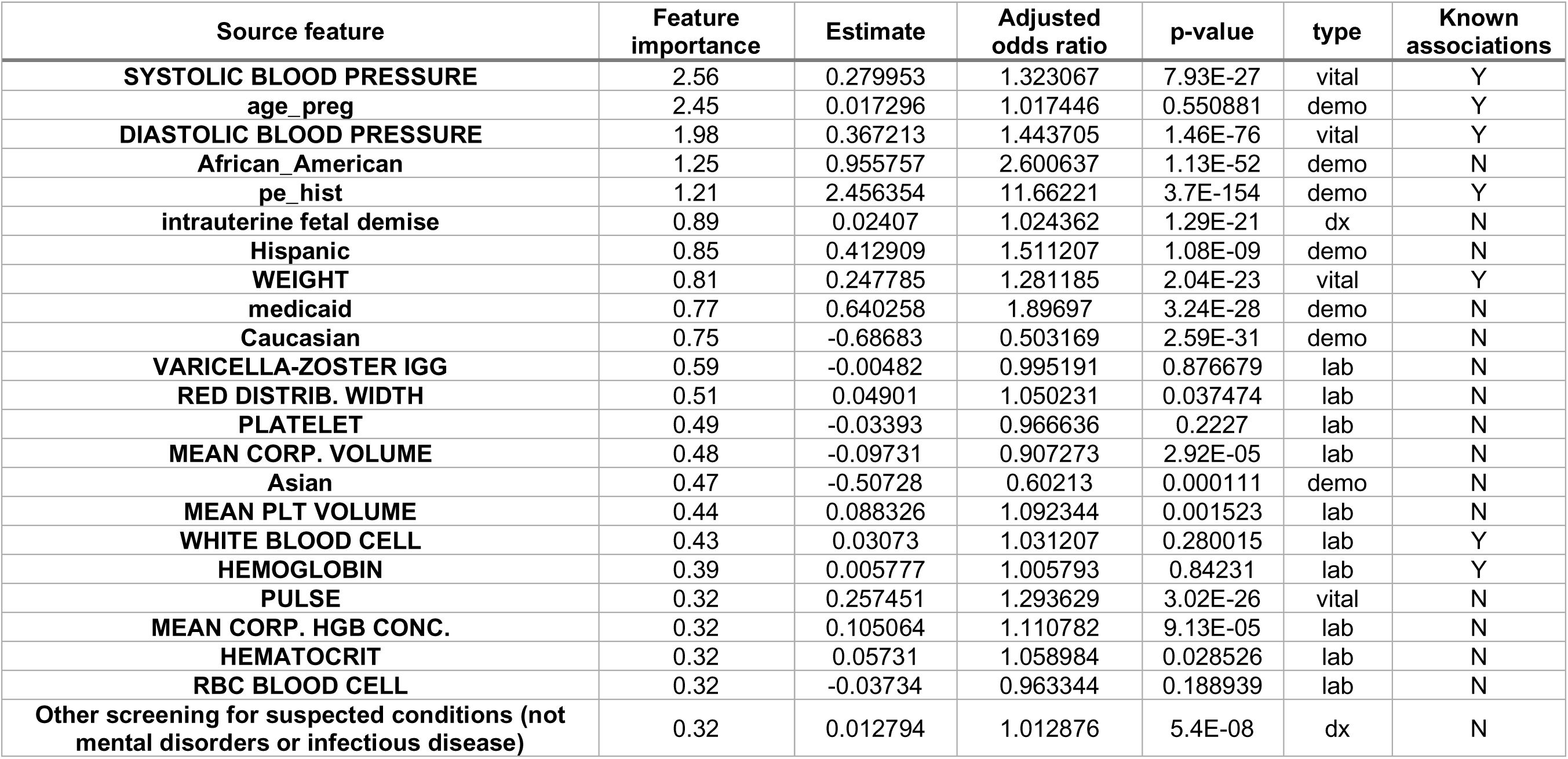
Selected unique features for model week 8.

| Source feature | Feature importance | Estimate | Adjusted odds ratio | p-value | type | Known associations |
| --- | --- | --- | --- | --- | --- | --- |
| **SYSTOLIC BLOOD PRESSURE** | 2.56 | 0.279953 | 1.323067 | 7.93E-27 | vital | Y |
| age_preg | 2.45 | 0.017296 | 1.017446 | 0.550881 | demo | Y |
| **DIASTOLIC BLOOD PRESSURE** | 1.98 | 0.367213 | 1.443705 | 1.46E-76 | vital | Y |
| African_American | 1.25 | 0.955757 | 2.600637 | 1.13E-52 | demo | N |
| pe_hist | 1.21 | 2.456354 | 11.66221 | 3.7E-154 | demo | Y |
| intrauterine fetal demise | 0.89 | 0.02407 | 1.024362 | 1.29E-21 | dx | N |
| Hispanic | 0.85 | 0.412909 | 1.511207 | 1.08E-09 | demo | N |
| **WEIGHT** | 0.81 | 0.247785 | 1.281185 | 2.04E-23 | vital | Y |
| medicaid | 0.77 | 0.640258 | 1.89697 | 3.24E-28 | demo | N |
| Caucasian | 0.75 | -0.68683 | 0.503169 | 2.59E-31 | demo | N |
| **VARICELLA-ZOSTER IGG** | 0.59 | -0.00482 | 0.995191 | 0.876679 | lab | N |
| **RED DISTRIB. WIDTH** | 0.51 | 0.04901 | 1.050231 | 0.037474 | lab | N |
| **PLATELET** | 0.49 | -0.03393 | 0.966636 | 0.2227 | lab | N |
| **MEAN CORP. VOLUME** | 0.48 | -0.09731 | 0.907273 | 2.92E-05 | lab | N |
| Asian | 0.47 | -0.50728 | 0.60213 | 0.000111 | demo | N |
| **MEAN PLT VOLUME** | 0.44 | 0.088326 | 1.092344 | 0.001523 | lab | N |
| **WHITE BLOOD CELL** | 0.43 | 0.03073 | 1.031207 | 0.280015 | lab | Y |
| **HEMOGLOBIN** | 0.39 | 0.005777 | 1.005793 | 0.84231 | lab | Y |
| **PULSE** | 0.32 | 0.257451 | 1.293629 | 3.02E-26 | vital | N |
| **MEAN CORP. HGB CONC.** | 0.32 | 0.105064 | 1.110782 | 9.13E-05 | lab | N |
| **HEMATOCRIT** | 0.32 | 0.05731 | 1.058984 | 0.028526 | lab | N |
| **RBC BLOOD CELL** | 0.32 | -0.03734 | 0.963344 | 0.188939 | lab | N |
| **Other screening for suspected conditions (not mental disorders or infectious disease)** | 0.32 | 0.012794 | 1.012876 | 5.4E-08 | dx | N |

**Supplementary Table 3:** Selected unique features for model week 12.

| Source feature | Feature importance | Estimate | Adjusted odds ratio | p-value | type | Known associations |
| --- | --- | --- | --- | --- | --- | --- |
| age_preg | 2.84 | 0.017303983 | 1.017455 | 0.550692 | demo | Y |
| pe_hist | 2.19 | 2.456335571 | 11.662 | 3.8E-154 | demo | Y |
| SYSTOLIC BLOOD PRESSURE | 2.09 | 0.322498107 | 1.380572 | 6.56E-32 | vital | Y |
| DIASTOLIC BLOOD PRESSURE | 1.82 | 0.418160206 | 1.519164 | 1.21E-93 | vital | Y |
| African_American | 1.58 | 0.955735195 | 2.600582 | 1.13E-52 | demo | N |
| Caucasian | 1.07 | -0.686797208 | 0.503185 | 2.61E-31 | demo | N |
| WEIGHT | 0.95 | 0.335163704 | 1.398169 | 2.91E-39 | vital | Y |
| Other screening for suspected conditions (not mental disorders or infectious disease) | 0.68 | 0.005166863 | 1.00518 | 0.00067 | dx | N |
| MEAN CORP. VOLUME | 0.66 | 0.051727137 | 1.053088 | 0.032007 | lab | N |
| higher order multiplee | 0.66 | 0.03192773 | 1.032443 | 9.01E-07 | dx | N |
| Hispanic | 0.65 | 0.412886639 | 1.511174 | 1.09E-09 | demo | N |
| RED DISTRIB. WIDTH | 0.64 | 0.101008717 | 1.106286 | 5.17E-09 | lab | N |
| MEAN CORP. HGB CONC. | 0.63 | -0.138477477 | 0.870683 | 1.06E-07 | lab | N |
| medicaid | 0.62 | 0.640315037 | 1.897078 | 3.21E-28 | demo | N |
| PH - DIPSTICK | 0.62 | 0.044755097 | 1.045772 | 0.115067 | lab | N |
| RBC BLOOD CELL | 0.6 | 0.067813999 | 1.070166 | 0.015482 | lab | N |
| MEAN CORP. HGB | 0.57 | -0.043569692 | 0.957366 | 0.121727 | lab | N |
| PLATELET | 0.53 | -0.07384261 | 0.928818 | 0.001703 | lab | N |
| Menstrual disorders | 0.53 | -0.002196103 | 0.997806 | 0.32397 | dx | N |
| LYMPHOCYTE % | 0.53 | 0.205215813 | 1.22779 | 2.76E-15 | lab | N |
| MEAN PLT VOLUME | 0.51 | 0.042397733 | 1.043309 | 0.129841 | lab | N |
| PULSE | 0.5 | 0.115064183 | 1.121945 | 8.21E-06 | vital | N |
| Asian | 0.49 | -0.50730215 | 0.602118 | 0.000111 | demo | N |
| Oxytocin | 0.47 | -0.009023532 | 0.991017 | 0.638098 | rx | N |
| WHITE BLOOD CELL | 0.47 | -0.065756363 | 0.936359 | 0.016343 | lab | Y |
| HEMOGLOBIN | 0.45 | 0.024006407 | 1.024297 | 0.405191 | lab | Y |
| Other pregnancy and delivery including normal | 0.42 | 0.010441012 | 1.010496 | 2.82E-13 | dx | N |
| VARICELLA-ZOSTER IGG | 0.42 | -0.022978555 | 0.977283 | 0.537668 | lab | N |
| HEMATOCRIT | 0.4 | 0.292375893 | 1.339606 | 7.48E-27 | lab | N |
| QTC | 0.35 | 0.211634956 | 1.235697 | 1.2E-41 | lab | N |

**Supplementary Table 4:** Selected unique features for model week 16.

| Source feature | Feature importance | Estimate | Adjusted odds ratio | p-value | type | Known associations |
| --- | --- | --- | --- | --- | --- | --- |
| **SYSTOLIC BLOOD PRESSURE** | 2.96 | 0.37141 | 1.449777 | 1.07E-39 | vital | Y |
| age_preg | 2.8 | 0.017258 | 1.017408 | 0.551748 | demo | Y |
| pe_hist | 2.79 | 2.456262 | 11.66114 | 3.8E-154 | demo | Y |
| **DIASTOLIC BLOOD PRESSURE** | 2.22 | 0.446819 | 1.563331 | 4.6E-106 | vital | Y |
| African_American | 1.75 | 0.956069 | 2.60145 | 1.04E-52 | demo | N |
| twin_pregnancy | 1.19 | 0.022797 | 1.023059 | 3.39E-09 | dx | N |
| **WEIGHT** | 1.11 | 0.221214 | 1.247591 | 8.21E-30 | vital | Y |
| Caucasian | 1 | -0.68697 | 0.5031 | 2.52E-31 | demo | N |
| Hispanic | 0.83 | 0.413049 | 1.511419 | 1.07E-09 | demo | N |
| **Other screening for suspected conditions (not mental disorders or infectious disease)** | 0.8 | 0.004822 | 1.004834 | 0.000388 | dx | N |
| **PULSE** | 0.77 | -0.03132 | 0.969161 | 0.275455 | vital | N |
| **RED DISTRIB. WIDTH** | 0.75 | -0.07223 | 0.930316 | 0.004888 | lab | N |
| Headache; including migraine | 0.7 | 0.031587 | 1.032091 | 4.35E-26 | dx | Y |
| **U-PROTEIN** | 0.68 | 0.303927 | 1.355171 | 3.71E-72 | lab | Y |
| **MEAN CORP. VOLUME** | 0.67 | -0.14404 | 0.86585 | 1.18E-12 | lab | N |
| **PLATELET** | 0.66 | 0.034389 | 1.034987 | 0.222239 | lab | N |
| **MEAN CORP. HGB** | 0.66 | -0.12481 | 0.882668 | 2.9E-07 | lab | N |
| gestational hypertension | 0.63 | 0.064516 | 1.066643 | 9.82E-87 | dx | Y |
| **LYMPHOCYTE %** | 0.61 | 0.209727 | 1.233342 | 1.59E-15 | lab | N |
| **WHITE BLOOD CELL** | 0.59 | 0.0764 | 1.079394 | 0.000328 | lab | Y |
| medicaid | 0.54 | 0.640293 | 1.897036 | 3.22E-28 | demo | N |
| **PH - DIPSTICK** | 0.53 | 0.080683 | 1.084027 | 0.003703 | lab | N |
| **HEMOGLOBIN** | 0.53 | 0.043851 | 1.044827 | 0.121799 | lab | Y |
| **HEMATOCRIT** | 0.52 | -0.0674 | 0.934819 | 0.008571 | lab | N |
| Other female genital disorders | 0.52 | 0.012553 | 1.012632 | 4.02E-10 | dx | N |
| **MEAN CORP. HGB CONC.** | 0.51 | -0.08618 | 0.917432 | 0.001618 | lab | N |
| Oxytocin | 0.48 | -0.00914 | 0.990898 | 0.632377 | rx | N |
| **MEAN PLT VOLUME** | 0.47 | 0.103712 | 1.109281 | 4.5E-06 | lab | N |
| Residual codes; unclassified | 0.45 | 0.010302 | 1.010355 | 2.24E-09 | dx | N |
| **RESPIRATIONS** | 0.41 | -0.04347 | 0.957462 | 0.076876 | vital | N |
| **RBC BLOOD CELL** | 0.4 | -0.09872 | 0.905994 | 0.000207 | lab | N |
| **MONOCYTE %** | 0.39 | 0.239263 | 1.270313 | 3.25E-21 | lab | N |
| **URIC ACID-BLD** | 0.39 | 0.246033 | 1.278941 | 5.12E-96 | lab | N |
| **VARICELLA-ZOSTER IGG** | 0.38 | -0.02732 | 0.973052 | 0.480327 | lab | N |
| **CREATININE-SERUM** | 0.38 | 0.019701 | 1.019896 | 0.032606 | lab | N |
| **LDH-BLD** | 0.36 | 0.248244 | 1.281773 | 2.01E-76 | lab | N |

**Supplementary Table 5:**
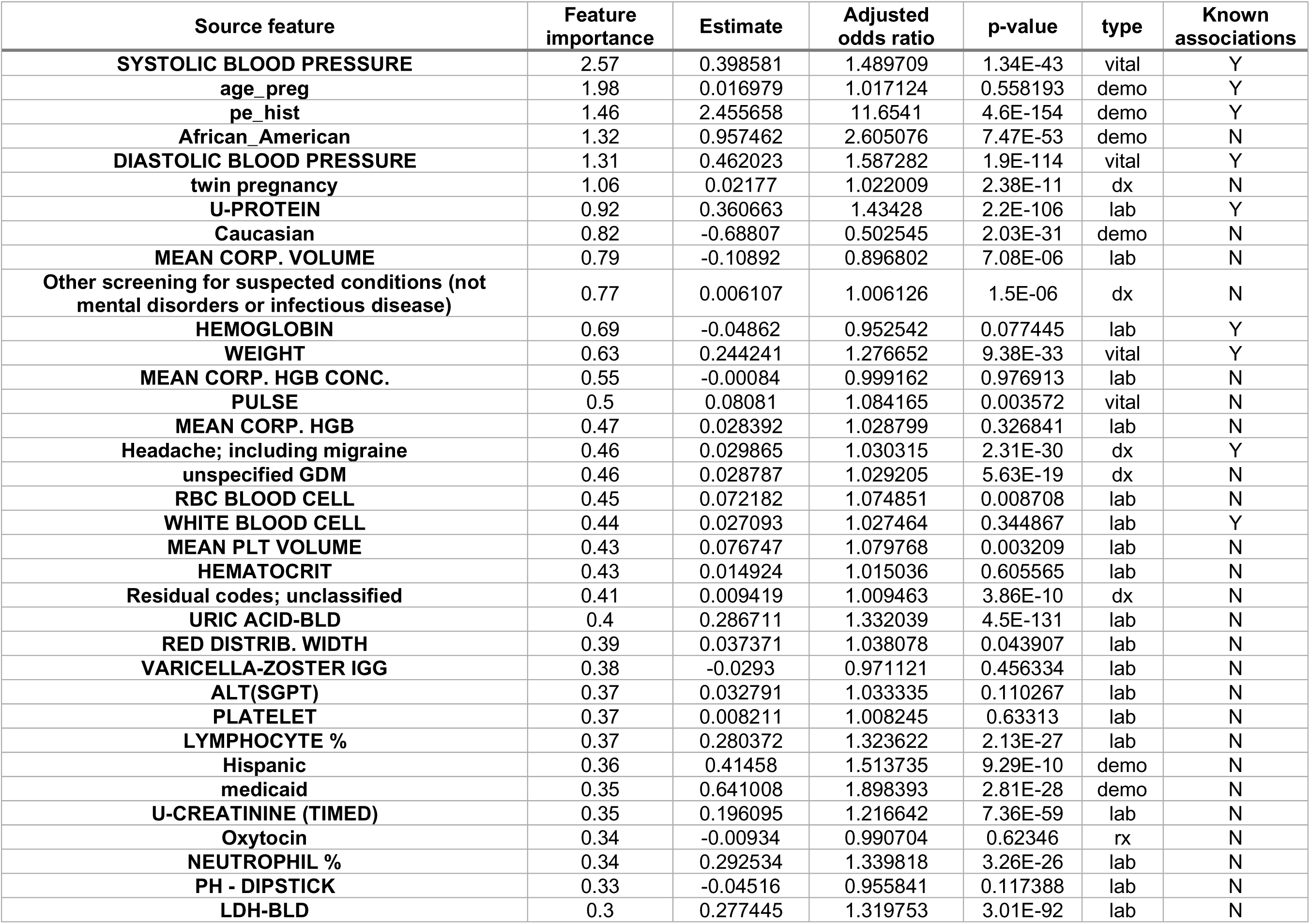
Selected unique features for model week 20.

**Supplementary Table 6:** Selected unique features for model week 22.

| Source feature | Feature importance | Estimate | Adjusted odds ratio | p-value | type | Known associations |
| --- | --- | --- | --- | --- | --- | --- |
| age_preg | 4.39 | 0.016656 | 1.016795 | 0.565702 | demo | Y |
| SYSTOLIC BLOOD PRESSURE | 3.51 | 0.416524 | 1.516681 | 2.48E-46 | vital | Y |
| pe_hist | 3.5 | 2.454486 | 11.64045 | 6.4E-154 | demo | Y |
| DIASTOLIC BLOOD PRESSURE | 2.78 | 0.486965 | 1.627369 | 2.4E-127 | vital | Y |
| African_American | 2.59 | 0.960299 | 2.612478 | 3.77E-53 | demo | N |
| twin pregnancy | 2.1 | 0.021054 | 1.021277 | 4.23E-12 | dx | N |
| RBC BLOOD CELL | 1.69 | 0.039529 | 1.040321 | 0.170245 | lab | N |
| Other screening for suspected conditions (not mental disorders or infectious disease) | 1.63 | 0.00736 | 1.007388 | 2.96E-09 | dx | N |
| U-PROTEIN | 1.59 | 0.382911 | 1.466547 | 1.2E-120 | lab | Y |
| WEIGHT | 1.47 | 0.234509 | 1.264288 | 1.14E-29 | vital | Y |
| RED DISTRIB. WIDTH | 1.18 | 0.107171 | 1.113124 | 3.27E-08 | lab | N |
| MEAN CORP. VOLUME | 1.16 | -0.06277 | 0.939157 | 0.01369 | lab | N |
| Caucasian | 1.16 | -0.68975 | 0.5017 | 1.45E-31 | demo | N |
| gestational hypertension | 1.1 | 0.059175 | 1.060961 | 1.4E-126 | dx | Y |
| MEAN PLT VOLUME | 1.09 | -0.00975 | 0.990299 | 0.736049 | lab | N |
| MEAN CORP. HGB CONC. | 1.08 | -0.05207 | 0.949263 | 0.065408 | lab | N |
| LYMPHOCYTE % | 0.98 | 0.28627 | 1.331452 | 2.2E-28 | lab | N |
| Oxytocin | 0.98 | -0.00988 | 0.990171 | 0.600301 | rx | N |
| unspecified GDM | 0.96 | 0.026182 | 1.026528 | 6.24E-18 | dx | N |
| intrauterine fetal demise | 0.91 | 0.018371 | 1.018541 | 4.52E-45 | dx | N |
| WHITE BLOOD CELL | 0.9 | -0.01607 | 0.984055 | 0.585434 | lab | Y |
| O2 SATURATION | 0.89 | -0.02102 | 0.9792 | 0.60592 | vital | N |
| PH - DIPSTICK | 0.87 | -0.11351 | 0.892693 | 5.21E-05 | lab | N |
| advanced maternal age | 0.82 | 0.003775 | 1.003782 | 0.025884 | dx | Y |
| NEUTROPHIL # | 0.82 | 0.273371 | 1.314388 | 3.94E-27 | lab | N |
| PULSE | 0.79 | 0.190048 | 1.209308 | 1.63E-14 | vital | N |
| Headache; including migraine | 0.78 | 0.029206 | 1.029637 | 1.34E-31 | dx | Y |
| VARICELLA-ZOSTER IGG | 0.77 | -0.02881 | 0.9716 | 0.461866 | lab | N |
| ALT(SGPT) | 0.77 | 0.062956 | 1.06498 | 0.039941 | lab | N |
| MEAN CORP. HGB | 0.75 | -0.11052 | 0.895364 | 2.8E-06 | lab | N |
| LDH-BLD | 0.74 | 0.291921 | 1.338997 | 4.54E-97 | lab | N |
| MONOCYTE % | 0.74 | 0.275806 | 1.317592 | 9.58E-28 | lab | N |
| HEMATOCRIT | 0.74 | 0.045964 | 1.047037 | 0.099382 | lab | N |
| SPEC GRAVITY-DIPSTICK | 0.73 | 0.009235 | 1.009278 | 0.909824 | lab | N |
| Other pregnancy and delivery including normal | 0.69 | 0.010137 | 1.010189 | 5.23E-16 | dx | N |
| HEMOGLOBIN | 0.68 | 0.007741 | 1.007771 | 0.788199 | lab | Y |
| medicaid | 0.67 | 0.641573 | 1.899467 | 2.53E-28 | demo | N |
| Residual codes; unclassified | 0.64 | 0.009026 | 1.009067 | 7.82E-11 | dx | N |
| POTASSIUMBLD | 0.64 | 0.272633 | 1.313418 | 1.37E-44 | lab | N |
| HEIGHT | 0.63 | 0.294155 | 1.341992 | 1.52E-26 | vital | N |

**Supplementary Table 7:** Selected unique features for model week 24.

| Source feature | Feature importance | Estimate | Adjusted odds ratio | p-value | type | Known associations |
| --- | --- | --- | --- | --- | --- | --- |
| age_preg | 4.38 | 0.017884 | 1.018045 | 0.537622 | demo | Y |
| pe_hist | 3.77 | 2.453614 | 11.6303 | 8.5E-154 | demo | Y |
| SYSTOLIC BLOOD PRESSURE | 3.72 | 0.583687 | 1.792637 | 3.3E-215 | vital | Y |
| DIASTOLIC BLOOD PRESSURE | 3.34 | 0.514945 | 1.673547 | 3.2E-141 | vital | Y |
| African_American | 2.61 | 0.960672 | 2.613452 | 4.27E-53 | demo | N |
| twin pregnancy | 2.37 | 0.021319 | 1.021548 | 8.74E-14 | dx | N |
| Other screening for suspected conditions (not mental disorders or infectious disease) | 2.26 | 0.008617 | 1.008654 | 2.73E-12 | dx | N |
| U-PROTEIN | 1.95 | 0.405699 | 1.500351 | 1.5E-135 | lab | Y |
| WEIGHT | 1.8 | 0.281771 | 1.325476 | 2.1E-56 | vital | Y |
| MEAN CORP. HGB | 1.76 | 0.068021 | 1.070388 | 0.014552 | lab | N |
| unspecified GDM | 1.72 | 0.025057 | 1.025374 | 7.48E-20 | dx | N |
| WHITE BLOOD CELL | 1.67 | 0.170879 | 1.186347 | 1.72E-13 | lab | Y |
| MEAN CORP. VOLUME | 1.4 | -0.09803 | 0.906626 | 2.23E-05 | lab | N |
| FETAL FIBRONECTIN | 1.38 | 0.083655 | 1.087254 | 0.0001 | lab | N |
| gestational hypertension | 1.35 | 0.058596 | 1.060346 | 4E-142 | dx | Y |
| intrauterine fetal demise | 1.22 | 0.017982 | 1.018145 | 7.08E-46 | dx | N |
| RBC BLOOD CELL | 1.17 | 0.031786 | 1.032296 | 0.266075 | lab | N |
| Caucasian | 1.15 | -0.69007 | 0.501541 | 1.42E-31 | demo | N |
| Headache; including migraine | 1.11 | 0.029542 | 1.029983 | 3.23E-36 | dx | Y |
| MONOCYTE % | 1.06 | 0.259413 | 1.296169 | 1.45E-22 | lab | N |
| RED DISTRIB. WIDTH | 1 | 0.04111 | 1.041967 | 0.1122 | lab | N |
| HEMATOCRIT | 0.97 | 0.017045 | 1.017191 | 0.556662 | lab | N |
| HEMOGLOBIN | 0.97 | 0.047942 | 1.04911 | 0.100017 | lab | Y |
| LDH-BLD | 0.97 | 0.304929 | 1.356528 | 1.9E-104 | lab | N |
| Oxytocin | 0.97 | 0.017167 | 1.017315 | 0.191914 | rx | N |
| MEAN PLT VOLUME | 0.96 | 0.054641 | 1.056162 | 0.054351 | lab | N |
| ALT(SGPT) | 0.94 | 0.085953 | 1.089755 | 0.000419 | lab | N |
| MEAN CORP. HGB CONC. | 0.9 | -0.08213 | 0.921149 | 0.003598 | lab | N |
| LYMPHOCYTE % | 0.88 | 0.2976 | 1.346623 | 1.8E-30 | lab | N |
| RESPIRATIONS | 0.87 | 0.041982 | 1.042876 | 0.028083 | vital | N |
| PH - DIPSTICK | 0.84 | -0.01745 | 0.9827 | 0.545635 | lab | N |
| PULSE | 0.77 | 0.095485 | 1.100193 | 0.000594 | vital | N |
| VARICELLA-ZOSTER IGG | 0.73 | -0.02805 | 0.972343 | 0.470532 | lab | N |
| nonreassuring fetal status | 0.71 | 0.00258 | 1.002583 | 0.038058 | dx | N |
| POTASSIUMBLD | 0.7 | 0.285293 | 1.330151 | 1.58E-48 | lab | N |
| O2 SATURATION | 0.7 | -0.02916 | 0.971262 | 0.496507 | vital | N |
| Contraceptive and procreative management | 0.7 | 0.005034 | 1.005046 | 0.020523 | dx | N |
| NEUTROPHIL # | 0.65 | 0.2875 | 1.333091 | 8.19E-30 | lab | N |

**Supplementary Table 8:** Selected unique features for model week 26.

| Source feature | Feature importance | Estimate | Adjusted odds ratio | p-value | type | Known associations |
| --- | --- | --- | --- | --- | --- | --- |
| **SYSTOLIC BLOOD PRESSURE** | 3.6 | 0.456552 | 1.578622 | 5.44E-52 | vital | Y |
| age_preg | 2.76 | 0.015092 | 1.015207 | 0.604166 | demo | Y |
| **DIASTOLIC BLOOD PRESSURE** | 2.32 | 0.533845 | 1.705478 | 1.8E-151 | vital | Y |
| pe_hist | 2.23 | 2.449529 | 11.58289 | 1E-151 | demo | Y |
| African_American | 1.61 | 0.964382 | 2.623166 | 4.17E-53 | demo | N |
| twin pregnancy | 1.38 | 0.020733 | 1.020949 | 6.93E-14 | dx | N |
| **U-PROTEIN** | 1.32 | 0.42952 | 1.53652 | 1.9E-150 | lab | Y |
| **WEIGHT** | 1.15 | -0.12492 | 0.88257 | 5.25E-09 | vital | Y |
| gestational hypertension | 1.08 | 0.057529 | 1.059216 | 9.2E-170 | dx | Y |
| **Other screening for suspected conditions (not mental disorders or infectious disease)** | 1.02 | 0.008511 | 1.008547 | 4.58E-12 | dx | N |
| **FETAL FIBRONECTIN** | 0.94 | 0.116482 | 1.123538 | 3.56E-08 | lab | N |
| **MEAN CORP. VOLUME** | 0.91 | -0.05542 | 0.946092 | 0.030968 | lab | N |
| Oxytocin | 0.85 | -0.01024 | 0.989808 | 0.577733 | rx | N |
| Caucasian | 0.85 | -0.69023 | 0.501463 | 2.21E-31 | demo | N |
| **URIC ACID-BLD** | 0.81 | 0.349676 | 1.418607 | 2.1E-189 | lab | N |
| Headache; including migraine | 0.81 | 0.029708 | 1.030154 | 2.37E-40 | dx | Y |
| **MEAN CORP. HGB** | 0.78 | -0.10406 | 0.901175 | 3.12E-05 | lab | N |
| **MEAN PLT VOLUME** | 0.76 | 0.065406 | 1.067592 | 0.016785 | lab | N |
| **HEMATOCRIT** | 0.74 | 0.004679 | 1.00469 | 0.871946 | lab | N |
| unspecified GDM | 0.68 | 0.019413 | 1.019602 | 1.59E-16 | dx | N |
| **RBC BLOOD CELL** | 0.65 | 0.004744 | 1.004755 | 0.870551 | lab | N |
| **LDH-BLD** | 0.62 | 0.328358 | 1.388686 | 1.6E-120 | lab | N |
| **MEAN CORP. HGB CONC.** | 0.61 | -0.08447 | 0.919002 | 0.002656 | lab | N |
| **MONOCYTE %** | 0.6 | 0.254296 | 1.289554 | 3.9E-21 | lab | N |
| **PLATELET** | 0.59 | -0.01539 | 0.984729 | 0.850649 | lab | N |
| **HEMOGLOBIN** | 0.58 | 0.036373 | 1.037043 | 0.214696 | lab | Y |
| **WHITE BLOOD CELL** | 0.54 | 0.000882 | 1.000882 | 0.975803 | lab | Y |
| **PULSE** | 0.53 | -0.1237 | 0.883647 | 8.1E-07 | vital | N |
| **ALT(SGPT)** | 0.52 | 0.178881 | 1.195878 | 4.15E-24 | lab | N |
| **RED DISTRIB. WIDTH** | 0.5 | 0.044961 | 1.045987 | 0.080831 | lab | N |
| **NEUTROPHIL %** | 0.47 | 0.329653 | 1.390485 | 1.12E-30 | lab | N |
| medicaid | 0.46 | 0.646144 | 1.908169 | 1.73E-28 | demo | N |
| Abdominal pain | 0.44 | 0.014128 | 1.014228 | 2.19E-16 | dx | N |
| **O2 SATURATION** | 0.44 | -0.00044 | 0.999565 | 0.987891 | vital | N |
| **CREATININE-SERUM** | 0.42 | 0.016216 | 1.016348 | 0.056421 | lab | N |
| intrauterine fetal demise | 0.42 | 0.017049 | 1.017195 | 1.88E-43 | dx | N |

**Supplementary Table 9:**
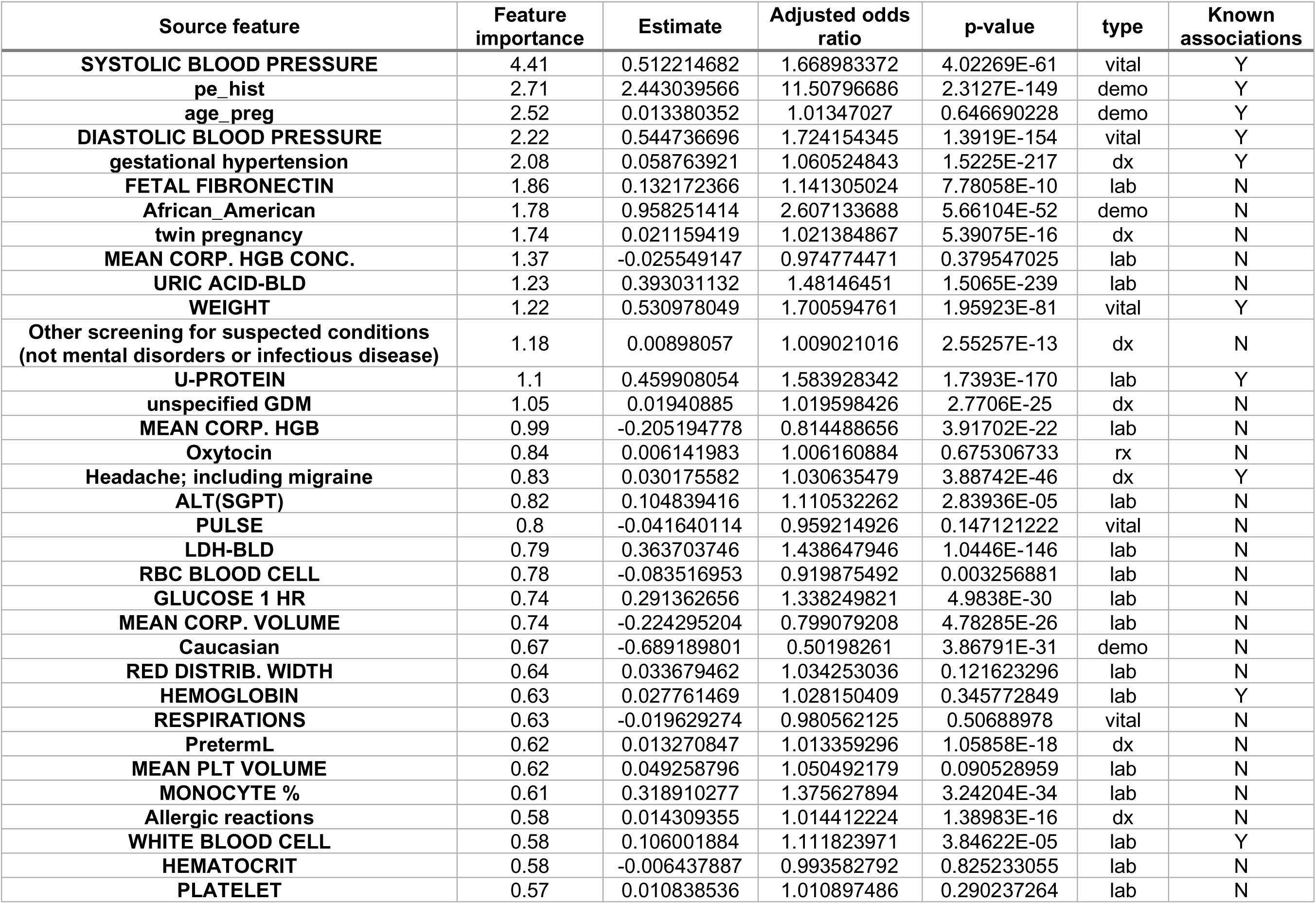
Selected unique features for model week 28.

**Supplementary Table 10.** : Selected unique features for model week 30.

| Source feature | Feature importance | Estimate | Adjusted odds ratio | p-value | type | Known associations |
| --- | --- | --- | --- | --- | --- | --- |
| **SYSTOLIC BLOOD PRESSURE** | 3.58 | 0.538657 | 1.713704 | 1.1E-63 | vital | Y |
| pe_hist | 2.8 | 2.437368 | 11.44288 | 2.5E-146 | demo | Y |
| age_preg | 2.25 | 0.016709 | 1.016849 | 0.570043 | demo | Y |
| twin pregnancy | 1.8 | 0.020292 | 1.020499 | 6.86E-16 | dx | N |
| gestational hypertension | 1.8 | 0.059017 | 1.060793 | 3.3E-273 | dx | Y |
| **FETAL FIBRONECTIN** | 1.67 | 0.155409 | 1.168135 | 6.81E-13 | lab | N |
| **DIASTOLIC BLOOD PRESSURE** | 1.59 | 0.545168 | 1.724898 | 5.5E-152 | vital | Y |
| African_American | 1.54 | 0.95883 | 2.608642 | 3.32E-51 | demo | N |
| **URIC ACID-BLD** | 1.39 | 0.428149 | 1.534415 | 3.1E-274 | lab | N |
| **Other screening for suspected conditions (not mental disorders or infectious disease)** | 1.18 | 0.009458 | 1.009503 | 1.53E-14 | dx | N |
| **MEAN CORP. VOLUME** | 0.99 | -0.22836 | 0.795839 | 2.53E-26 | lab | N |
| Headache; including migraine | 0.98 | 0.030143 | 1.030602 | 1.58E-50 | dx | Y |
| **WEIGHT** | 0.96 | 0.239602 | 1.270744 | 1.83E-27 | vital | Y |
| **U-PROTEIN** | 0.89 | 0.495144 | 1.640735 | 3.5E-194 | lab | Y |
| **RED DISTRIB. WIDTH** | 0.79 | 0.059003 | 1.060778 | 0.004377 | lab | N |
| **MEAN CORP. HGB CONC.** | 0.74 | -0.08015 | 0.922978 | 0.004978 | lab | N |
| Oxytocin | 0.73 | 0.005843 | 1.00586 | 0.680944 | rx | N |
| **PLATELET** | 0.73 | 0.01577 | 1.015895 | 0.177961 | lab | N |
| Caucasian | 0.7 | -0.68421 | 0.50449 | 2.5E-30 | demo | N |
| intrauterine fetal demise | 0.69 | 0.018431 | 1.018602 | 1.97E-57 | dx | N |
| **MEAN CORP. HGB** | 0.64 | -0.23514 | 0.790458 | 1.12E-24 | lab | N |
| **Other pregnancy and delivery including normal** | 0.62 | 0.004259 | 1.004268 | 0.000314 | dx | N |
| **PULSE** | 0.6 | -0.07689 | 0.925994 | 0.005543 | vital | N |
| **MEAN PLT VOLUME** | 0.6 | 0.036115 | 1.036775 | 0.216423 | lab | N |
| **HEMATOCRIT** | 0.59 | -0.01725 | 0.982903 | 0.557052 | lab | N |
| **RBC BLOOD CELL** | 0.58 | 0.175926 | 1.19235 | 1.18E-10 | lab | N |
| **WHITE BLOOD CELL** | 0.57 | 0.10515 | 1.110877 | 7.63E-05 | lab | Y |
| **O2 SATURATION** | 0.53 | -0.03036 | 0.970099 | 0.668428 | vital | N |
| **LYMPHOCYTE %** | 0.52 | 0.342798 | 1.408884 | 3.11E-37 | lab | N |
| **LDH-BLD** | 0.5 | 0.399441 | 1.490992 | 1.4E-173 | lab | N |
| **FIBRINOGEN** | 0.5 | 0.45427 | 1.575024 | 4.6E-294 | lab | N |
| **ALT(SGPT)** | 0.5 | 0.106334 | 1.112193 | 1.84E-05 | lab | N |
| **HEMOGLOBIN** | 0.49 | 0.035909 | 1.036562 | 0.219787 | lab | Y |
| **ALK PHOSPHATASE, BLD** | 0.48 | 0.292409 | 1.339651 | 2.99E-62 | lab | N |

**Supplementary Table 11.** : Selected unique features for model week 32.

| Source feature | Feature importance | Estimate | Adjusted odds ratio | p-value | type | Known associations |
| --- | --- | --- | --- | --- | --- | --- |
| **SYSTOLIC BLOOD PRESSURE** | 4.65 | 0.60619 | 1.833432 | 4.83E-72 | vital | Y |
| pe_hist | 2.62 | 2.427387 | 11.32924 | 3.2E-141 | demo | Y |
| gestational hypertension | 2.46 | 0.058112 | 1.059834 | 0 | dx | Y |
| **DIASTOLIC BLOOD PRESSURE** | 1.89 | 0.642766 | 1.901734 | 2.36E-81 | vital | Y |
| age_preg | 1.89 | 0.007904 | 1.007936 | 0.790512 | demo | Y |
| twin pregnancy | 1.64 | 0.018471 | 1.018643 | 3.95E-13 | dx | N |
| **FETAL FIBRONECTIN** | 1.49 | 0.181901 | 1.199496 | 5.52E-17 | lab | N |
| African_American | 1.44 | 0.936165 | 2.550182 | 2.63E-47 | demo | N |
| **MEAN CORP. HGB** | 1.38 | -0.16166 | 0.850733 | 1.11E-12 | lab | N |
| Headache; including migraine | 1.18 | 0.031175 | 1.031666 | 6.64E-62 | dx | Y |
| **Other screening for suspected conditions (not mental disorders or infectious disease)** | 1.14 | 0.010179 | 1.010231 | 2.06E-16 | dx | N |
| **WEIGHT** | 1.11 | 0.33154 | 1.393112 | 4.11E-68 | vital | Y |
| **U-PROTEIN** | 0.88 | 0.561581 | 1.753443 | 1.2E-247 | lab | Y |
| unspecified GDM | 0.83 | 0.016284 | 1.016417 | 1.54E-25 | dx | N |
| **MEAN PLT VOLUME** | 0.82 | -0.07591 | 0.926902 | 0.010609 | lab | N |
| **FIBRINOGEN** | 0.81 | 0.527481 | 1.694658 | 0 | lab | N |
| **PULSE** | 0.81 | -0.05776 | 0.943876 | 0.042852 | vital | N |
| **URIC ACID-BLD** | 0.8 | 0.495093 | 1.640651 | 0 | lab | N |
| **MEAN CORP. HGB CONC.** | 0.79 | -0.14499 | 0.865031 | 2.3E-07 | lab | N |
| **O2 SATURATION** | 0.77 | -0.00084 | 0.999157 | 0.977392 | vital | N |
| **PLATELET** | 0.77 | 0.87738 | 2.404591 | 0.004874 | lab | N |
| **RED DISTRIB. WIDTH** | 0.76 | 0.056718 | 1.058357 | 0.003923 | lab | N |
| Essential hypertension | 0.71 | 0.05644 | 1.058063 | 8.3E-147 | dx | Y |
| **MEAN CORP. VOLUME** | 0.67 | -0.13314 | 0.875345 | 9.54E-06 | lab | N |
| PretermL | 0.67 | 0.013712 | 1.013807 | 5.51E-29 | dx | N |
| **WHITE BLOOD CELL** | 0.66 | -0.01806 | 0.982104 | 0.534089 | lab | Y |
| **MONOCYTE %** | 0.56 | 0.34719 | 1.415086 | 3.25E-34 | lab | N |
| **HEMOGLOBIN** | 0.56 | -0.04404 | 0.956912 | 0.072439 | lab | Y |
| **RESPIRATIONS** | 0.55 | -0.01313 | 0.98696 | 0.67305 | vital | N |
| **LYMPHOCYTE %** | 0.55 | 0.347576 | 1.415631 | 9.78E-36 | lab | N |
| **HEMATOCRIT** | 0.55 | -0.04825 | 0.952894 | 0.092603 | lab | N |
| **PH - DIPSTICK** | 0.54 | -0.12553 | 0.882028 | 1.12E-05 | lab | N |
| **EOSINOPHIL %** | 0.54 | 0.154687 | 1.167293 | 2.86E-14 | lab | N |
| **ALK PHOSPHATASE, BLD** | 0.5 | 0.289199 | 1.335357 | 7.44E-62 | lab | N |

**Supplementary Table 12.** : Selected unique features for model week 34.

| Source feature | Feature importance | Estimate | Adjusted odds ratio | p-value | type | Known associations |
| --- | --- | --- | --- | --- | --- | --- |
| **SYSTOLIC BLOOD PRESSURE** | 5.3 | 0.653821 | 1.922875 | 1.43E-72 | vital | Y |
| gestational hypertension | 3.23 | 0.056492 | 1.058118 | 0 | dx | Y |
| pe_hist | 2.51 | 2.420706 | 11.2538 | 1.7E-131 | demo | Y |
| age_preg | 2.34 | 0.008315 | 1.008349 | 0.78699 | demo | Y |
| **FIBRINOGEN** | 1.92 | 0.588711 | 1.801665 | 0 | lab | N |
| **DIASTOLIC BLOOD PRESSURE** | 1.71 | 0.555188 | 1.742268 | 4.5E-141 | vital | Y |
| **WHITE BLOOD CELL** | 1.65 | 0.111964 | 1.118473 | 4.54E-05 | lab | Y |
| **Other pregnancy and delivery including normal** | 1.49 | -0.00156 | 0.99844 | 0.221939 | dx | N |
| **URIC ACID-BLD** | 1.44 | 0.548969 | 1.731467 | 0 | lab | N |
| **U-PROTEIN** | 1.34 | 0.609215 | 1.838987 | 2.1E-270 | lab | Y |
| large for GA | 1.33 | 0.010029 | 1.01008 | 4.09E-05 | dx | N |
| **Headache; including migraine** | 1.29 | 0.030646 | 1.03112 | 3.7E-61 | dx | Y |
| **WEIGHT** | 1.28 | 0.174777 | 1.190981 | 4.8E-15 | vital | Y |
| **MEAN CORP. HGB** | 1.24 | -0.15373 | 0.857506 | 1.51E-10 | lab | N |
| **HEMOGLOBIN** | 1.2 | -0.08934 | 0.914536 | 0.002092 | lab | Y |
| **MEAN CORP. VOLUME** | 1.05 | -0.08684 | 0.916828 | 0.000778 | lab | N |
| twin pregnancy | 1.03 | 0.020257 | 1.020463 | 1.27E-15 | dx | N |
| **HEMATOCRIT** | 1 | 0.010178 | 1.01023 | 0.740753 | lab | N |
| chronic hypertension | 0.94 | 0.056093 | 1.057696 | 1.5E-197 | dx | Y |
| African_American | 0.9 | 0.934435 | 2.545776 | 4.75E-44 | demo | N |
| **PLATELET** | 0.86 | 4.172745 | 64.89333 | 3.45E-09 | lab | Y |
| fibroids in pregnancy | 0.86 | 0.016304 | 1.016437 | 3.42E-08 | dx | N |
| **FETAL FIBRONECTIN** | 0.85 | 0.215031 | 1.239901 | 1.05E-22 | lab | N |
| **RED DISTRIB. WIDTH** | 0.85 | -0.00717 | 0.992859 | 0.812497 | lab | N |
| **RESPIRATIONS** | 0.79 | 0.025386 | 1.025711 | 0.307249 | vital | N |
| **Other screening for suspected conditions (not mental disorders or infectious disease)** | 0.77 | 0.00906 | 1.009101 | 8.12E-13 | dx | N |
| **MONOCYTE %** | 0.75 | 0.393142 | 1.481628 | 5.4E-45 | lab | N |
| **PULSE** | 0.73 | 0.1139 | 1.12064 | 4.98E-05 | vital | N |
| PretermL | 0.71 | 0.013535 | 1.013627 | 1.38E-30 | dx | N |
| **MEAN PLT VOLUME** | 0.67 | 0.142306 | 1.152929 | 3.63E-07 | lab | N |
| **GLUCOSE 1 HR** | 0.66 | 0.312527 | 1.366875 | 1.8E-28 | lab | N |
| **ALK PHOSPHATASE, BLD** | 0.61 | 0.283027 | 1.327141 | 4.29E-65 | lab | N |
| **MEAN CORP. HGB CONC.** | 0.58 | 0.023209 | 1.023481 | 0.447844 | lab | N |
| **HEIGHT** | 0.57 | 0.418862 | 1.52023 | 8.49E-41 | vital | N |
| **EOSINOPHIL %** | 0.56 | 0.15872 | 1.17201 | 2.09E-14 | lab | N |
| **U-CREATININE (TIMED)** | 0.55 | 0.332537 | 1.394501 | 2.3E-143 | lab | N |
| Essential hypertension | 0.53 | 0.054599 | 1.056117 | 9.5E-147 | dx | Y |
| **RBC BLOOD CELL** | 0.53 | -0.16819 | 0.84519 | 4.11E-09 | lab | N |

**Supplementary Table 13:** Selected unique features for model week 35.

| Source feature | Feature importance | Estimate | Adjusted odds ratio | p-value | type | Known associations |
| --- | --- | --- | --- | --- | --- | --- |
| **SYSTOLIC BLOOD PRESSURE** | 3.69 | 0.684297 | 1.982377 | 2.19E-72 | vital | Y |
| **gestational hypertension** | 2.11 | 0.054435 | 1.055944 | 0 | dx | Y |
| **pe_hist** | 1.77 | 2.44342 | 11.51234 | 4.3E-130 | demo | Y |
| **age_preg** | 1.51 | -0.00437 | 0.995637 | 0.889579 | demo | Y |
| **FIBRINOGEN** | 1.47 | 0.631588 | 1.880595 | 0 | lab | N |
| **DIASTOLIC BLOOD PRESSURE** | 1.38 | 0.577832 | 1.782171 | 2.3E-148 | vital | Y |
| **MEAN CORP. VOLUME** | 1.04 | -0.24674 | 0.781341 | 1.13E-24 | lab | N |
| **URIC ACID-BLD** | 1.02 | 0.580663 | 1.787224 | 0 | lab | N |
| **PLATELET** | 1.01 | -0.03824 | 0.962481 | 0.756975 | lab | N |
| **MEAN PLT VOLUME** | 0.99 | 0.014146 | 1.014246 | 0.65429 | lab | N |
| **U-PROTEIN** | 0.97 | 0.64151 | 1.899347 | 5.5E-284 | lab | Y |
| **African_American** | 0.91 | 0.93502 | 2.547264 | 4.96E-42 | demo | N |
| **chronic hypertension** | 0.85 | 0.054726 | 1.056251 | 4.5E-189 | dx | Y |
| **WEIGHT** | 0.8 | 0.125164 | 1.133334 | 1.93E-07 | vital | Y |
| **Headache; including migraine** | 0.78 | 0.03261 | 1.033147 | 8.31E-75 | dx | Y |
| **WHITE BLOOD CELL** | 0.73 | 0.026192 | 1.026538 | 0.394295 | lab | Y |
| **MEAN CORP. HGB** | 0.69 | -0.09856 | 0.906139 | 0.000864 | lab | N |
| **large for GA** | 0.65 | 0.010398 | 1.010452 | 1.26E-05 | dx | N |
| **fibroids in pregnancy** | 0.64 | 0.018256 | 1.018423 | 2.05E-10 | dx | N |
| **RED DISTRIB. WIDTH** | 0.64 | 0.072813 | 1.07553 | 0.00257 | lab | N |
| **Other ear and sense organ disorders** | 0.62 | 0.024606 | 1.024912 | 5.14E-08 | dx | N |
| **MEAN CORP. HGB CONC.** | 0.58 | -0.00154 | 0.998461 | 0.961018 | lab | N |
| **HEMATOCRIT** | 0.57 | -0.06992 | 0.932471 | 0.020119 | lab | N |
| **O2 SATURATION** | 0.54 | 0.003124 | 1.003129 | 0.898801 | vital | N |
| **PH - DIPSTICK** | 0.51 | -0.18519 | 0.830948 | 9.56E-10 | lab | N |
| **RBC BLOOD CELL** | 0.49 | -0.05373 | 0.947691 | 0.065065 | lab | N |
| **Other pregnancy and delivery including normal** | 0.49 | -0.00217 | 0.997834 | 0.110438 | dx | N |
| **in vitro fertilization** | 0.49 | 0.024409 | 1.024709 | 4.24E-13 | dx | N |
| **Caucasian** | 0.46 | -0.66319 | 0.515206 | 3.35E-25 | demo | N |
| **EOSINOPHIL %** | 0.44 | 0.163849 | 1.178036 | 5.79E-15 | lab | N |
| **HEMOGLOBIN** | 0.41 | 0.018711 | 1.018887 | 0.52783 | lab | Y |
| **obesity In pregnancy** | 0.39 | 0.030777 | 1.031256 | 8.41E-52 | dx | Y |
| **Residual codes; unclassified** | 0.39 | 0.010444 | 1.010499 | 1.54E-19 | dx | N |
| **LYMPHOCYTE %** | 0.38 | 0.416495 | 1.516636 | 4.2E-47 | lab | N |
| **RESPIRATIONS** | 0.38 | 0.006189 | 1.006209 | 0.833065 | vital | N |
| **PULSE** | 0.38 | 0.026296 | 1.026645 | 0.400374 | vital | N |
| **ALT(SGPT)** | 0.37 | 0.15643 | 1.169329 | 8.29E-10 | lab | N |
| **SPEC GRAVITY-DIPSTICK** | 0.35 | -0.02679 | 0.973568 | 0.892581 | lab | N |
| **MONOCYTE #** | 0.34 | 0.433823 | 1.543145 | 4.59E-53 | lab | N |

**Supplementary Table 14:** Selected unique features for model week 36.

| Source feature | Feature importance | Estimate | Adjusted odds ratio | p-value | type | Known associations |
| --- | --- | --- | --- | --- | --- | --- |
| **SYSTOLIC BLOOD PRESSURE** | 3.29 | 0.72831 | 2.071576 | 3.44E-71 | vital | Y |
| gestational hypertension | 2.34 | 0.053564 | 1.055024 | 0 | dx | Y |
| **U-PROTEIN** | 2.13 | 0.683811 | 1.981414 | 2.9E-298 | lab | Y |
| **DIASTOLIC BLOOD PRESSURE** | 2.11 | 0.544372 | 1.723526 | 1.2E-122 | vital | Y |
| pe_hist | 1.83 | 2.452551 | 11.61795 | 2.9E-121 | demo | Y |
| age_preg | 1.75 | -0.02762 | 0.972756 | 0.400237 | demo | Y |
| **FIBRINOGEN** | 1.7 | 0.671959 | 1.95807 | 0 | lab | N |
| chronic hypertension | 1.57 | 0.054727 | 1.056253 | 9.6E-191 | dx | Y |
| **Other pregnancy and delivery including normal** | 1.51 | -0.00483 | 0.995183 | 0.000855 | dx | N |
| **PULSE** | 1.42 | 0.239107 | 1.270114 | 5.27E-18 | vital | N |
| **URIC ACID-BLD** | 1.4 | 0.6104 | 1.841167 | 0 | lab | N |
| **WEIGHT** | 1.28 | 0.326035 | 1.385464 | 2.75E-54 | vital | Y |
| **PLATELET** | 0.91 | 7.674223 | 2152.151 | 2.12E-16 | lab | N |
| **Other ear and sense organ disorders** | 0.81 | 0.02456 | 1.024865 | 1.87E-07 | dx | N |
| **RESPIRATIONS** | 0.78 | -0.03537 | 0.965248 | 0.17836 | vital | N |
| **MEAN PLT VOLUME** | 0.76 | 0.676362 | 1.96671 | 5.88E-67 | lab | N |
| **EOSINOPHIL %** | 0.74 | 0.165792 | 1.180327 | 1.75E-14 | lab | N |
| **MEAN CORP. HGB** | 0.74 | -0.07098 | 0.931483 | 0.018216 | lab | N |
| Abdominal pain | 0.67 | 0.011784 | 1.011853 | 1.36E-12 | dx | N |
| Headache; including migraine | 0.67 | 0.031655 | 1.032161 | 4.8E-67 | dx | Y |
| Residual codes; unclassified | 0.65 | 0.010121 | 1.010172 | 2.22E-17 | dx | N |
| **O2 SATURATION** | 0.65 | -0.00594 | 0.994077 | 0.878103 | vital | N |
| **WHITE BLOOD CELL** | 0.65 | 0.134475 | 1.143936 | 8.68E-06 | lab | Y |
| **TEMPERATURE** | 0.64 | -0.03578 | 0.964851 | 0.249146 | vital | N |
| **RED DISTRIB. WIDTH** | 0.62 | -0.03069 | 0.969772 | 0.33706 | lab | N |
| **HEMATOCRIT** | 0.62 | 0.038145 | 1.038882 | 0.243547 | lab | N |
| **MEAN CORP. VOLUME** | 0.62 | 0.642798 | 1.901794 | 7.13E-58 | lab | N |
| **RBC BLOOD CELL** | 0.6 | -0.04311 | 0.957802 | 0.188481 | lab | N |
| **ALK PHOSPHATASE, BLD** | 0.6 | 0.308066 | 1.36079 | 2.31E-72 | lab | N |
| **MEAN CORP. HGB CONC.** | 0.59 | -0.05921 | 0.942505 | 0.067575 | lab | N |
| **HEMOGLOBIN** | 0.58 | -0.06242 | 0.939488 | 0.039214 | lab | Y |
| fibroids in pregnancy | 0.55 | 0.018031 | 1.018195 | 1.87E-09 | dx | N |
| Administrative/social admission | 0.55 | 0.01568 | 1.015803 | 3.51E-33 | dx | N |
| Hispanic | 0.54 | 0.401334 | 1.493817 | 2.38E-07 | demo | N |
| twin pregnancy | 0.52 | 0.015845 | 1.015971 | 2.59E-06 | dx | N |
| Benign neoplasm of uterus | 0.5 | 0.020208 | 1.020414 | 1.31E-10 | dx | N |
| **Other screening for suspected conditions (not mental disorders or infectious disease)** | 0.5 | 0.010497 | 1.010552 | 8.35E-15 | dx | N |
| **ALT(SGPT)** | 0.49 | 0.220039 | 1.246125 | 9.43E-14 | lab | N |
| large for GA | 0.48 | 0.008952 | 1.008992 | 0.000193 | dx | N |
| advanced maternal age | 0.48 | 0.005319 | 1.005333 | 0.00114 | dx | Y |

**Supplementary Table 15:**
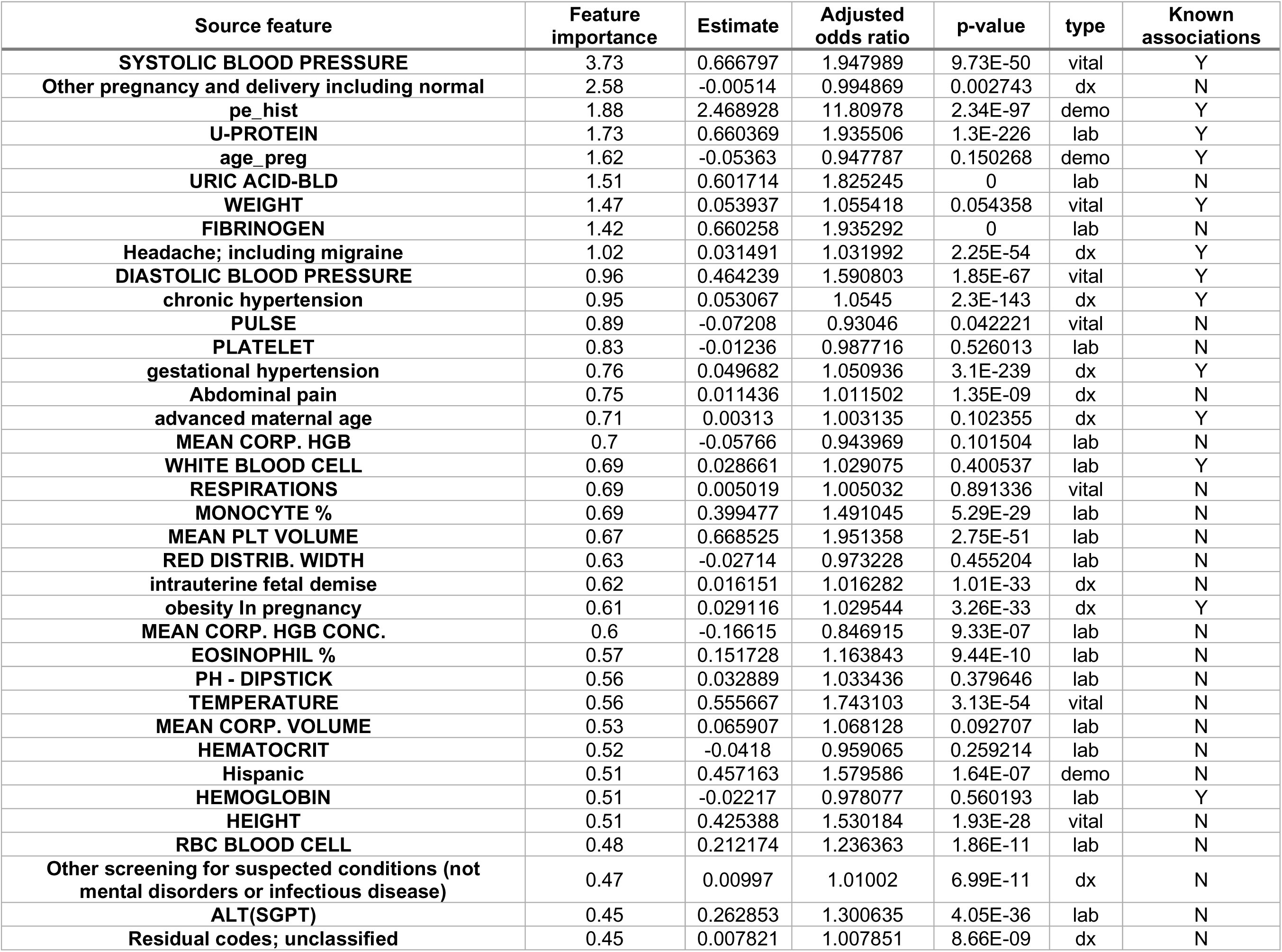
Selected unique features for model week 37.

**Supplementary Table 16:** Selected unique features for model week 38.

| Source feature | Feature importance | Estimate | Adjusted odds ratio | p-value | type | Known associations |
| --- | --- | --- | --- | --- | --- | --- |
| **SYSTOLIC BLOOD PRESSURE** | 1.77 | 0.708632 | 2.031211 | 5.54E-42 | vital | Y |
| **Other pregnancy and delivery including normal** | 1.46 | -0.00553 | 0.994489 | 0.006918 | dx | N |
| **WEIGHT** | 1.39 | 0.591225 | 1.806199 | 5.63E-42 | vital | Y |
| **pe_hist** | 1.13 | 2.397544 | 10.99614 | 1.54E-64 | demo | Y |
| **Headache; including migraine** | 1.07 | 0.033321 | 1.033883 | 5.78E-53 | dx | Y |
| **age_preg** | 1.06 | -0.08337 | 0.920008 | 0.052113 | demo | Y |
| **URIC ACID-BLD** | 1.02 | 0.59756 | 1.817678 | 4.4E-251 | lab | N |
| **FIBRINOGEN** | 0.88 | 0.656714 | 1.928445 | 2.6E-265 | lab | N |
| **PLATELET** | 0.78 | -0.01152 | 0.988546 | 0.898893 | lab | N |
| **TEMPERATURE** | 0.76 | 0.545392 | 1.725285 | 6.68E-40 | vital | N |
| **MEAN CORP. HGB CONC.** | 0.73 | -0.11111 | 0.894841 | 0.007002 | lab | N |
| **obesity In pregnancy** | 0.71 | 0.029139 | 1.029568 | 4.56E-26 | dx | Y |
| **DIASTOLIC BLOOD PRESSURE** | 0.69 | 0.440462 | 1.553425 | 2.73E-41 | vital | Y |
| **RESPIRATIONS** | 0.68 | 0.040801 | 1.041645 | 0.180907 | vital | N |
| **HEMATOCRIT** | 0.68 | -0.08239 | 0.920917 | 0.005788 | lab | N |
| **MEAN CORP. HGB** | 0.67 | -0.20279 | 0.81645 | 4.14E-09 | lab | N |
| **MEAN PLT VOLUME** | 0.64 | 0.684791 | 1.983358 | 1.77E-40 | lab | N |
| **U-PROTEIN** | 0.62 | 0.644881 | 1.90576 | 5.1E-164 | lab | Y |
| **Other circulatory disease** | 0.58 | 0.042533 | 1.043451 | 1.83E-71 | dx | N |
| **intrauterine fetal demise** | 0.57 | 0.016831 | 1.016973 | 1.62E-28 | dx | N |
| **Hispanic** | 0.53 | 0.417013 | 1.517423 | 4.31E-05 | demo | N |
| **nonreassuring fetal status** | 0.51 | 0.003795 | 1.003802 | 0.028266 | dx | N |
| **Benign neoplasm of uterus** | 0.51 | 0.021257 | 1.021484 | 1.45E-07 | dx | N |
| **Allergic reactions** | 0.47 | 0.017642 | 1.017799 | 1.61E-22 | dx | N |
| **MONOCYTE %** | 0.47 | 0.445079 | 1.560613 | 3.49E-31 | lab | N |
| **SPEC GRAVITY-DIPSTICK** | 0.46 | 0.008159 | 1.008193 | 0.779043 | lab | N |
| **RBC BLOOD CELL** | 0.46 | 0.058162 | 1.059887 | 0.176843 | lab | N |
| **Residual codes; unclassified** | 0.45 | 0.009113 | 1.009155 | 3.07E-09 | dx | N |
| **Abdominal pain** | 0.44 | 0.010506 | 1.010562 | 1.77E-06 | dx | N |
| **ALK PHOSPHATASE, BLD** | 0.44 | 0.331939 | 1.393667 | 9.61E-50 | lab | N |
| **PULSE** | 0.42 | -0.03886 | 0.961882 | 0.370665 | vital | N |
| **WHITE BLOOD CELL** | 0.42 | 0.101528 | 1.106861 | 0.007926 | lab | Y |
| **fibroids in pregnancy** | 0.42 | 0.018757 | 1.018934 | 1.57E-06 | dx | N |
| **GTT 3 HOUR** | 0.41 | 0.154483 | 1.167054 | 1.92E-06 | lab | N |
| **HEIGHT** | 0.41 | 0.424744 | 1.5292 | 1.02E-21 | vital | N |

**Supplementary Table 17:** Selected unique features for model week 39.

| Source feature | Feature importance | Estimate | Adjusted odds ratio | p-value | type | Known associations |
| --- | --- | --- | --- | --- | --- | --- |
| age_preg | 10.53 | -0.11124987 | 0.894715159 | 0.04448106 | demo | Y |
| Other pregnancy and delivery including normal | 9.55 | -0.004001481 | 0.996006514 | 0.153396547 | dx | N |
| intrauterine fetal demise | 3.17 | 0.015588482 | 1.015710617 | 1.14008E-15 | dx | N |
| Residual codes; unclassified | 3.16 | 0.008988653 | 1.009029173 | 5.38562E-06 | dx | N |
| Other screening for suspected conditions (not mental disorders or infectious disease) | 2.97 | 0.010421225 | 1.010475715 | 3.13878E-06 | dx | N |
| SYSTOLIC BLOOD PRESSURE | 2.59 | 0.761641504 | 2.141789091 | 2.61591E-29 | vital | Y |
| Headache; including migraine | 2.39 | 0.038011568 | 1.038743249 | 2.02811E-51 | dx | Y |
| DIASTOLIC BLOOD PRESSURE | 2.26 | -0.164154023 | 0.848611306 | 0.001691062 | vital | Y |
| FIBRINOGEN | 2.26 | 0.668933134 | 1.952153523 | 2.0781E-174 | lab | N |
| TEMPERATURE | 1.91 | 0.620172957 | 1.859249584 | 9.26509E-32 | vital | N |
| WEIGHT | 1.78 | 0.626795642 | 1.871603673 | 3.18649E-29 | vital | Y |
| pe_hist | 1.69 | 2.328142598 | 10.25886898 | 5.99289E-33 | demo | Y |
| Caucasian | 1.65 | -0.695580641 | 0.498784749 | 7.41951E-10 | demo | N |
| nonreassuring fetal status | 1.64 | 0.001503977 | 1.001505109 | 0.522886695 | dx | N |
| PULSE | 1.56 | -0.08296264 | 0.920385532 | 0.124683631 | vital | N |
| MEAN PLT VOLUME | 1.48 | 0.068216109 | 1.070596649 | 0.200320819 | lab | N |
| high risk pregnancy | 1.45 | 0.017521674 | 1.017676079 | 1.73223E-17 | dx | N |
| MEAN CORP. HGB | 1.34 | -0.050990629 | 0.950287575 | 0.330304422 | lab | N |
| MEAN CORP. VOLUME | 1.3 | -0.093044334 | 0.911153104 | 0.0532552 | lab | N |
| U-PROTEIN | 1.27 | 0.638368735 | 1.893389739 | 2.0134E-103 | lab | Y |
| Other circulatory disease | 1.26 | 0.042058453 | 1.042955441 | 1.07653E-43 | dx | N |
| RED DISTRIB. WIDTH | 1.25 | 0.069658883 | 1.072142393 | 0.099684345 | lab | N |
| WHITE BLOOD CELL | 1.23 | 0.190048181 | 1.209307861 | 3.14863E-05 | lab | Y |
| Allergic reactions | 1.22 | 0.018829603 | 1.019007998 | 4.41412E-18 | dx | N |
| AST (SGOT) | 1.19 | 0.297207029 | 1.346093951 | 2.06114E-32 | lab | N |
| URIC ACID-BLD | 1.15 | 0.595776927 | 1.814440085 | 1.1016E-159 | lab | N |
| MEAN CORP. HGB CONC. | 1.14 | -0.037068034 | 0.963610575 | 0.506923008 | lab | N |
| RESPIRATIONS | 1.1 | 0.068104436 | 1.070477099 | 0.038099008 | vital | N |
| advanced maternal age | 1.06 | 0.002144445 | 1.002146746 | 0.475452172 | dx | Y |
| Administrative/social admission | 1.04 | 0.016252014 | 1.016384796 | 8.13165E-14 | dx | N |
| HEIGHT | 1.02 | 0.436859851 | 1.547839134 | 8.91864E-15 | vital | N |
| fibroids in pregnancy | 1.01 | 0.018673628 | 1.01884907 | 0.000240715 | dx | N |
| PLATELET | 0.88 | -0.014015647 | 0.986082115 | 0.746480153 | lab | N |
| Asian | 0.85 | -0.707109076 | 0.49306756 | 0.016542086 | demo | N |
| ALT(SGPT) | 0.84 | 0.082703322 | 1.086219504 | 1.43744E-05 | lab | N |
| gestational hypertension | 0.82 | 0.043186052 | 1.04413214 | 3.37455E-74 | dx | Y |
| PH - DIPSTICK | 0.82 | 0.110267793 | 1.116577042 | 0.041730474 | lab | N |
| O2 SATURATION | 0.8 | -0.048512005 | 0.952645902 | 0.723358835 | vital | N |

**Supplementary Table 18:**
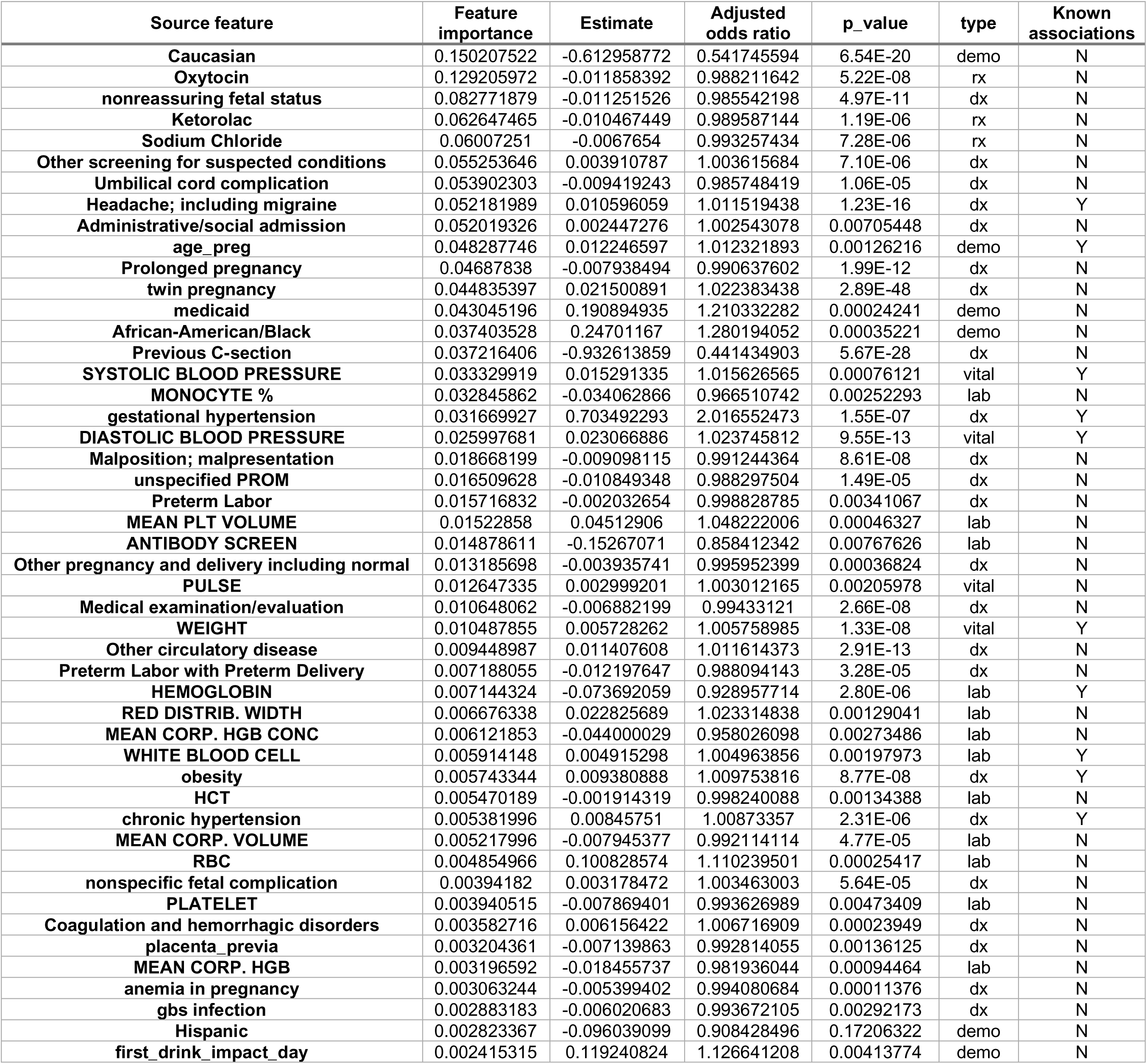
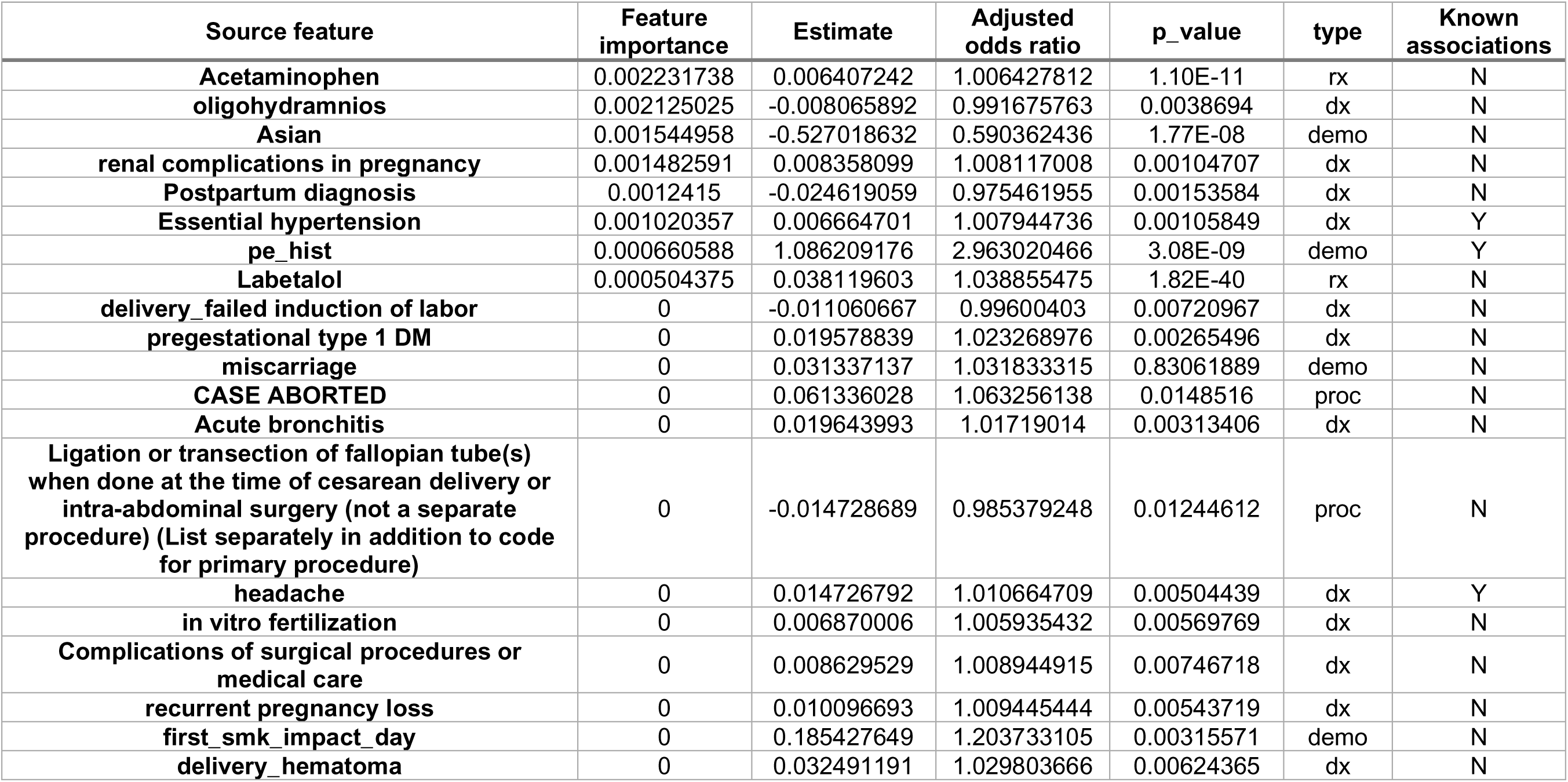
Selected unique features for model intrapartum.

| Source feature | Feature importance | Estimate | Adjusted odds ratio | p_value | type | Known associations |
| --- | --- | --- | --- | --- | --- | --- |
| Caucasian | 0.150207522 | -0.612958772 | 0.541745594 | 6.54E-20 | demo | N |
| Oxytocin | 0.129205972 | -0.011858392 | 0.988211642 | 5.22E-08 | rx | N |
| nonreassuring fetal status | 0.082771879 | -0.011251526 | 0.985542198 | 4.97E-11 | dx | N |
| Ketorolac | 0.062647465 | -0.010467449 | 0.989587144 | 1.19E-06 | rx | N |
| Sodium Chloride | 0.06007251 | -0.0067654 | 0.993257434 | 7.28E-06 | rx | N |
| Other screening for suspected conditions | 0.055253646 | 0.003910787 | 1.003615684 | 7.10E-06 | dx | N |
| Umbilical cord complication | 0.053902303 | -0.009419243 | 0.985748419 | 1.06E-05 | dx | N |
| Headache; including migraine | 0.052181989 | 0.010596059 | 1.011519438 | 1.23E-16 | dx | Y |
| Administrative/social admission | 0.052019326 | 0.002447276 | 1.002543078 | 0.00705448 | dx | N |
| age_preg | 0.048287746 | 0.012246597 | 1.012321893 | 0.00126216 | demo | Y |
| Prolonged pregnancy | 0.04687838 | -0.007938494 | 0.990637602 | 1.99E-12 | dx | N |
| twin pregnancy | 0.044835397 | 0.021500891 | 1.022383438 | 2.89E-48 | dx | N |
| medicaid | 0.043045196 | 0.190894935 | 1.210332282 | 0.00024241 | demo | N |
| African-American/Black | 0.037403528 | 0.24701167 | 1.280194052 | 0.00035221 | demo | N |
| Previous C-section | 0.037216406 | -0.932613859 | 0.441434903 | 5.67E-28 | dx | N |
| SYSTOLIC BLOOD PRESSURE | 0.033329919 | 0.015291335 | 1.015626565 | 0.00076121 | vital | Y |
| MONOCYTE % | 0.032845862 | -0.034062866 | 0.966510742 | 0.00252293 | lab | N |
| gestational hypertension | 0.031669927 | 0.703492293 | 2.016552473 | 1.55E-07 | dx | Y |
| DIASTOLIC BLOOD PRESSURE | 0.025997681 | 0.023066886 | 1.023745812 | 9.55E-13 | vital | Y |
| Malposition; malpresentation | 0.018668199 | -0.009098115 | 0.991244364 | 8.61E-08 | dx | N |
| unspecified PROM | 0.016509628 | -0.010849348 | 0.988297504 | 1.49E-05 | dx | N |
| Preterm Labor | 0.015716832 | -0.002032654 | 0.998828785 | 0.00341067 | dx | N |
| MEAN PLT VOLUME | 0.01522858 | 0.04512906 | 1.048222006 | 0.00046327 | lab | N |
| ANTIBODY SCREEN | 0.014878611 | -0.15267071 | 0.858412342 | 0.00767626 | lab | N |
| Other pregnancy and delivery including normal | 0.013185698 | -0.003935741 | 0.995952399 | 0.00036824 | dx | N |
| PULSE | 0.012647335 | 0.002999201 | 1.003012165 | 0.00205978 | vital | N |
| Medical examination/evaluation | 0.010648062 | -0.006882199 | 0.99433121 | 2.66E-08 | dx | N |
| WEIGHT | 0.010487855 | 0.005728262 | 1.005758985 | 1.33E-08 | vital | Y |
| Other circulatory disease | 0.009448987 | 0.011407608 | 1.011614373 | 2.91E-13 | dx | N |
| Preterm Labor with Preterm Delivery | 0.007188055 | -0.012197647 | 0.988094143 | 3.28E-05 | dx | N |
| HEMOGLOBIN | 0.007144324 | -0.073692059 | 0.928957714 | 2.80E-06 | lab | Y |
| RED DISTRIB. WIDTH | 0.006676338 | 0.022825689 | 1.023314838 | 0.00129041 | lab | N |
| MEAN CORP. HGB CONC | 0.006121853 | -0.044000029 | 0.958026098 | 0.00273486 | lab | N |
| WHITE BLOOD CELL | 0.005914148 | 0.004915298 | 1.004963856 | 0.00197973 | lab | Y |
| obesity | 0.005743344 | 0.009380888 | 1.009753816 | 8.77E-08 | dx | Y |
| HCT | 0.005470189 | -0.001914319 | 0.998240088 | 0.00134388 | lab | N |
| chronic hypertension | 0.005381996 | 0.00845751 | 1.00873357 | 2.31E-06 | dx | Y |
| MEAN CORP. VOLUME | 0.005217996 | -0.007945377 | 0.992114114 | 4.77E-05 | lab | N |
| RBC | 0.004854966 | 0.100828574 | 1.110239501 | 0.00025417 | lab | N |
| nonspecific fetal complication | 0.00394182 | 0.003178472 | 1.003463003 | 5.64E-05 | dx | N |
| PLATELET | 0.003940515 | -0.007869401 | 0.993626989 | 0.00473409 | lab | N |
| Coagulation and hemorrhagic disorders | 0.003582716 | 0.006156422 | 1.006716909 | 0.00023949 | dx | N |
| placenta_previa | 0.003204361 | -0.007139863 | 0.992814055 | 0.00136125 | dx | N |
| MEAN CORP. HGB | 0.003196592 | -0.018455737 | 0.981936044 | 0.00094464 | lab | N |
| anemia in pregnancy | 0.003063244 | -0.005399402 | 0.994080684 | 0.00011376 | dx | N |
| gbs infection | 0.002883183 | -0.006020683 | 0.993672105 | 0.00292173 | dx | N |
| Hispanic | 0.002823367 | -0.096039099 | 0.908428496 | 0.17206322 | demo | N |
| first_drink_impact_day | 0.002415315 | 0.119240824 | 1.126641208 | 0.00413774 | demo | N |

**Supplementary Table 18: Selected unique features for model intrapartum (continued)**
| Source feature | Feature importance | Estimate | Adjusted odds ratio | p_value | type | Known associations |
| --- | --- | --- | --- | --- | --- | --- |
| Acetaminophen | 0.002231738 | 0.006407242 | 1.006427812 | 1.10E-11 | rx | N |
| oligohydramnios | 0.002125025 | -0.008065892 | 0.991675763 | 0.0038694 | dx | N |
| Asian | 0.001544958 | -0.527018632 | 0.590362436 | 1.77E-08 | demo | N |
| renal complications in pregnancy | 0.001482591 | 0.008358099 | 1.008117008 | 0.00104707 | dx | N |
| Postpartum diagnosis | 0.0012415 | -0.024619059 | 0.975461955 | 0.00153584 | dx | N |
| Essential hypertension | 0.001020357 | 0.006664701 | 1.007944736 | 0.00105849 | dx | Y |
| pe_hist | 0.000660588 | 1.086209176 | 2.963020466 | 3.08E-09 | demo | Y |
| Labetalol | 0.000504375 | 0.038119603 | 1.038855475 | 1.82E-40 | rx | N |
| delivery_failed induction of labor | 0 | -0.011060667 | 0.99600403 | 0.00720967 | dx | N |
| pregestational type 1 DM | 0 | 0.019578839 | 1.023268976 | 0.00265496 | dx | N |
| miscarriage | 0 | 0.031337137 | 1.031833315 | 0.83061889 | demo | N |
| CASE ABORTED | 0 | 0.061336028 | 1.063256138 | 0.0148516 | proc | N |
| Acute bronchitis | 0 | 0.019643993 | 1.01719014 | 0.00313406 | dx | N |
| Ligation or transection of fallopian tube(s) when done at the time of cesarean delivery or intra-abdominal surgery (not a separate procedure) (List separately in addition to code for primary procedure) | 0 | -0.014728689 | 0.985379248 | 0.01244612 | proc | N |
| headache | 0 | 0.014726792 | 1.010664709 | 0.00504439 | dx | Y |
| in vitro fertilization | 0 | 0.006870006 | 1.005935432 | 0.00569769 | dx | N |
| Complications of surgical procedures or medical care | 0 | 0.008629529 | 1.008944915 | 0.00746718 | dx | N |
| recurrent pregnancy loss | 0 | 0.010096693 | 1.009445444 | 0.00543719 | dx | N |
| first_smk_impact_day | 0 | 0.185427649 | 1.203733105 | 0.00315571 | demo | N |
| delivery_hematoma | 0 | 0.032491191 | 1.029803666 | 0.00624365 | dx | N |

**Supplementary Table 19:** Selected unique features for model postpartum.

| Source feature | Feature importance | Estimate | Adjusted odds ratio | p_value | type | Known associations |
| --- | --- | --- | --- | --- | --- | --- |
| Ibuprofen | 0.364716606 | -0.01296254 | 0.987121112 | 1.04E-30 | rx | N |
| Caucasian | 0.240070946 | -0.397921519 | 0.671714743 | 0.022826759 | demo | N |
| OB-related trauma to perineum and vulva | 0.213517372 | -0.013542605 | 0.985265174 | 3.97E-21 | dx | N |
| age_preg | 0.17398007 | 0.038154054 | 1.038891266 | 4.30E-05 | demo | Y |
| Other aftercare | 0.154464294 | -0.01818284 | 0.981693531 | 2.45E-15 | dx | N |
| Administrative/social admission | 0.137355629 | 0.005672244 | 1.007675073 | 0.001819962 | dx | N |
| Contraceptive and procreative management | 0.128979895 | -0.009443669 | 0.990932444 | 8.88E-07 | dx | N |
| Umbilical cord complication | 0.104475754 | -0.005182352 | 0.994909965 | 0.0002134 | dx | N |
| gestational hypertension | 0.101781649 | 0.018124124 | 1.018196444 | 1.90E-28 | dx | Y |
| African-American/Black | 0.099505476 | 0.850423852 | 2.340638725 | 8.31E-07 | demo | N |
| Immunizations and screening for infectious diseases | 0.087812433 | -0.006912188 | 0.993499583 | 3.02E-06 | dx | N |
| SYSTOLIC BLOOD PRESSURE | 0.06972831 | 0.017756126 | 1.01830803 | 0.001350675 | vital | Y |
| PULSE | 0.059997127 | 0.010106951 | 1.010203362 | 0.001135782 | vital | N |
| Medical examination/evaluation | 0.059511343 | -0.00557685 | 0.99311459 | 0.006598619 | dx | N |
| Headache; including migraine | 0.0583503 | 0.006450628 | 1.008291684 | 0.00545399 | dx | Y |
| DIASTOLIC BLOOD PRESSURE | 0.054403084 | 0.01945539 | 1.020098368 | 0.000130308 | vital | Y |
| MEAN CORP. VOLUME | 0.053784586 | -0.008597857 | 0.99146911 | 0.007918128 | lab | N |
| WHITE BLOOD CELL | 0.046290387 | -0.000641479 | 0.999447766 | 0.003521754 | lab | Y |
| TEMPERATURE | 0.045716487 | 0.219254459 | 1.29755792 | 0.001654393 | vital | N |
| unspecified fluid disorder | 0.042850071 | -0.010161876 | 0.990712458 | 0.000346916 | dx | N |
| Allergic reactions | 0.040199987 | -0.005135422 | 0.994413624 | 0.000179517 | dx | N |
| HCT | 0.039177449 | -0.026837951 | 0.973623319 | 0.003051283 | lab | N |
| HEMOGLOBIN | 0.033595806 | -0.088121674 | 0.917055952 | 0.002901927 | lab | Y |
| MEAN CORP. HGB | 0.032618618 | -0.0396916 | 0.961085793 | 0.008396058 | lab | N |
| medicaid | 0.031659207 | 0.159002897 | 1.172341343 | 0.187813075 | demo | N |
| RESPIRATIONS | 0.028930863 | 0.044547228 | 1.046981541 | 0.001144948 | vital | N |
| chronic hypertension | 0.027141798 | 0.008299004 | 1.006927645 | 0.001890334 | dx | Y |
| WEIGHT | 0.025493696 | 0.006233304 | 1.006271732 | 0.001307854 | vital | Y |
| Residual codes; unclassified | 0.025284996 | -0.004883181 | 0.996869887 | 0.000813826 | dx | N |
| decreased fetal movement | 0.024260535 | 0.006941224 | 1.007660168 | 0.000256835 | dx | N |
| Hispanic | 0.015932268 | 0.356425473 | 1.428215086 | 0.047964851 | demo | N |
| Labetalol | 0.010603545 | 0.029787345 | 1.030235426 | 2.88E-33 | rx | N |
| mental health | 0.010160836 | 0.01181943 | 1.012007109 | 0.001298682 | dx | N |
| pe_hist | 0.008054557 | 1.898771907 | 6.677688583 | 2.02E-11 | demo | Y |
| twin pregnancy | 0.006906508 | 0.011611433 | 1.012797965 | 1.12E-05 | dx | N |
| Other circulatory disease | 0.006237221 | 0.006950202 | 1.007174409 | 0.00536639 | dx | N |
| first_drink_impact_day | 0.003864094 | 0.272437872 | 1.313161872 | 0.003097715 | demo | N |
| Other endocrine disorders | 0.002160961 | 0.01208437 | 1.010729811 | 0.000606789 | dx | N |
| Hypertension with complications and secondary hypertension | 0.00192391 | 0.021280143 | 1.019747704 | 0.000317305 | dx | Y |
| first_smk_impact_day | 0.001369617 | 0.049109645 | 1.050335508 | 0.72639165 | demo | N |
| Laparoscopy, surgical; cholecystectomy | 0.000887704 | 0.037601009 | 1.038316871 | 0.028805294 | proc | N |
| Endocervical curettage (not done as part of a dilation and curettage) | 0.000733898 | 0.03674391 | 1.037427312 | 0.021154699 | proc | N |
| Laparoscopy, surgical, repair, ventral, umbilical, spigelian or epigastric hernia (includes mesh insertion, when performed) | 0 | 0.04337429 | 1.044328703 | 0.015733315 | proc | N |

Supplementary Table 19: Selected unique features for model postpartum (continued)
| Source feature | Feature importance | Estimate | Adjusted odds ratio | p_value | type | Known associations |
| --- | --- | --- | --- | --- | --- | --- |
| Total abdominal hysterectomy (corpus and cervix), with or without removal of tube(s), with or without removal of ovary(s) | 0 | 0.020873501 | 1.021092877 | 0.023667936 | proc | N |
| Laparoscopic treatment of ectopic pregnancy; without salpingectomy and/or oophorectomy | 0 | 0.134467869 | 1.143927902 | 0.030601679 | proc | N |
| Asian | 0 | -0.062982727 | 0.938959692 | 0.782407711 | demo | N |
| hellp syndrome (severe preeclampsia that affects liver, platelets, hemolysis) | 0 | 0.060868247 | 1.054113349 | 8.58E-10 | dx | N |
| miscarriage | 0 | -0.055010964 | 0.94647477 | 0.848525972 | demo | N |

**Supplementary Table 20:** Metrics for MSH training dataset.

| week | ACOG (high risk factors) AUC | ACOG (all risk factors) AUC | SEN | SPE | F1 score | ACC | PPV | NPV | AUC | AP |
| --- | --- | --- | --- | --- | --- | --- | --- | --- | --- | --- |
| 4 | 0.621 [0.617, 0.626] | 0.671 [0.666, 0.677] | 0.538 [0.523, 0.562] | 0.752 [0.743, 0.762] | 0.066 [0.065, 0.068] | 0.749 [0.739, 0.758] | 0.035 [0.034, 0.036] | 0.990 [0.989, 0.990] | 0.688 [0.680, 0.696] | 0.081 [0.064, 0.096] |
| 8 |  |  | 0.534 [0.533, 0.575] | 0.790 [0.788, 0.794] | 0.079 [0.077, 0.081] | 0.787 [0.784, 0.789] | 0.042 [0.042, 0.043] | 0.990 [0.990, 0.991] | 0.707 [0.694, 0.732] | 0.099 [0.060, 0.117] |
| 12 |  |  | 0.528 [0.493, 0.534] | 0.845 [0.828, 0.855] | 0.094 [0.090, 0.098] | 0.839 [0.824, 0.848] | 0.052 [0.049, 0.054] | 0.991 [0.990, 0.991] | 0.739 [0.730, 0.750] | 0.091 [0.079, 0.113] |
| 16 |  |  | 0.533 [0.503, 0.569] | 0.844 [0.837, 0.855] | 0.100 [0.099, 0.104] | 0.839 [0.833, 0.849] | 0.055 [0.054, 0.058] | 0.991 [0.990, 0.992] | 0.747 [0.742, 0.763] | 0.096 [0.096, 0.106] |
| 20 |  |  | 0.551 [0.496, 0.562] | 0.856 [0.843, 0.864] | 0.103 [0.098, 0.109] | 0.850 [0.839, 0.858] | 0.057 [0.054, 0.062] | 0.991 [0.990, 0.992] | 0.764 [0.758, 0.777] | 0.103 [0.090, 0.114] |
| 22 |  |  | 0.534 [0.511, 0.597] | 0.857 [0.840, 0.867] | 0.110 [0.098, 0.117] | 0.852 [0.836, 0.862] | 0.061 [0.054, 0.065] | 0.991 [0.990, 0.992] | 0.765 [0.762, 0.777] | 0.105 [0.081, 0.142] |
| 24 |  |  | 0.547 [0.518, 0.575] | 0.863 [0.855, 0.875] | 0.114 [0.103, 0.121] | 0.859 [0.850, 0.869] | 0.063 [0.057, 0.068] | 0.991 [0.991, 0.992] | 0.775 [0.770, 0.783] | 0.114 [0.109, 0.145] |
| 26 |  |  | 0.568 [0.529, 0.584] | 0.863 [0.854, 0.873] | 0.118 [0.115, 0.126] | 0.858 [0.850, 0.868] | 0.065 [0.064, 0.071] | 0.992 [0.991, 0.992] | 0.786 [0.781, 0.800] | 0.141 [0.118, 0.149] |
| 28 |  |  | 0.583 [0.514, 0.626] | 0.888 [0.879, 0.898] | 0.137 [0.130, 0.151] | 0.883 [0.873, 0.894] | 0.079 [0.073, 0.085] | 0.992 [0.991, 0.993] | 0.814 [0.786, 0.838] | 0.148 [0.128, 0.189] |
| 30 |  |  | 0.577 [0.572, 0.610] | 0.887 [0.882, 0.902] | 0.149 [0.127, 0.158] | 0.883 [0.878, 0.896] | 0.084 [0.071, 0.091] | 0.992 [0.992, 0.993] | 0.820 [0.805, 0.831] | 0.186 [0.150, 0.200] |
| 32 |  |  | 0.594 [0.561, 0.623] | 0.911 [0.908, 0.920] | 0.165 [0.160, 0.183] | 0.907 [0.904, 0.914] | 0.096 [0.092, 0.106] | 0.993 [0.992, 0.994] | 0.835 [0.824, 0.851] | 0.214 [0.199, 0.243] |
| 34 |  |  | 0.666 [0.586, 0.728] | 0.925 [0.919, 0.936] | 0.197 [0.174, 0.218] | 0.922 [0.914, 0.931] | 0.116 [0.100, 0.131] | 0.995 [0.993, 0.996] | 0.852 [0.848, 0.868] | 0.234 [0.185, 0.316] |
| 35 |  |  | 0.701 [0.666, 0.727] | 0.933 [0.929, 0.943] | 0.219 [0.186, 0.232] | 0.929 [0.926, 0.941] | 0.130 [0.110, 0.139] | 0.996 [0.995, 0.996] | 0.869 [0.864, 0.907] | 0.242 [0.197, 0.284] |
| 36 |  |  | 0.768 [0.714, 0.800] | 0.934 [0.929, 0.947] | 0.215 [0.195, 0.247] | 0.932 [0.927, 0.944] | 0.127 [0.112, 0.146] | 0.997 [0.996, 0.998] | 0.895 [0.884, 0.920] | 0.259 [0.232, 0.298] |
| 37 |  |  | 0.776 [0.762, 0.809] | 0.929 [0.927, 0.943] | 0.168 [0.161, 0.183] | 0.928 [0.926, 0.941] | 0.094 [0.089, 0.104] | 0.998 [0.998, 0.998] | 0.915 [0.885, 0.924] | 0.180 [0.170, 0.197] |
| 38 |  |  | 0.775 [0.697, 0.811] | 0.936 [0.913, 0.950] | 0.141 [0.110, 0.166] | 0.934 [0.912, 0.949] | 0.077 [0.059, 0.093] | 0.998 [0.998, 0.999] | 0.906 [0.889, 0.922] | 0.113 [0.078, 0.140] |
| 39 |  |  | 0.697 [0.617, 0.816] | 0.951 [0.942, 0.957] | 0.100 [0.088, 0.138] | 0.948 [0.941, 0.956] | 0.053 [0.047, 0.077] | 0.998 [0.998, 0.999] | 0.898 [0.885, 0.929] | 0.076 [0.058, 0.085] |
| Intrapartum | N/A | N/A | 0.646 [0.620, 0.650] | 0.841 [0.839, 0.843] | 0.322 [0.315, 0.328] | 0.828 [0.825, 0.830] | 0.215 [0.210, 0.220] | 0.972 [0.970, 0.972] | 0.820 [0.815, 0.825] | 0.352 [0.338, 0.361] |
| Postpartum | N/A | N/A | 0.530 [0.479, 0.572] | 0.951 [0.948, 0.963] | 0.195 [0.176, 0.224] | 0.946 [0.943, 0.958] | 0.118 [0.105, 0.147] | 0.994 [0.994, 0.995] | 0.893 [0.888, 0.899] | 0.194 [0.182, 0.244] |

**Supplementary Table 21:**
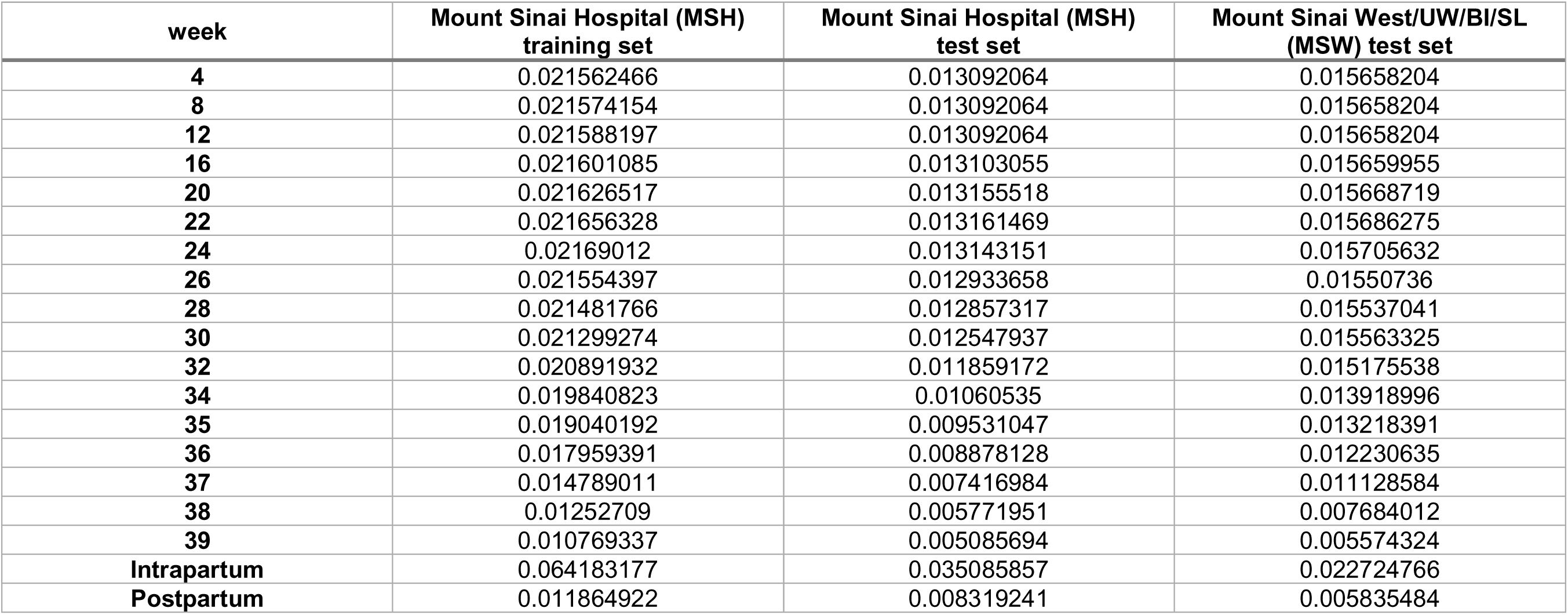
Preeclampsia prevalence at different datasets.

| **week** | **Mount Sinai Hospital (MSH)**<br>**training set** | **Mount Sinai Hospital (MSH)**<br>**test set** | **Mount Sinai West/UW/BI/SL**<br>**(MSW) test set** |
| --- | --- | --- | --- |
| **4** | 0.021562466 | 0.013092064 | 0.015658204 |
| **8** | 0.021574154 | 0.013092064 | 0.015658204 |
| **12** | 0.021588197 | 0.013092064 | 0.015658204 |
| **16** | 0.021601085 | 0.013103055 | 0.015659955 |
| **20** | 0.021626517 | 0.013155518 | 0.015668719 |
| **22** | 0.021656328 | 0.013161469 | 0.015686275 |
| **24** | 0.02169012 | 0.013143151 | 0.015705632 |
| **26** | 0.021554397 | 0.012933658 | 0.01550736 |
| **28** | 0.021481766 | 0.012857317 | 0.015537041 |
| **30** | 0.021299274 | 0.012547937 | 0.015563325 |
| **32** | 0.020891932 | 0.011859172 | 0.015175538 |
| **34** | 0.019840823 | 0.01060535 | 0.013918996 |
| **35** | 0.019040192 | 0.009531047 | 0.013218391 |
| **36** | 0.017959391 | 0.008878128 | 0.012230635 |
| **37** | 0.014789011 | 0.007416984 | 0.011128584 |
| **38** | 0.01252709 | 0.005771951 | 0.007684012 |
| **39** | 0.010769337 | 0.005085694 | 0.005574324 |
| **Intrapartum** | 0.064183177 | 0.035085857 | 0.022724766 |
| **Postpartum** | 0.011864922 | 0.008319241 | 0.005835484 |

**Supplementary Table 22:** Validation performance at MSH test dataset.

| week | ACOG (high risk factors) AUC | ACOG (all risk factors) AUC | SEN | SPE | F1 score | ACC | PPV | NPV | AUC | AP |
| --- | --- | --- | --- | --- | --- | --- | --- | --- | --- | --- |
| 4 | 0.583 [0.577, 0.589] | 0.662 [0.653, 0.672] | 0.440 [0.427, 0.452] | 0.821 [0.806, 0.832] | 0.062 [0.059, 0.063] | 0.816 [0.801, 0.826] | 0.033 [0.032, 0.034] | 0.990 [0.990, 0.990] | 0.658 [0.655, 0.668] | 0.048 [0.046, 0.049] |
| 8 |  |  | 0.578 [0.565, 0.596] | 0.795 [0.769, 0.809] | 0.112 [0.106, 0.115] | 0.790 [0.766, 0.803] | 0.062 [0.058, 0.064] | 0.988 [0.987, 0.988] | 0.755 [0.749, 0.764] | 0.085 [0.083, 0.093] |
| 12 |  |  | 0.552 [0.527, 0.562] | 0.814 [0.798, 0.829] | 0.116 [0.111, 0.122] | 0.808 [0.793, 0.823] | 0.065 [0.061, 0.069] | 0.987 [0.987, 0.988] | 0.744 [0.738, 0.746] | 0.091 [0.085, 0.095] |
| 16 |  |  | 0.580 [0.552, 0.596] | 0.797 [0.770, 0.812] | 0.113 [0.104, 0.123] | 0.791 [0.765, 0.806] | 0.063 [0.057, 0.069] | 0.987 [0.987, 0.988] | 0.741 [0.732, 0.745] | 0.106 [0.099, 0.111] |
| 20 |  |  | 0.546 [0.517, 0.578] | 0.831 [0.792, 0.846] | 0.126 [0.114, 0.131] | 0.824 [0.787, 0.838] | 0.071 [0.063, 0.074] | 0.987 [0.987, 0.987] | 0.748 [0.742, 0.751] | 0.121 [0.108, 0.127] |
| 22 |  |  | 0.554 [0.516, 0.596] | 0.825 [0.786, 0.854] | 0.125 [0.115, 0.133] | 0.818 [0.781, 0.846] | 0.071 [0.064, 0.077] | 0.987 [0.986, 0.988] | 0.758 [0.751, 0.759] | 0.104 [0.093, 0.116] |
| 24 |  |  | 0.565 [0.554, 0.580] | 0.828 [0.809, 0.847] | 0.129 [0.122, 0.145] | 0.821 [0.803, 0.841] | 0.073 [0.068, 0.083] | 0.987 [0.987, 0.988] | 0.761 [0.758, 0.772] | 0.120 [0.115, 0.141] |
| 26 |  |  | 0.584 [0.564, 0.599] | 0.823 [0.811, 0.856] | 0.135 [0.129, 0.157] | 0.817 [0.806, 0.849] | 0.076 [0.073, 0.091] | 0.987 [0.987, 0.988] | 0.786 [0.776, 0.790] | 0.141 [0.134, 0.179] |
| 28 |  |  | 0.593 [0.585, 0.612] | 0.858 [0.848, 0.877] | 0.166 [0.160, 0.179] | 0.852 [0.842, 0.870] | 0.096 [0.092, 0.106] | 0.989 [0.988, 0.989] | 0.801 [0.798, 0.805] | 0.200 [0.169, 0.208] |
| 30 |  |  | 0.619 [0.606, 0.631] | 0.867 [0.858, 0.884] | 0.176 [0.171, 0.194] | 0.861 [0.852, 0.877] | 0.103 [0.099, 0.115] | 0.989 [0.989, 0.989] | 0.815 [0.812, 0.820] | 0.251 [0.245, 0.257] |
| 32 |  |  | 0.652 [0.634, 0.662] | 0.873 [0.852, 0.885] | 0.193 [0.167, 0.199] | 0.869 [0.847, 0.879] | 0.112 [0.096, 0.118] | 0.990 [0.990, 0.990] | 0.829 [0.825, 0.832] | 0.257 [0.236, 0.271] |
| 34 |  |  | 0.613 [0.587, 0.630] | 0.903 [0.885, 0.917] | 0.213 [0.197, 0.239] | 0.896 [0.879, 0.909] | 0.129 [0.117, 0.148] | 0.990 [0.989, 0.990] | 0.834 [0.830, 0.837] | 0.291 [0.274, 0.320] |
| 35 |  |  | 0.620 [0.615, 0.641] | 0.912 [0.908, 0.931] | 0.223 [0.216, 0.260] | 0.905 [0.902, 0.924] | 0.136 [0.130, 0.164] | 0.991 [0.991, 0.991] | 0.852 [0.849, 0.854] | 0.277 [0.254, 0.319] |
| 36 |  |  | 0.645 [0.633, 0.673] | 0.920 [0.913, 0.934] | 0.233 [0.224, 0.261] | 0.914 [0.908, 0.928] | 0.142 [0.135, 0.164] | 0.992 [0.992, 0.993] | 0.868 [0.862, 0.871] | 0.277 [0.232, 0.336] |
| 37 |  |  | 0.668 [0.659, 0.682] | 0.918 [0.911, 0.929] | 0.225 [0.205, 0.239] | 0.914 [0.907, 0.924] | 0.135 [0.122, 0.146] | 0.993 [0.993, 0.994] | 0.865 [0.859, 0.870] | 0.183 [0.178, 0.215] |
| 38 |  |  | 0.615 [0.584, 0.662] | 0.923 [0.887, 0.941] | 0.162 [0.129, 0.192] | 0.919 [0.884, 0.936] | 0.094 [0.072, 0.115] | 0.995 [0.994, 0.995] | 0.844 [0.834, 0.851] | 0.126 [0.104, 0.172] |
| 39 |  |  | 0.548 [0.540, 0.602] | 0.943 [0.920, 0.945] | 0.171 [0.150, 0.182] | 0.938 [0.916, 0.940] | 0.103 [0.086, 0.109] | 0.994 [0.994, 0.995] | 0.834 [0.824, 0.839] | 0.108 [0.092, 0.131] |
| Intrapartum | N/A | N/A | 0.444 [0.437, 0.453] | 0.925 [0.919, 0.929] | 0.268 [0.263, 0.272] | 0.907 [0.901, 0.910] | 0.194 [0.185, 0.196] | 0.976 [0.976, 0.977] | 0.833 [0.831, 0.834] | 0.208 [0.205, 0.211] |
| Postpartum | N/A | N/A | 0.516 [0.491, 0.543] | 0.950 [0.941, 0.953] | 0.138 [0.126, 0.139] | 0.946 [0.937, 0.949] | 0.080 [0.071, 0.081] | 0.996 [0.996, 0.996] | 0.842 [0.841, 0.846] | 0.108 [0.105, 0.111] |

**Supplementary Table 23:**
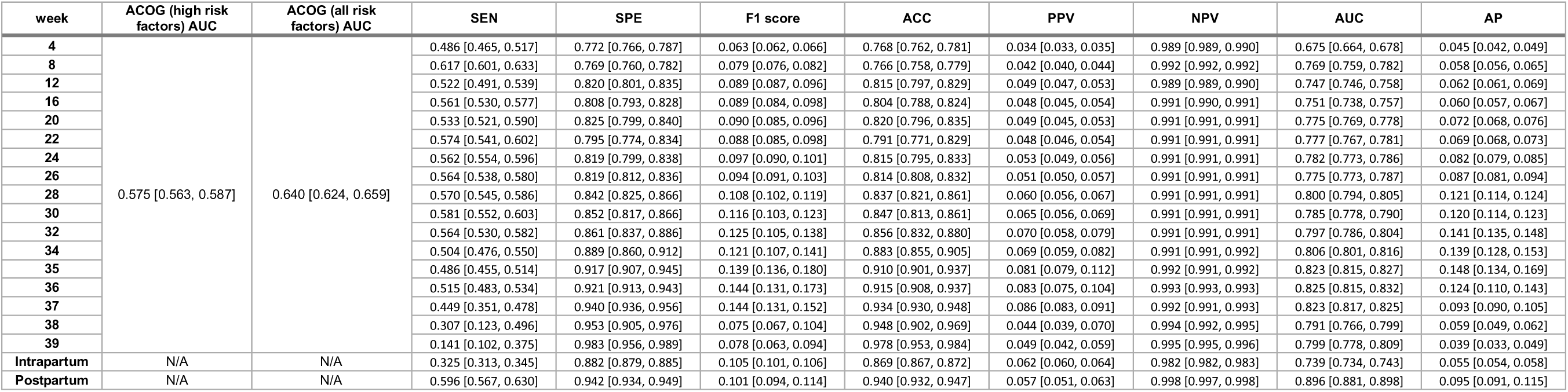
Validation performance at MSW test dataset.

**Supplementary Table 24:** Clinical feature characteristics for each patient across pregnancy.

| Feature type | Overall (std) | prior pregnancy (std) | pregnancy to delivery (std) | Postpartum (std) |
| --- | --- | --- | --- | --- |
| Procedures by CPT4 | 1.12 (0.43) | 1.09 (0.34) | 1.05 (0.24) | 1.23 (0.49) |
| Diagnosis by ICD9/10 | 21.75 (17.16) | 10.17 (6.67) | 18.62 (14.51) | 7.8 (8.48) |
| labs by LOINC | 35.24 (19.03) | 19.99 (12.33) | 32.74 (16.97) | 23.24 (17.89) |
| Drugs by Name | 10.63 (8.35) | 5.63 (4.95) | 7.27 (5.58) | 4.89 (5.71) |
| # of vital measurements | 246.9 (273.76) | 97.57 (140.34) | 160.13 (192.95) | 49.63 (130.3) |

## References

1. Shih, T. et al. The Rising Burden of Preeclampsia in the United States Impacts Both Maternal and Child Health. American Journal of Perinatology (2016) doi:10.1055/s-0035-1564881.

2. Huppertz, B. Biology of preeclampsia: Combined actions of angiogenic factors, their receptors and placental proteins. Biochim. Biophys. Acta - Mol. Basis Dis. (2020) doi:10.1016/j.bbadis.2018.11.024.

3. Seidler, A. L., Askie, L. & Ray, J. G. Optimal aspirin dosing for preeclampsia prevention. American Journal of Obstetrics and Gynecology (2018) doi:10.1016/j.ajog.2018.03.018.

4. Skalis, G. et al. MicroRNAs in Preeclampsia. MicroRNA (2018) doi:10.2174/2211536607666180813123303.

5. Nobakht M. Gh, B. F. Application of metabolomics to preeclampsia diagnosis. Systems Biology in Reproductive Medicine (2018) doi:10.1080/19396368.2018.1482968.

6. Tarca, A. L. et al. The prediction of early preeclampsia: Results from a longitudinal proteomics study. PLoS One (2019) doi:10.1371/journal.pone.0217273.

7. Gray, K. J., Saxena, R. & Karumanchi, S. A. Genetic predisposition to preeclampsia is conferred by fetal DNA variants near FLT1, a gene involved in the regulation of angiogenesis. Am. J. Obstet. Gynecol. (2018) doi:10.1016/j.ajog.2017.11.562.

8. Brodowski, L. et al. Preeclampsia-associated alteration of DNA methylation in fetal endothelial progenitor cells. Front. Cell Dev. Biol. (2019) doi:10.3389/fcell.2019.00032.

9. Liu, L. Y. et al. Integrating multiple ‘omics’ analyses identifies serological protein biomarkers for preeclampsia. BMC Med. (2013) doi:10.1186/1741-7015-11-236.

10. Serra, B. et al. A new model for screening for early-onset preeclampsia. Am. J. Obstet. Gynecol. (2020) doi:10.1016/j.ajog.2020.01.020.

11. Copel, J. A. et al. Gottesfeld-Hohler Memorial Foundation Risk Assessment for Early-Onset Preeclampsia in the United States: Think Tank Summary. Obstet. Gynecol. (2020) doi:10.1097/AOG.0000000000003582.

12. ACOG Practice Bulletin No. 202: Gestational Hypertension and Preeclampsia. Obstet. Gynecol. (2019) doi:10.1097/AOG.0000000000003018.

13. Jeyabalan, A. Epidemiology of preeclampsia: Impact of obesity. Nutr. Rev. (2013) doi:10.1111/nure.12055.

14. Green, L. J. et al. Gestation-Specific Vital Sign Reference Ranges in Pregnancy. Obstet. Gynecol. (2020) doi:10.1097/AOG.0000000000003721.

15. Practice, A. A. P. C. on F. and N. and A. C. on O. Guidelines for Perinatal Care, 8th Edition. (American Academy of Pediatrics, 2017).

16. Sibai, B. M. et al. Risk factors for preeclampsia in healthy nulliparous women: a prospective multicenter study. The National Institute of Child Health and Human Development Network of Maternal-Fetal Medicine Units. Am. J. Obstet. Gynecol. (1995).

17. Nørgaard, S. K. et al. Diastolic blood pressure is a potentially modifiable risk factor for preeclampsia in women with pre-existing diabetes. Diabetes Res. Clin. Pract. (2018) doi:10.1016/j.diabres.2018.02.014.

18. Conde-Agudelo, A. & Belizán, J. M. Risk factors for pre-eclampsia in a large cohort of Latin American and Caribbean women. *BJOG An Int*. J. Obstet. Gynaecol. (2000) doi:10.1111/j.1471-0528.2000.tb11582.x.

19. Anderson, U. D., Jälmby, M., Faas, M. M. & Hansson, S. R. The hemoglobin degradation pathway in patients with preeclampsia – Fetal hemoglobin, heme, heme oxygenase-1 and hemopexin – Potential diagnostic biomarkers? Pregnancy Hypertens. (2018) doi:10.1016/j.preghy.2018.02.005.

20. Sitotaw, C., Asrie, F. & Melku, M. Evaluation of platelet and white cell parameters among pregnant women with Preeclampsia in Gondar, Northwest Ethiopia: A comparative cross-sectional study. Pregnancy Hypertens. (2018) doi:10.1016/j.preghy.2018.06.006.

21. Bartsch, E. et al. Clinical risk factors for pre-eclampsia determined in early pregnancy: Systematic review and meta-analysis of large cohort studies. BMJ (2016) doi:10.1136/bmj.i1753.

22. Sperling, J. D., Dahlke, J. D., Huber, W. J. & Sibai, B. M. The Role of Headache in the Classification and Management of Hypertensive Disorders in Pregnancy. Obstetrics and Gynecology (2015) doi:10.1097/AOG.0000000000000966.

23. Tolcher, M. C. et al. Impact of USPSTF recommendations for aspirin for prevention of recurrent preeclampsia. Am. J. Obstet. Gynecol. (2017) doi:10.1016/j.ajog.2017.04.035.

24. Wagner, J. L., White, R. S., Tangel, V., Gupta, S. & Pick, J. S. Socioeconomic, Racial, and Ethnic Disparities in Postpartum Readmissions in Patients with Preeclampsia: a Multi-state Analysis, 2007–2014. J. Racial Ethn. Heal. Disparities (2019) doi:10.1007/s40615-019-00580-1.

25. Manten, G. T. R. et al. Increased high molecular weight fibrinogen in pre-eclampsia. Thromb. Res. (2003) doi:10.1016/j.thromres.2003.08.025.

26. Vilchez, G., Lagos, M., Kumar, K. & Argoti, P. Is mean platelet volume a better biomarker in pre-eclampsia? J. Obstet. Gynaecol. Res. (2017) doi:10.1111/jog.13312.

27. Yücel, B. & Ustun, B. Neutrophil to lymphocyte ratio, platelet to lymphocyte ratio, mean platelet volume, red cell distribution width and plateletcrit in preeclampsia. Pregnancy Hypertens. (2017) doi:10.1016/j.preghy.2016.12.002.

28. Kupfermine, M. J., Peaceman, A. M., Wigton, T. R., Rehnberg, K. A. & Socol, M. L. Fetal fibronectin levels are elevated in maternal plasma and amniotic fluid of patients with severe preeclampsia. Am. J. Obstet. Gynecol. (1995) doi:10.1016/0002-9378(95)90587-1.

29. Jaiswar, S. P., Amrit, G., Rekha, S., Natu, S. N. & Mohan, S. Lactic dehydrogenase: A biochemical marker for preeclampsia-eclampsia. J. Obstet. Gynecol. India (2011) doi:10.1007/s13224-011-0093-9.

30. Muntner, P. et al. Measurement of blood pressure in humans: A scientific statement from the american heart association. Hypertension (2019) doi:10.1161/HYP.0000000000000087.

31. van Rijn, B. B. et al. Maternal TLR4 and NOD2 gene variants, pro-inflammatory phenotype and susceptibility to early-onset preeclampsia and HELLP syndrome. PLoS One (2008) doi:10.1371/journal.pone.0001865.

32. Lundberg, S. M. & Lee, S. I. A unified approach to interpreting model predictions. In Advances in Neural Information Processing Systems (2017).

33. Lundberg, S. M. et al. Explainable machine-learning predictions for the prevention of hypoxaemia during surgery. *Nat*. Biomed. Eng. 2, 749–760 (2018).

34. Geldenhuys, J., Rossouw, T. M., Lombaard, H. A., Ehlers, M. M. & Kock, M. M. Disruption in the regulation of immune responses in the placental subtype of preeclampsia. Frontiers in Immunology (2018) doi:10.3389/fimmu.2018.01659.

35. Davalos, D. & Akassoglou, K. Fibrinogen as a key regulator of inflammation in disease. Seminars in Immunopathology (2012) doi:10.1007/s00281-011-0290-8.

36. Rolnik, D. L. et al. ASPRE trial: performance of screening for preterm pre-eclampsia. Ultrasound Obstet. Gynecol. 50, 492–495 (2017).

37. Blue, N. R. et al. Effect of ibuprofen vs acetaminophen on postpartum hypertension in preeclampsia with severe features: a double-masked, randomized controlled trial. Am. J. Obstet. Gynecol. (2018) doi:10.1016/j.ajog.2018.02.016.

38. Hirshberg, J. S. & Cahill, A. G. Pain relief: determining the safety of ibuprofen with postpartum preeclampsia. American Journal of Obstetrics and Gynecology (2018) doi:10.1016/j.ajog.2018.04.026.

39. Hauspurg, A. et al. Blood pressure trajectory and category and risk of hypertensive disorders of pregnancy in nulliparous women. Am. J. Obstet. Gynecol. (2019) doi:10.1016/j.ajog.2019.06.031.

40. Whelton, P. K. et al. 2017 ACC/AHA/AAPA/ABC/ACPM/AGS/APhA/ASH/ASPC/NMA/PCNA Guideline for the Prevention, Detection, Evaluation, and Management of High Blood Pressure in Adults: Executive Summary: A Report of the American College of Cardiology/American Heart Association Task. Hypertens. (Dallas, Tex. 1979) 71, 1269–1324 (2018).

41. Wilson, E. et al. Varicella Vaccine Exposure during Pregnancy: Data from 10 Years of the Pregnancy Registry. J. Infect. Dis. (2008) doi:10.1086/522136.

42. Rolnik, D. L. et al. Early screening and prevention of preterm pre-eclampsia with aspirin: time for clinical implementation. Ultrasound Obstet. Gynecol. 50, 551–556 (2017).

43. Poon, L. C. et al. The International Federation of Gynecology and Obstetrics (FIGO) initiative on pre-eclampsia: A pragmatic guide for first-trimester screening and prevention. Int. J. Gynecol. Obstet. (2019) doi:10.1002/ijgo.12802.

44. Sotiriadis, A. et al. ISUOG Practice Guidelines: role of ultrasound in screening for and follow-up of pre-eclampsia. Ultrasound Obstet. Gynecol. (2019) doi:10.1002/uog.20105.

45. O’Gorman, N. et al. Competing risks model in screening for preeclampsia by maternal factors and biomarkers at 11-13 weeks gestation. Am. J. Obstet. Gynecol. (2016) doi:10.1016/j.ajog.2015.08.034.

46. Sammour, M. B., El-Kabarity, H., Fawzy, M. M. & Schindler, A. E. WHO Recommendations for prevention and treatment of pre-eclampsia and eclampsia. WHO Recommendations for Prevention and Treatment of Pre-Eclampsia and Eclampsia (2011).

47. Yao, F. et al. Shrinkage estimation for functional principal component scores with application to the population kinetics of plasma folate. Biometrics (2003) doi:10.1111/1541-0420.00078.

48. Wang, J.-L., Chiou, J.-M. & Müller, H.-G. Functional Data Analysis. Annu. Rev. Stat. Its Appl. (2016) doi:10.1146/annurev-statistics-041715-033624.

49. Zou, H. & Zhang, H. H. On the adaptive elastic-net with a diverging number of parameters. Ann. Stat. (2009) doi:10.1214/08-AOS625.

50. Chen, T. & Guestrin, C. XGBoost: A scalable tree boosting system. in Proceedings of the ACM SIGKDD International Conference on Knowledge Discovery and Data Mining (2016). doi:10.1145/2939672.2939785.

51. Lundberg, S. M. et al. From local explanations to global understanding with explainable AI for trees. *Nat*. Mach. Intell. (2020) doi:10.1038/s42256-019-0138-9.

52. Ke, G., et al. LightGBM: A highly efficient gradient boosting decision tree. in Advances in Neural Information Processing Systems (2017).

53. Bergstra, J., Yamins, D. & Cox, D. D. Making a science of model search: Hyperparameter optimization in hundreds of dimensions for vision architectures. in *30th International Conference on Machine Learning*, ICML 2013 (2013).

54. Hyland, S. L. et al. Early prediction of circulatory failure in the intensive care unit using machine learning. Nat. Med. (2020) doi:10.1038/s41591-020-0789-4.

55. Artzi, N. S. et al. Prediction of gestational diabetes based on nationwide electronic health records. Nat. Med. 26, 71–76 (2020).

## References

1. Cowen, M. E. et al. Casemix Adjustment of Managed Care Claims Data Using the Clinical Classification for Health Policy Research Method. Med. Care (1998) doi:10.1097/00005650-199807000-00016.

2. Yetisen, A. K., Akram, M. S. & Lowe, C. R. Paper-based microfluidic point-of-care diagnostic devices. Lab on a Chip (2013) doi:10.1039/c3lc50169h.

3. ACOG Practice Bulletin No. 202: Gestational Hypertension and Preeclampsia. Obstet. Gynecol. (2019) doi:10.1097/AOG.0000000000003018.

4. Shannon, P. et al. Cytoscape: A software Environment for integrated models of biomolecular interaction networks. Genome Res. (2003) doi:10.1101/gr.1239303.

